# COVID-19 Modeling: A Review

**DOI:** 10.1101/2022.08.22.22279022

**Authors:** Longbing Cao, Qing Liu

## Abstract

The unprecedented and overwhelming SARS-CoV-2 virus and COVID-19 disease significantly challenged our way of life, society and the economy. Many questions emerge, a critical one being how to quantify the challenges, realities, intervention effect and influence of the pandemic. With the massive effort that has been in relation to modeling COVID-19, what COVID-19 issues have been modeled? What and how well have epidemiology, AI, data science, machine learning, deep learning, mathematics and social science characterized the COVID-19 epidemic? what are the gaps and opportunities of quantifying the pandemic? Such questions involve a wide body of knowledge and literature, which are unclear but important for present and future health crisis quantification. Here, we provide a comprehensive review of the challenges, tasks, methods, progress, gaps and opportunities in relation to modeling COVID-19 processes, data, mitigation and impact. With a research landscape of COVID-19 modeling, we further categorize, summarize, compare and discuss the related methods and the progress which has been made in modeling COVID-19 epidemic transmission processes and dynamics, case identification and tracing, infection diagnosis and medical treatments, non-pharmaceutical interventions and their effects, drug and vaccine development, psychological, economic and social influence and impact, and misinformation, etc. The review shows how modeling methods such as mathematical and statistical models, domain-driven modeling by epidemiological compartmental models, medical and biomedical analysis, AI and data science, in particular shallow and deep machine learning, simulation modeling, social science methods and hybrid modeling have addressed the COVID-19 challenges, what gaps exist and what research directions can be followed for a better future.

## 1 Introduction

The novel coronavirus (SARS-CoV-2) which causes the COVID-19 disease resulted in not only a global health crisis, but also a global economic and social crisis. Its evolution and influence on the entire world has been profound, overwhelming, unprecedented, and unanticipated. COVID-19 has triggered and has continuously motivated extensive research on its understanding, management, and control. COVID-19 research has been conducted by scientists and researchers from a wide variety of disciplines from almost every country and region in the world [44].

The COVID-19 research landscape is immense, however one critical area is modeling COVID-19. *COVID-19 modeling* aims to quantify, characterize, process, analyze, predict and simulate COVID-19 characteristics and its induced problems, issues and challenges and to extract evidence, insights or indications to understand and manage them. These involve broad problem domains, in particular, in characterizing the intricate nature of COVID-19 and discovering insights for virus containment, disease treatment, drug and vaccine development, and mitigating its broad socioeconomic impact. COVID-19 modeling also involves various disciplines and research areas, including but not limited to statistical modeling, epidemic and health modeling, data-driven analytics and learning, in both shallow and deep, specific and comprehensive manners.

Although many reviews and surveys have emerged increasingly in the literature, particularly those on the applications of AI, machine learning, and deep learning to COVID-19, it is unclear as to what such comprehensive COVID-19 modeling looks like. This motivates this review of the global reaction to modeling COVID-19. Specifically, instigated and informed by our systematic analysis of how global scientists have responded to tackling COVID-19 [44], this paper makes the first attempt to provide a comprehensive and systematic overview of cross-disciplinary and cross-domain research on COVID-19 modeling.

Here, we introduce our review objectives, the research questions to be explored in this review, the review methods, some findings, and the contributions and limitations of this comprehensive review. Fig. 1 shows the meta-synthetic and meta-analytical (MsMa) review method applied in this review. MsMa is built on PRISMA and the NLP-based literature extraction and analysis in our other work on global literature analysis [44]. Interested readers can refer to [44] for more information about reference search, processing, analysis, modeling problem screening, and modeling method screening. The method of representative reference selection is explained in Section 1.2.

**Fig. 1.**
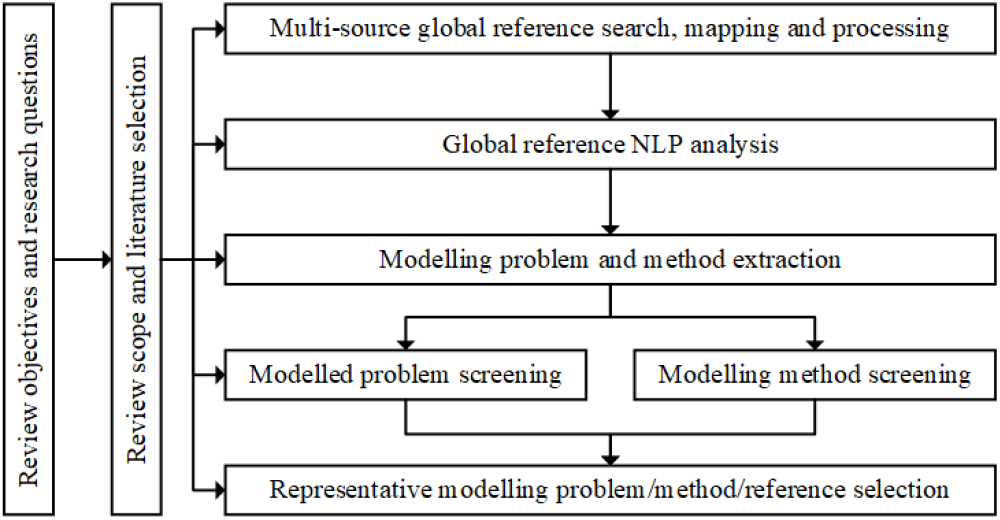
The flowchart of meta-synthetic and meta-analytical (MsMa) review of COVID-19 modeling problems and methods in this review, building on PRISMA and NLP-based literature analysis in [44].

### 1.1 Review Objectives and Questions

In this review, we address a wide spectrum of major problems and aspects associated with the SARS-CoV-2 virus and the COVID-19 disease covering coronavirus evolution and identification, epidemic transmission, COVID-19 disease and infection diagnosis, medical treatment, biomedical analysis, non-pharmaceutical interventions, coronavirus resurgence and mutation, and the influence and impact of the virus and disease. However, we are not motivated to review every research area in relation to all these problems and aspects. Instead, we focus on COVID-19 modeling, i.e., how are the above problems and aspects quantified, processed, or analyzed.

#### 1.1.1 Review objectives

Specifically, we aim to achieve the following objectives by conducting this review:

- generating a comprehensive picture of the major problems and challenges related to the COVID-19 disease;
- understanding the major characteristics and complexities of the data associated with COVID-19;
- generating a conceptual map of the research issues and techniques mostly concerned in modeling COVID-19;
- identifying and summarizing the main techniques and methods in each major modeling discipline for modeling COVID-19;
- identifying and summarizing the main methods in model-based (e.g., mathematical and statistical), domain-driven (e.g., epidemic and medical), and datadriven (e.g., shallow and deep learning) modeling of COVID-19;
- identifying and summarizing the influence and impact of COVID-19 on mental, economic and social aspects;
- identifying and comparing the strengths and weaknesses of major modeling methods in terms of their reported performance in modeling COVID-19;
- identifying the gaps and opportunities for further research on modeling COVID-19 and other similar global crises.

In contrast to the existing reviews and surveys such as [145, 56], this review aims to be:

- more comprehensive than the other references by covering cross-disciplinary problems and techniques for modeling COVID-19;
- analytics and learning-focused where we show statistical machine learning, model-based machine learning, deep learning, and other AI and data science techniques have been widely applied in modeling COVID-19;
- specific to modeling COVID-19, the domain-specific research on its epidemiology, medicine, vaccine, biology and pathology etc. is excluded; in addition, classic typical methods applied in these areas but have not been widely used in COVID-19 research are also excluded;
- unique in critical analysis and summarizing the challenges of the COVID-19 disease, data and modeling by model-, domain- and data-driven approaches;
- structural and critical by categorizing, comparing, criticizing and generalizing typical modeling methods in different disciplines and areas for modeling COVID-19; and
- insightful by extracting conclusive and contrastive (to other epidemics) findings about the virus and disease.

#### 1.1.2 Research questions

To address the above objectives, we identify useful indications, trends and opportunities in relation to the following research questions so that we can generate a comprehensive but also concrete picture of the state-of-the-art research on modeling COVID-19.

- What are the main characteristics and challenges of the SARS-CoV-2 virus and the COVID-19 disease which make it difficult to model COVID-19?
- What does coronavirus data look like and what are the main characteristics and challenges which make COVID-19 data difficult to model and different from other sources of data?
- What are the main objectives of modeling COVID-19?
- What does the COVID-19 modeling landscape look like?
- What are the global research patterns and trends in relation to modeling COVID-19?
- What are the major modeling methods and how do they perform in each major modeling discipline, including mathematical modeling, data-driven learning, and domain-driven modeling?
- Where are the gaps in the domain-driven, model-driven and data-driven methods in modeling COVID-19?
- What influence and impact has COVID-19 had and how are these modeled?
- What lessons can we learn from analyzing the literature on modeling COVID-19?
- What are the knowledge (i.e., addressed but not well) and understanding (yet to be addressed) gaps in modeling COVID-19?

In addition, we are also interested in exploring answers to the following questions:

- What forms the research landscape of COVID-19 modeling, i.e., what major COVID-19 problems have been modeled and what modeling techniques were used to address the COVID-19 challenges?
- How well have AI and data science, specifically data analytics, shallow and deep learning, deepened and broadened the understanding and management of the COVID-19 pandemic?
- Where have AI and data science techniques played a part in modeling COVID-19 problems?
- Where can AI and data science make new, more or better differences in containing COVID-19?

### 1.2 Review Methods

Here, we discuss the review scope, the scope of the modeling methods, and the literature selected in this review. As shown in Fig 1, we utilize a MsMa review method. Our broad experience and understanding of the COVID-19 pandemic and its research prompt the aforementioned review objectives and research questions. They further determine the review scope. Informed by the multi-source global literature search and processing and their natural language processing (NLP) analytical results in [44], systematic and comprehensive modeling problems and methods are included in this review, aiming for a comprehensive landscape of COVID-19 modeling. By scrutinizing and consolidating the modeling-oriented foci and indications from the literature analysis and other reviews on COVID-19, the representative modeling problems, methods and related references are then highlighted in this work for review depth and illustrations.

#### 1.2.1 Review scope

Different from the other surveys and reviews we have identified (such as [145, 56]) in relation to COVID-19, addressing the aforementioned objectives determines the scope of this review. An important guidance on scoping the COVID-19 issues and modeling methods concerned in this review comes from our comprehensive literature analysis of how global scientists have responded to understanding COVID-19 [44].

Accordingly, we identify a broad COVID-19 problem space (as shown in Table 1) and obtain a comprehensive view of how these COVID-19 problems have been modeled and by what respective modeling techniques (as illustrated in Fig. 5). A comprehensive spectrum covers the coronavirus challenges, data issues, modeling techniques, gaps and opportunities in relation to modeling COVID-19. This generates a more comprehensive understanding of the COVID-19 problems and modeling methods than other reviews. However, we do not aim to cover everything related to COVID-19. Instead, our selection of literature and methods is centered on quantifying COVID-19.

**Table 1.** Problems and Objectives Associated with Modeling COVID-19.

| Objectives | Modeling factors | Approaches | References |
| --- | --- | --- | --- |
| Epidemic dynamics and transmission | Epidemiological factors (e.g., origins, incubation period, transmission rate, morbidity, mortality, and highly-to-least vulnerable population), daily new-infected-recovered-death case numbers, reporting time, mobility, the side information about population and their demographics, etc. | Regression, compartmental models, time/age-dependent compartmental models, probabilistic compartmental models, wavelet transformation, simulation methods, etc. | [94, 290, 5, 58, 246, 321, 209, 125, 184, 167, 77] |
| Diagnosis, identification and tracing | Clinical, pathological and genomic data (symptoms, medical facilities, hospitalization records, medical history, respiratory signals, medical imaging like CT and X-ray, physical and chemical measures, gene sequences, and proteins), case number, infection details, patient demographics, mobility, and contacts, etc. | Regression, statistical learning, shallow learning (e.g., decision trees, and random forest), deep neural networks, image and signal processing methods, transfer learning, etc. | [279, 315, 60, 18, 63, 180, 69, 11, 183, 110, 14] |
| Medical treatment and pharmaceutical interventions | Clinical measures, hospitalization records, medical test records, drug selection, drug-target interactions, pharmaceutical treatments, ICU records, ventilator use, health-care records, etc. | Classifiers, time-series methods, deep neural networks, information fusion, etc. | [320, 293, 20, 156, 280, 116] |
| Pathological and biomedical analysis & drug/vaccine development | Genomic data, protein structures, pathogenic data, drug records, protein-disease interactions, drug-target interactions, vaccine information, vaccination transactions, antibody neutralization, immunization response, viral mutation, etc. | Classifiers, outlier detectors, genome analysis, protein analysis, deep neural networks, etc. | [132, 8, 20, 169, 239, 317, 227, 198, 275, 296, 233, 143] |
| Non-pharmaceutical intervention and policies | Intervention, quarantine, mitigation and transmission control measures and policies on populations, communities and individuals, epidemiological factors, daily new-infected-recovered-death case numbers, reporting time, social activities, mobility, communications, and change points | Regression, customized compartmental models, Bayesian hierarchical models, stochastic compartmental models, etc. | [30, 219, 5, 94, 257, 172, 88, 71, 144, 311, 168] |
| Second wave and resurgence | Daily new-infected-recovered-death case numbers, reporting time, second wave/resurgence information, NPIs including quarantine and mitigation measures and policies on populations, communities and individuals, social activities, mobility, seasonal and environmental factors (e.g., season, geographical location, temperature, humidity, and wind speed), etc. | Compartmental models, simulation models, stochastic and regressive compartmental models, epidemic renormalisation group, statistical models, etc. | [151, 12, 36, 163, 206, 218] |
| Mental, socioeconomic influence and impact | Quarantine and mitigation measures and policies on populations, communities and individuals, domain knowledge and precaution guidance from authorities, social activities, mobility, patient demographics (e.g., age, gender, racist, cultural background, and habit), news, reports, social media sentiment, misinformation, vaccination, business data, economic data, logistic data, trade data, public health data, mental health data, and clinical data, etc. | Statistical analysis, qualitative methods such as questionnaire methods, age-structured SIR and SEIR models, deep neural networks (e.g., BERT and LSTM), structural equation models, simulation models, etc. | [48, 141, 274, 53, 104, 93, 136, 271, 233, 19, 106, 243, 23] |

Regarding the depth of the reviewed specific techniques, the broad-reaching scope limits the space for detailing every aspect of the reviewed issues and modeling methods. In each area, we select their representative issues and methods for specific and detailed discussion. For each featured technique, we introduce one general and advanced model/method to represent their family of methods by highlighting their novel designs and mechanisms in handling COVID-19. Instead of detailing classic methods well covered in the related literature, we highlight those more advanced methods with great visibility in the literature and those with hybrid designs and good performance.

#### 1.2.2 Modeling methods

Since COVID-19 involves various problems and issues related to different domains and disciplines, modeling COVID-19 also involves different disciplinary methods. The literature analysis in [44] provides useful hints on the global modeling trends, instigating our selection of the most commonly applied modeling techniques. Accordingly, this review covers a broad-reaching spectrum of modeling techniques and representative progress made by computer scientists (in particular data modelers, analysts and machine learners), medical analytics scientists (in particular, epidemiological, biomedical and medical analysts), and quantitative social scientists (particularly influence modeling) in quantifying COVID-19. We divide the above modeling techniques into three categories:

- *data-driven modeling* including analytics and learning methods;
- *model-based modeling* including mathematical and statistical models; and
- *domain-driven modeling* including representative compartmental models in epidemiology and social science methods for quantifying COVID-19 problems.

In particular, the review also concentrates on AI, data science and machine learning techniques. We explore what and how shallow and deep data analytics and machine learning methods have been applied to address the various issues and problems of COVID-19.

By involving the above empirical aspects and trends of modeling COVID-19, we consider those mostly representative modeling methods. They are categorized into mathematical models, data-driven models, domain-driven models, social science research, simulation methods, and their hybridization, as discussed in Sections 5 to 10.

#### 1.2.3 Literature selection

We first predefine a list of keywords related to COVID-19 modeling, as listed in Appendix 13. We then use these keywords combined with ‘COVID-19’ and two other general combinations: ‘COVID-19 + review’, ‘COVID-19 + survey’, to search for the candidate references. We follow the work in [44]^1^ to analyze and identify those references related to COVID-19 modeling. This results in 43,921 publications on COVID-19 modeling.

The references are then further processed to select a small number of the mostly relevant, quality and representative manuscripts for the inclusion in this paper. Inspired by the guidelines usually followed in health and medical reviews, including the Systematic Reviewers and Meta-analysis (PRISMA) guideline, the Health States Quality-Controlled data, and the general practices in computing surveys, we utilize the MsMa approach shown in Fig. 1 to select the literature. The selected references for this review reflect the following considerations:

- references mainly relevant to those representative modeling techniques highlighted in the findings of global scientists on containing COVID-19;
- references mainly relevant to typical COVID-19 problems and issues that have been widely discussed in the global literature on containing COVID-19;
- references that were documented and mostly compliant with the general practices in the analytics and learning communities and in alignment with their methodologies including data and feature selection, model documentation and evaluation;
- publications appearing in quality peer-review venues to verify their quality (i.e., preprints and publications in low-quality venues are generally excluded); and
- references presenting interesting and representative results in comparison with others.

Further, for the literature on each category of modeling techniques, the ‘representativeness’ of the modeling techniques affects the references to be included:

- the usage intensity and frequencies of modeling techniques in addressing COVID-19;
- the usefulness and results demonstrated by modeling techniques in understanding or handling COVID-19; and
- the novelty of one technique in comparison with other similar methods in the COVID-19 literature.

The above systematic and manual literature selection results in about 300 representative references kept in this paper. However, it is worthy of noting, our reviewed techniques and results in this paper are not limited to these references. In fact, the above empirical guidelines are further informed by the NLP-based analysis of the references on modeling in our other literature analysis [44]. This report informs the general trending patterns and dynamics of problems, issues and techniques concerned by broad modeling communities in quantifying COVID-19.

Although the above guidelines screen the references cited in this paper, many references are not cited because of space considerations. There are a huge number of references available in the body of literature [44]. Much more work is required to select more representative references from the some 50k literature related to AI, data science and machine learning for COVID-19 in particular [44]. We refer interested readers to the appendices in [44] for the metadata of these references and the original sources from which we acquired the full references.

### 1.3 Some Findings

The most important finding of this review relates to the answers to and the observations and insights gained to the question *what does the COVID-19 modeling look like*. It generates a relatively full body of knowledge and a comprehensive conceptual spectrum of this research area: the reality and challenges of the COVID-19 pandemic, data complexities, modeling complexities, the knowledge map of COVID-19 modeling objectives and techniques, the knowledge gaps, and open opportunities in modeling COVID-19. These make this review the most comprehensive and systematic in the COVID-19 literature [44].

Below, we summarize some high-to-low level observations and illustrate some quantitative indications of the above major aspects from this review.

- *COVID-19 problems and complexities*: The research covers the full spectrum of SARS-CoV-2 and COVID-19 problems and issues, including epidemic dynamics and transmission, coronavirus and disease diagnosis, infection identification, contact tracing, virus mutation, resurgence, medical diagnosis and treatment, pharmaceutical interventions, pathological and biomedical analysis, drug and vaccine development, non-pharmaceutical interventions, and socioeconomic influence and impact. The COVID-19 disease characteristics and complexities include complex hidden epidemic attributes, high contagion, high mutation, high proportion of asymptomatic-to-mild symptomatic infections, varied and long incubation periods, ethnic sensitivity, and other high uncertainties, showing significant differences between SARS-CoV-2 and other existing viruses and epidemics.
- *COVID-19 data and challenges*: The literature involves comprehensive COVID-19 data, including (1) core data such as daily confirmed, recovered and deceased cases, daily asymptomatic infections, pathological, clinical and genomic results of virus and disease tests, genomic analysis, drug-target matching, and patients’ demographics, healthcare activities, hospitalized information, and public health services; (2) intervention and response data: such as non-pharmaceutical intervention (NPI) policies and events, vaccination, welfare and human service response, people’s responses, behaviors, public activities, mobility/transport, and business activities; (3) external data such as social media, Q/A, questionnaires, weather, environment, misinformation, and news. This comprehensive COVID-19 data spectrum also involves almost all the data complexities widely explored in modeling, analytics and learning, including data uncertainty, dynamics, nonstationarity, data quality issues such as incompleteness, inconsistency, inequality and incomparability, lack of ground truth information, and limited and inconsistent daily reporting.
- *COVID-19 modeling challenges*: The COVID-19 complexities and their data challenges bring about various modeling issues and challenges, including producing a universal representation of the data genomics of COVID-19, involving cross-disciplinary knowledge in creating a unified COVID-19 quantification, enabling multi-objective and multi-task modeling of comprehensive problems, modeling low-to-poor quality COVID data, modeling small and limited COVID data, COVID learning with weak-to-no prior knowledge and ground truth, modeling hierarchical and diverse forms of COVID heterogeneities, modeling weak-to-hidden couplings and interactions between multi-source, multimodal and external COVID data, disclosing hidden and unknown attributes and dynamics of the virus and disease over time, region, and context, and applying overparameterized and pretrained deep neural networks on often small but complex COVID data.
- *COVID-19 modeling techniques*: The COVID-19 modeling landscape is comprehensive and systematic and covers almost all modeling methods from a wide disciplinary body of knowledge, including conventional mathematical and statistical modeling, simulation methods, epidemiological and epidemic modeling, modern data-driven discovery, shallow and deep machine learning, evolutionary computing methods, event and behavior modeling, and social science methods such as psychological and economic analysis methods.
- *COVID-19 disciplinary modeling*: Of the 346k publications collected on COVID-19, about 44k publications (and extra 7k preprints) are on modeling COVID-19 [44]. In these modeling publications, about 14%, 8% and 50% of publications are from computer science, social science, and medical science, respectively.
- *COVID-19 mathematical and statistical modeling*: There are about 25k references on mathematical and statistical modeling, where regression models including linear, logistic, Cox and Poisson regression are mostly applied, with univariate, multivariate and Bayesian statistics ranking second. The COVID-19 problem keywords which are most frequently used are risk factor, healthcare, anxiety, lockdown, vaccine, depression, stress, vaccination, distress, and mortality rate.
- *COVID-19 epidemic modeling* : Compartmental models are involved in over 5.6k publications, with the most used models being susceptible-exposed-infectious-removed (SEIR), susceptible–infectious–recovered (SIR), susceptible–infectious–recovered–dead (SIRD), and growth models. The COVID problem keywords which are most used are lockdown, reproduction number, policy, intervention, vaccine and vaccination, healthcare, social distancing, control measure, and transmission rate.
- *COVID-19 medical and biomedical modeling*: There are over 21k publications from medical science, where the techniques which are most frequently applied are regression models including logistic, linear and Cox regression, multivariate statistics, simulation, statistical models, machine learning, artificial intelligence, and Bayesian models. The COVID-19 problem keywords which are most used are risk factor, healthcare, vaccine and vaccination, anxiety, depression, stress, distress, screening, and CT.
- *COVID-19 machine learning*: Of the 44k modeling publications, over 8.5k publications are on shallow and classic machine learning, where support vector machine (SVM), sentiment analysis, random forest, classification, decision tree, clustering, natural language processing (NLP), artificial neural networks (ANN) are the most reported techniques. The COVID-19 problem keywords which are most used in these machine learning publications are healthcare, X-ray, CT, lockdown, vaccine, tweets, policy, social media, lung and vaccination. In contrast, over 4.5k references investigated deep learning methods, including convolutional neural networks (CNNs), neural networks, transfer learning, long-short term memory (LSTM), ResNet, VGG, deep neural networks (DNNs), recurrent neural networks (RNNs), and MobileNet. The COVID-19 problem keywords which are most frequently used are X-ray, CT, lung, screening, healthcare, monitoring, vaccine, RT-PCR, and lockdown.
- *COVID-19 modeling tasks*: On one hand, modeling tasks address almost all COVID-19 problems and issues [44], including 4.4k publications related to mental health, anxiety, depression and stress, 1.5k related to the second wave, 2k related to lockdown and social distancing, 2.4k related to vaccine and vaccination, 1.2k related to risk factors, and 3.3k on the prediction of COVID-19 spread, cases, and mortality, 5.5k on epidemic modeling, 5.6k on diagnosis and identification, 2.8k on influence and impact, 3k on simulation, and 0.7k on resurgence and mutation. On the other hand, the literature covers an overwhelming number of analytical and learning tasks including roughly 1.3k on unsupervised learning and clustering, 2.6k on classification, 0.2k on multi-source and multi-modal data modeling and multi-task learning, and 3k on forecasting and prediction.
- *Epidemic attributes*: as summarized in Section 3.2 on COVID-19 disease characteristics, it is estimated that the reproduction number (probably larger than 3 in the original wave and over 2 in the resurgence after receiving vaccination) is much higher than SARS and MERS, the incubation may last for an average of 5 to even beyond 14 days, the asymptomatic infections may be much higher than 20% with even up to 80% undocumented infections in some countries, and some virus mutants may increase the transmission rate by more than 50% over the original strain.
- *COVID-19 learning performance*^2^: As illustrated in Section 6.2, COVID-19 shallow learning references report an accuracy of over 90% in predicting COVID-19 outbreaks, over 96% in disease diagnosis on clinical reports, over 98% in diagnosis on medical images, and close to 99% in diagnosis further involving latent features with specific settings and data. As detailed in Sections 6.3 and 7.2, in the references on COVID-19 deep learning and medical imaging analysis, DNN variants achieve significant prediction performance on COVID-19 images and signals, e.g., an accuracy of over 92% on cough sound using LSTM, over 99% on chest X-ray images using CNN and ImageNet variants, and less than 5% MAE on real unlabelled lung CT images using attention and gated U-Net.
- *COVID-19 non-pharmaceutical intervention effect*: As detailed in Section 8.1, modeling the effect of COVID-19 interventions and policies, NPIs such as business lockdowns, school closures, limiting gatherings, and social distancing were shown to be crucial in containing virus outbreaks and reducing COVID-19 case numbers, e.g., reducing the reproduction number by 13%-42% individually or even 77% jointly using these control measures, and resulting in over 40% transmission reduction by restricting human mobility and interactions.
- *COVID-19 mental, social and economic impact* : In the literature, mental health, anxiety, stress, and depression form the top concerns, showing the overwhelming negative COVID-19 impact on public mental health. The pandemic also had a significantly diverse impact on economic growth and the workforce (e.g., over 20% estimated annual GDP loss in 2020), public health systems, global supply chains, sociopolitical systems, and information disorder.

In addition, this review also identifies significant gaps and opportunities in modeling COVID-19. On one hand, as detailed in Section 11.1, the review finds various issues and limitations in the existing research, e.g., an insufficient, biased and partial understanding of COVID-19 complexities and data challenges; a simple and direct application of modeling techniques on often simple data; the lack of robust, generalizable and tailored designs and insights into the virus and disease nature and complexities. On the other hand, as discussed in Section 11.2, we also identify various opportunities for AI, data science, and machine learning in relation to COVID-19. Significant new opportunities include (1) studying rarely to poorly addressed problems such as epidemiologically modeling mutated virus attributes, complex interactions between core and external factors, and the influence of external factors on epidemic dynamics and NPI effect; (2) developing new directions and methods such as hybridizing multiple sources of data or methods to characterize the complex COVID systems; and (3) enabling novel AI, data science and machine learning research on the large-scale simulation of the intricate evolutionary mechanisms in COVID, discovering robust and actionable evidence to dynamically personalize the control of a potential resurgence and balance economic and mental recovery and virus containment.

### 1.4 Contributions and Limitations

We aim to make the first attempt at creating a comprehensive and concrete picture of COVID-19 modeling.

#### 1.4.1 Contributions

COVID-19 has had a devastating impact on every aspect of life, however, there is currently no research available on how global scientists have responded to this unprecedented global crisis. Accordingly, to the best of our knowledge, this is the first attempt to address this expectation^3^. It extracts the comprehensive and structured research landscape of what problems and issues related to COVID-19 have been widely discussed in the literature, how the coronavirus and COVID-19 have been quantified in the broad disciplines, and what are the typical modeling methods favored in the literature.

The review extracts various insights about modeling COVID-19 in terms of the global trends, representative methods across disciplines, and typical performance from those reported applications. It also incorporates much discussion on the topics, methods, gaps, opportunities, and directions to tackle those issues which are rarely or poorly addressed and areas which remain open in the broad research landscape of modeling COVID-19.

The review is organized as follows. We start by categorizing the characteristics and challenges of the COVID-19 disease, the data and the modeling in Section 3. A transdisciplinary landscape is formed to categorize and match both COVID-19 modeling tasks and objectives and categorize the corresponding methods and general frameworks in Section 4. The review then focuses on structuring, analyzing and comparing the work on mathematical, data-driven (shallow and deep machine learning), domain-driven (epidemic, medical and biomedical analyses) modeling in Sections 5, 6 and 7, respectively. Section 8 further discusses the modeling on the influence and impact of COVID-19, Section 9 reviews the work on COVID-19 simulations, and the related work on COVID-19 hybrid modeling is reviewed in Section 10. Lastly, Section 11 discusses the significant gaps and opportunities in modeling COVID-19.

#### 1.4.2 Limitations

However, this review also presents the various limitations of the existing work and highlights opportunities for further work and for the readers’ attention.

As discussed in Section 1.1, this review aims to be comprehensive, specific, unique, structural, critical and insightful. Achieving this aim requires a broad body of knowledge and an extensive volume of literature, consequently making the review very comprehensive, somehow unfocused as in the topic-specific reviews, and shallow in the sense it does not delve deeply into every reviewed area and modeling technique.

We also do not verify the actionability and operationability of all the cited methods and their results in terms of their epidemic, clinical, socioeconomic, or other practical tests and applications, as in [230] and suggested in [73]. The reference selection is not centered on the practical applicability and test as in [230]. We thus do not enforce guidelines such as PRISMA in screening the references. Further, we do not undertake our own quantitative evaluation and validation of each method and reference cited in this review as in, for example, [295]. Hence, the performance reported in this review is from their publications directly by assuming that this has been checked by their peer-review process.

This leaves a major deficit in the actionability of these review findings, i.e., whether and where these well-performing models and their reported results are problematic (e.g., with methodological, design, setting, data or evaluation biases). We thus refer interested readers to practical codes [73] and other practice-related reviews [295, 230] for their evaluation and suggestions of machine learning models for COVID-19 diagnosis and practical tests.

In addition, we also direct the readers’ attention to the potential issue in the fairness, bias, evaluation, accountability, transparency and interpretability of their methodologies, designs, data manipulation, evaluation methods, and result presentation. Interested readers can refer to the relevant discussions on statistical, AI, data-driven, and machine learning methods and their reproducibility, actionability, code of conduct, and regulation in practical tests and applications (e.g., [45, 191, 40, 174, 204, 73, 42].

In addition, this review can also be further improved in various other aspects. (1) As the scope and capacity of the review is limited, we do not cover the domain-specific literature on pure medical, biomedical and social science-oriented topics and methods without involving modeling methods and without addressing COVID-19. This thus filters out many well-recognized techniques in their domain-specific communities, for instance, Markov chain Monte Carlo (MCMC)-based Bayesian inference is widely appreciated in epidemiology, whereas here we only introduce those techniques involving COVID-19. (2) There are over 50k references applying or discussing the role of AI and data science in addressing COVID-19 by researchers from medical, computer and social science communities [44]. It is not possible to fully cover or highlight these in detail in this review. Instead, we collect, categorize and summarize the representative and trending techniques and methods, and encourage interested readers to refer to the metadata of the categorized references and even the sources where we crawled the literature in [44] and its appendix. (3) As discussed in the above, different from the narrowly-focused review papers in the area which highlight specific techniques and their relevant references, we categorize and formulate those mostly applied and novel modeling techniques with interesting and good-performing results. We summarize their representative, novel and generalizable formulations from each major family of modeling techniques, which are expected to capture the typical and major frameworks, designs and mechanisms of their methods. This also indicates that we neither introduce all or most of the methods in each family nor present all details of the models. (4) This review does not answer many important questions concerning modelers, governments, policy-makers, and domain experts, e.g., what has the modeling disclosed to us about the nature of COVID-19 and whether the results reported in the literature disclose the reality of COVID-19, which is complicated and requires more purposeful reviews and modeling studies. (5) There are many challenging problems yet to be informed or addressed by the existing modeling research, as discussed in Section 11. (6) There are increasingly more and newer references including preprints emerging online every day, which pose significant challenges to instantly reflect the up-to-date important references and their contributions to modeling COVID-19.

## 2 Related Work

We briefly summarize the relevant reviews from general and modeling-related perspectives. The work on modeling COVID-19 is then reviewed in terms of the general applications of AI and machine learning methods, specifically concerning COVID-19 problems, specific modeling techniques, and practical issues of applying machine learning in COVID-19 practice.

First, various reviews focus on generating a general overview and understanding of various non-modeling-oriented, domain-specific (e.g., medical, health, biological, and social) areas of COVID-19. Examples are the review of COVID-19 characteristics [81], epidemiology [205], epidemiology, virology and pathogenesis [277], serological tests [155], diagnosis [309], asymptomatic and presymptomatic infection [34, 142], long-term effects [164], non-pharmaceutical interventions [211], mental health [299], and social media [260]. Readers interested in such areas may refer to them for more information, however, they go beyond the scope and focus of our review.

Second, few general reviews are available on the applications of broad AI, machine learning and quantification techniques in COVID-19. The review in [56] lists and categorizes the applications of various AI techniques in COVID-19 detection, diagnosis, virology, pathogenesis, drug and vaccine development, and coronavirus transmission and prediction. The review in [43] provides a brief summary of the challenges, global AI research, the modeling trends and gaps in combating COVID-19. The report provided by AAAS [1] provides a brief summary of selective applications and the impact of AI in fighting COVID-19 in particular in the US. It briefly discusses some AI applications and classifies them into five classes: forecasting, diagnosis, containment and monitoring, drug development and treatments, and social and medical management. The report then focuses on major ethical and human rights frameworks and concerns in relation to medical AI applications in fighting COVID-19. They involve autonomy, beneficence, nonmaleficence, justice, privacy, confidentiality, equality, and non-discrimination and the legal, regulatory and societal issues in their implementation and practice. However, none of these reviews provide a comprehensive and detailed coverage, critical analysis, or technical evaluation of broad AI for COVID-19.

Many reviews and surveys have emerged over time to discuss the applications of specific AI and machine learning methods in fighting COVID-19. They typically focus on specific topics and areas in modeling COVID-19. Examples are epidemic and transmission forecasting [226], asymptomatic transmission [35], virus detection, spread prevention, and medical assistance [241], COVID-19 case prognosis, analysis and tracking [228], diagnosis and prognosis prediction [295], imaging data segmentation and diagnosis [242], and drug and vaccine development [132]. None of them cover all aspects with review depth and width as in our review.

Most reviews focus on specific AI, machine learning, and modeling methods. They include mathematical and statistical modeling [182], epidemiological modeling [205], general AI and machine learning methods [132, 182, 56, 228], data science [145], computational intelligence [261], computer vision and image processing [264, 242], and deep learning [312, 121]. No reviews cover all methods from a systematic perspective as in our review.

It is worth noting that almost all reviews focus on the summarization and categorization of the related work on COVID-19. No work has yet been found on re-evaluating the results and performance reported in the literature. Instead, few references focus on the meta-analysis of the documentation, validity, applicability and bias etc. issues of AI and machine learning models and their results for COVID-19 clinical practice by employing certain meta-analysis guidelines and practical test criteria. In [230], the authors discussed various common pitfalls and issues existing in the machine learning literature on COVID-19 detection and prognosis. By applying the systematic reviews and meta-analyses (PRISMA) checklist and the author’s full-text review of the few eligible selected references, the various issues in most references were pointed out, in particular, those related to presentation quality, detailed data, feature and model specifications for reproducibility, validation, and the risk of bias. In [295], the authors reviewed and appraised the validity and usefulness of predictive models for COVID-19 diagnosis and prognosis. They identified that almost all models suffer from problems including poor and nontransparent documentation, the risk of bias, and problematic performance, therefore, they are invalid for direct and widespread clinical applications. It is noted that such concerns may conflict with more optimistic opinions of AI and machine learning for COVID-19 [266] and the widespread commercial and enterprise analytics [42]. Many identified issues and concerns could be substantially addressed by enforcing the health practical guidelines and codes [73] and applying actionable knowledge discovery methodologies and processes [45, 39].

Overall, a major gap in these reviews is that they only paint a partial picture of what happened in their selected areas based on several references, specific techniques, or particular review purposes. There are no comprehensive surveys or critical analyses of the intricate challenges posed by the virus, the disease, the data, and their modeling from a cross-disciplinary and cross-domain perspective. Our review aims to address these issues.

## 3 COVID-19 Pandemic, Characteristics and Complexities

The COVID-19 pandemic is a complex physical-social-technical system which also involves cyberspace [43]. Accordingly, COVID-19 modeling has to address the general and domain-specific complexities, challenges and research questions in the continuum, take advantage of the advances in the related physical, social, technical and cyber sciences, and involve domain-specific knowledge and factors.

Here, we briefly delineate the COVID-19 pandemic status and summarize the main characteristics and challenges of the COVID-19 disease, the data challenges, and the challenges in modeling COVID-19. Fig. 2 summarizes the major challenges of COVID-19 in terms of the virus and disease, data, and modeling.

**Fig. 2.**
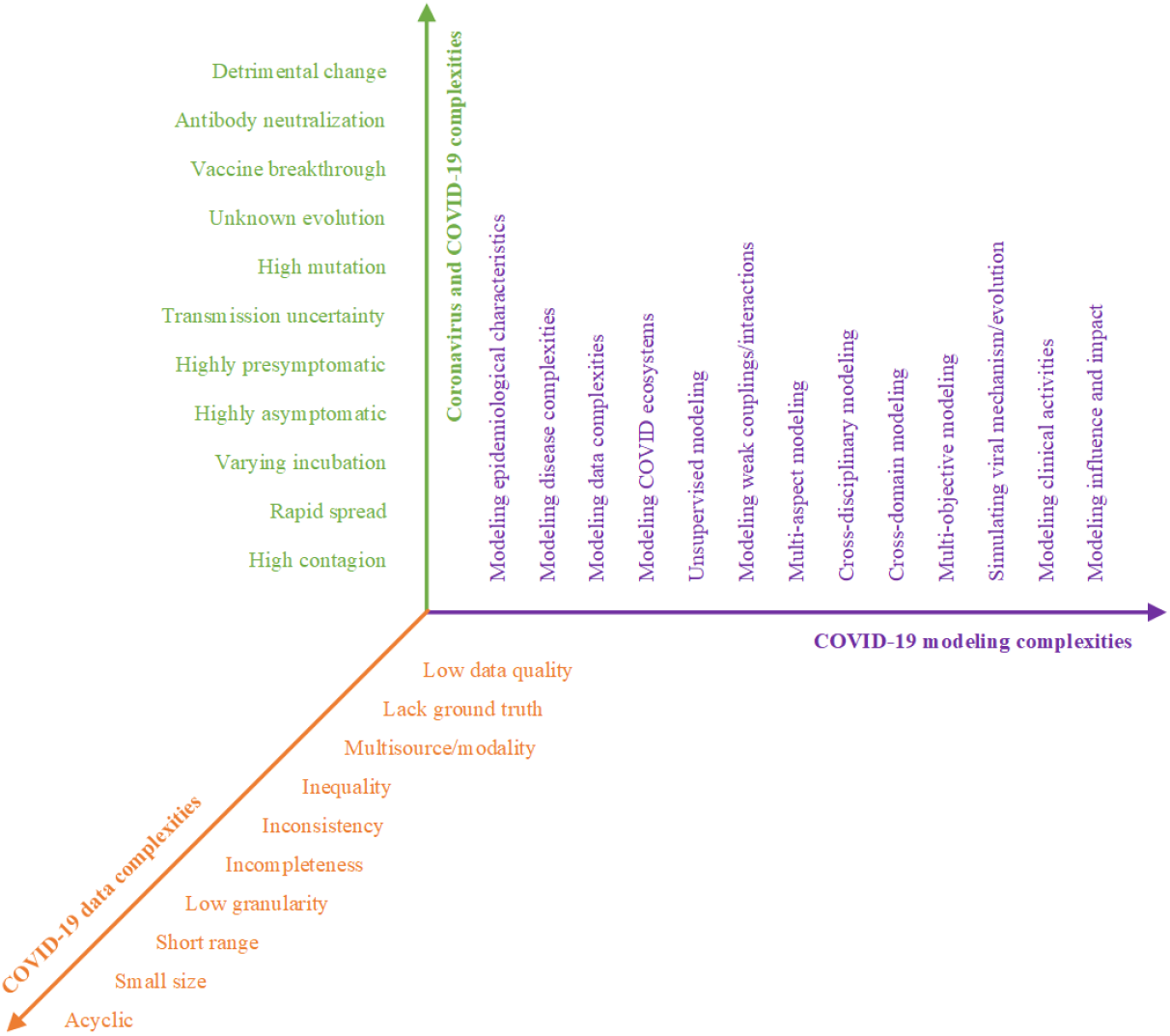
Three-dimensional COVID-19 complexity space: virus and disease, data, and modeling.

### 3.1 COVID-19 Pandemic

The coronavirus disease 2019, designated as COVID-19, is caused by the severe acute respiratory syndrome coronavirus 2 (SARS-CoV-2) virus^4^. SARS-CoV-2 and COVID-19 have overwhelmingly shocked and shaken the entire world. After the first outbreak in Wuhan China in Dec 2019, the disease spread rapidly across the entire world in only two months due to its strong human-to-human transmission ability. The World Health Organization (WHO) declared COVID-19 a pandemic on 11 March 2020. To date, COVID-19 has infected more than 577M people with 6.4M confirmed to have lost their lives^5^. The continuously and iteratively mutative infections have been even more serious, posing a severe threat to 228 countries and territories. The coronavirus has experienced various generations of resurgences and mutations, which continuously challenge pandemic containment, vaccination, and treatments.

With the widespread-to-cluster infections and resurgences, continuously evolving and increasingly contagious mutations from Alpha, Beta, Gamma, Delta to Omicron (currently the strain BA.5)^6^, and the slow, sparse and unbalanced rollouts of global vaccinations for global herd immunity, COVID-19 has continuously and fundamentally transformed global public health, the economy and society with an increasingly unprecedented impact on every aspect of life and the world. COVID-19 has not only exerted an unprecedented pressure on global healthcare systems, but also fundamentally challenged the relevant scientific research on understanding, modeling, diagnosing and controlling the virus and disease and its huge impact. Many questions and challenges arise about COVID-19 in relation to the nature of the coronavirus; the virus’ epidemic characteristics, transmission and influence processes; the disease’s medical and genomic characteristics, dynamics and evolution; and the pros and cons of existing containment, diagnosis, treatment and precaution strategies.

Compared with the epidemics that have been defeated in recent decades, such as the severe acute respiratory syndrome (SARS) and the Middle East respiratory syndrome (MERS), SARS-CoV-2 and COVID-19 are much more complicated and more transmissible from human to human and more uncertain, evolving, contagious, and exceptional [195]. Its unique and exceptional uncertainty, transmission and mutation form some of the key factors that make COVID-19 a continuous, unprecedented and evolving pandemic. Another key challenge of COVID-19 is its long incubation period (ranging from 1 to 14 days or even longer for its early strains like Delta) and high asymptomatic proportion. In the incubation period, infectious individuals are contagious but show no symptoms. Consequently, susceptible individuals may not be aware that they have been infected, hindering their timely identification, contact tracing, and containment. Unfortunately, the number of asymptomatic and mild symptomatic infections is significant over all coronavirus strain-based infections. Asymptomatic infectives are a concerning hidden source of the widespread infection and resurgence of COVID-19 [142], making it difficult-to-impossible to be absolutely clear about their source, diagnosis and mitigation. This brief comparison poses challenging questions to be answered scientifically, including how COVID-19 differs from other epidemic or endemic diseases, what forms the realities of the asymptomatic phase and infection, what are more effective methods to contain and treat the virus and disease, and how the virus mutates and reacts to vaccines and the environment.

Specifically, to slow the pandemic and bring infections under control, most governments implemented many NPIs during the outbreak. Typical NPIs include social distancing, school and university closure, infective isolation or quarantine, banning public events and travel, and business lockdown, etc. These interventions unfortunately incur significant socioeconomic costs and have an adverse impact on businesses. Their practical implementation and management have thus triggered various debates including on the appropriate trade-off between epidemic control and the negative impact introduced by NPIs and between strict mitigation (COVID zero or zero COVID) and herd immunity over the virus evolution. It is quantitatively unclear as to what makes a better trade-off, what are the best timing and appropriate extent of mitigation, what is the positive and negative impacts of NPIs on pandemic control and socioeconomic wellness, and how has COVID-19 influenced other aspects of public life, work, mental and medical health, and the economy on both individual and population (e.g., a city, country to the globe) levels.

### 3.2 Coronavirus and COVID-19 Complexities

Containing and modeling COVID-19 is highly challenging because its epidemiological, clinical and pathological characteristics are sophisticated, evolving and poorly understood^7^ [112, 115, 205]. Despite common epidemic clinical symptoms like fever, cough, tiredness and loss of taste or smell [124], SARS-CoV-2 and COVID-19 have many other sophisticated characteristics [195] that make them more mysterious, contagious and challenging for quantification, modeling and containment. We summarize a few of these below.

#### High contagiousness and rapid spread

The high contagiousness of SARS-CoV-2 is one of the most important factors driving the COVID-19 pandemic. In epidemiology, the *reproduction number R*_0_ denotes the transmission ability of an epidemic or endemic. It is the expected number of cases directly generated by one case in a population where all individuals are susceptible to infection [91]. If *R*_0_ *>* 1, the epidemic will begin to transmit rapidly in the population, while *R*_0_ *<* 1 indicates that the epidemic will gradually vanish and will not lead to a large-scale outbreak. Different computational methods have estimated and produced varying reproduction values of COVID-19 in different regions. For example, Sanche et al. [235] report a median *R*_0_ value of 5.7 with a 95% confidence interval (CI) [3.8, 8.9] during the early stages of the epidemic in Wuhan China. Gatto et al. [92] estimate a generalized reproduction value of 3.60 (95% CI: 3.49 to 3.84) using the SEIR-like transmission model in Italy. de Souza et al. [70] report a value of 3.1 (95% CI: 2.4 to 5.5) in Brazil. The review finds that the *R*_0_ of COVID-19 may be larger than 3.0 in the initial stage, higher than that of SARS (1.7-1.9) and MERS (*<* 1) [213]. It is generally agreed in the literature that SARS-CoV-2 is more transmissible than the severe acute respiratory syndrome coronavirus (SARS-CoV) and the Middle East respiratory syndrome coronavirus (MERS-CoV) although SARS-CoV-2 shares 79% of the genomic sequence identity with SARS-CoV and 50% with MERS-CoV [166, 81, 212, 115].

#### A varying incubation period

The *incubation period* of COVID-19, also known as the pre-symptomatic period, refers to the time from becoming infected by exposure to the virus and symptom onset. For the early virus variants like Alpha and Beta, a median incubation period of approximately 5 days was reported in [147], which is similar to SARS. In [205], the mean incubation period was found to range from 4 to 6 days, which is comparable to SARS (4.4 days) and MERS (5.5 days). Although an average incubation period of 5-6 days is reported in the literature, the actual incubation period may be as long as 14 days [195, 308, 147]. Unlike SARS and MERS, COVID-19 infected individuals are already contagious during their incubation periods. As it is likely that they are unaware of being infected and have no-to-mild symptoms during this period, they may easily become unknown sources of widespread transmission. This characteristic has informed screening and control policies, e.g., mandatory 14-day quarantine and isolation, corresponding to the longest predicted incubation time, for the ancestral strains. However, with virus mutation, incubation periods vary and shorten over generations. Omicron has an average of 3 days after hospital admission, in comparison to 4 days for the Delta strains and 5-6 days for Alpha [248]. The widely varying COVID-19 incubation periods and their uncertainty in specific hotspots and with particular strains make their source tracing, case identification and infection control very difficult.

#### A large proportion of asymptomatic, presymptomatic and undocumented infections

This shows that COVID-19 has a broad clinical spectrum which includes asymptomatic, presymptomatic and mild illness [50, 152, 212, 194]. Asymptomatic and presymptomatic infections may not be screened and diagnosed before symptom onset, leading to a large number of undocumented infections and the potential risk of contact with infected individuals [142]. The review in [35] reports that, of those who tested positive in the studies conducted in seven countries, the proportion of asymptomatic cases ranged from 6% to 41%. The study in [289] reports that 23% of those infected were asymptomatic. Buitrago-Garcia et al. [34] found that most people who were infected did not remain asymptomatic throughout the course of infection, and only 20% of infections remained asymptomatic during follow-up. In contrast, the study in [154] shows that a large percentage (86%) of infections are undocumented, about 80% of documented cases are due to transmission from undocumented cases, and the transmission rate of undocumented infections is about 55% of that of documented cases. However, these estimates require further larger-scale verification and study. The accurate proportion of asymptomatic, presymptomatic and mild-symptomatic transmissions and infections of different generations of coronaviruses remains unclear. It is also unclear whether the higher contagiousness of new variants like Delta and Omicron is associated with their higher asymptomatic and presymptomatic transmissions and how the asymptomatic, presymptomatic and mild-symptomatic transmissions of new strains evolve over their mutation.

#### High mutation and high contagion with mysterious strains

The up-to-date five major SARS-CoV-2 variants of concern are B.1.1.7 (labeled Alpha), B.1.351 (Beta), P.1 (Gamma), B.1.617.2 (Delta), and B.1.1.529 (Omicron)^8^. Research shows their mutations likely result in increased transmissibility, immunity and severity for most mutated lineages and mutations^9^ For example, Beta variants have higher transmissibility (B.1.1.7 has approximately 50% increased transmission than the original variant) [220] and reproduction rate (B.1.1.7 has an increased reproduction rate of 1-1.4) [273], challenging the vaccines and containment and mitigation methods. The basic reproduction number (R0) of the variant B.1.617.2 (Delta) is between 3.2 and 8 [160]. The transmissibility of the Omicron variant is about 3.2 times than that of Delta, and the doubling time is approximately 3 days [162]. Overall, the identified variants of concern generally have increased transmissibility, increased detrimental change of epidemiology, and more severe virulence and disease presentation (e.g., increased hospitalizations or deaths). In general, they result in the decreased effectiveness of public health and social measures, reduced effectiveness of available diagnostics, vaccines and therapeutics, increased diagnostic detection failures, and reduced neutralization by antibodies and vaccine breakthrough generated during previous infection or vaccination^10 11^.

##### Discussion

While the above observations summarize the most recent understanding of SARS-CoV-2 and COVID-19 complexities, it is also noted that our knowledge about the nature of the coronavirus and its lineages and mutations is still limited and nondeterministic. Without knowing its origins and with the often limited and specific observation-based studies, there could be much misinformation and biased characterization of the virus and its evolution and impact on transmissibility, immunity, and severity as well as on vaccination and the NPIs required [232]. There is weak to no ground truth about the realities of infections, symptoms shown in medical imaging, and appropriate mitigation and treatment measures. There have been no joint global pathological, epidemiological, biomedical and socioeconomic studies which provide a deep and systematic understanding of the COVID-19 virus and disease complexities, common knowledge, and ground truth.

### 3.3 COVID-19 Data Complexities

COVID-19 involves multisource, multimodal, internal and external factors and often small, sparse and quality-inconsistent data^12^ [145, 44]. Typical data sources and factors in relation to COVID-19 include but are not limited to:

- epidemiological factors (e.g., origin, incubation period, transmission rate, mortality, morbidity, and high to least vulnerable population, etc.);
- daily new-infected-recovered-death case numbers, their reporting time and region of occurrences;
- clinical, pathological and genomic data (e.g., symptoms, medical facilities, hospitalization records, medical history, medical imaging, pharmaceutical treatments, gene and protein sequences);
- domain knowledge and precautionary guidelines from authorities on the virus and disease;
- infective demographics (e.g., age, gender, race, cultural background, and habit);
- vaccination data (e.g., vaccine information, vaccination transactions, and vaccination effects);
- quarantine and mitigation measures and policies (e.g., social distancing and border control) relating to communities and individuals;
- social activities and mobility of COVID-19 patients and infectives;
- seasonal and environmental factors (e.g., season, geographical location, temperature, humidity, and wind speed);
- news, reports and social media discussions on coronavirus and COVID-19;
- social, economic, cultural, political etc. factors related to the COVID-19 impact;
- mental, emotional, sentimental and psychological effects; and
- fake news, rumor and misinformation.

Such COVID-19 data are heterogeneously coded in character, text, number or image; in unordered, temporal/sequential or spatial modes; in static or dynamic forms; and with varied characteristics. Such data also present significant complexities challenging the existing research on COVID-19 and direct applications of modeling methods in COVID-19. The main characteristics of COVID-19 data are summarized below.

#### Acyclic, small-size and short-range case numbers

The publicly available data on COVID-19 case numbers are limited. Except in rare scenarios such as in the US, most countries and regions report a short-range (2-3 months or even shorter such as for local hotspot-based outbreaks), low-granularity (typically daily), and small-size (daily case numbers for a short period of time and a small population of the outbreak clusters) record of core COVID-19 data. Such data are typically acyclic without the obvious seasonal or periodical patterns as of influenza [65] and recurrent dengue epidemics in tropical countries [265].

#### Problematic statistics of COVID-19 cases

The reported new-infected-recovered-death case numbers are estimated to be much lower than their real values in most countries and regions. This may be due to many reasons, such as presymptomatic and asymptomatic infections, limited testing capability, non-standard manual recording, different confirmation standards, an evolving understanding of the nature of the virus and disease, and other subjective factors. The method of calculating case statistics may vary significantly from country to country. The actual figures in some countries and regions may even be unknown. No clear differentiation is made between hotspot and country/region-based case reporting. Such gaps between the infection reality and what has been documented may be more apparent in the first wave, in the early stage of outbreaks, and in some countries and regions [172]. As result, the actual number of infections and the number of regions affected by the COVID-19 pandemic may be much higher than those publicly reported.

#### Lack of high-quality micro-level data

The number of COVID-19 cases, including daily infected cases, daily new cases, daily recovery cases, and daily death cases, are collected on a daily basis in most countries and regions, while daily susceptible case numbers were also reported in China, including the first wave in Wuhan. The macro-level and low-dimensional data are far from comprehensive for inferring the complex transmission processes accurately. More fine-grained data with various aspects of features and high dimensions are needed. For example, during the initial phase of the Wuhan outbreak, the dissemination of SARS-CoV-2 was primarily determined by human mobility in Wuhan. However, no empirical evidence on the effect of key geographic factors on local epidemic transmission was available [225]. The risk of COVID-19 death varies across various sociodemographic characteristics [76], including age, sex, civil status, individual disposable income, region of residence, and country of birth. More specific data is required to address the sociodemographic inequalities of those contracting the coronavirus. To contain the spread of COVID-19, different governments propose and initiate a series of similar-to-different NPIs. No quantitative evidence or systematic evaluation analyzes how these measures precisely affect epidemic transmission, leading to challenges in inferring NPI-based COVID-19 transmission and mitigation.

#### Data incompleteness, inconsistencies, inequality and incomparability

Typically, it is difficult to find all-round information about COVID-19 patients’ infection source, their demographics, behaviors, social activities (including mobility and in social media), clinical history, diagnoses and treatments, and reinfections. COVID-19 public data also presents strong inconsistencies and inequalities across reporting hotspots, countries and regions, their reporting and updating frequencies and timelinesses, case confirmation standards and criteria, collection methods, and stages and conditions of case confirmation [231]. Data from different countries and areas may be unequal and incomparable due to their non-unified statistical criteria, confirmation standards, sampling and coverage methods, health and medical conditions and protocols. Other inconsistent areas include a patient’s race, life habits, and mitigation policies enforced on them or their living areas.

#### Lack of reliable data particularly in an initial outbreak

The spread of an epidemic in its initial phase can be regarded as transmission under perfect conditions. In its initial phase, the intrinsic epidemiological characteristics of COVID-19, such as reproduction rate, transmission rate, recovery rate, and mortality rate are closer to their true values. However, such data are often insufficient and unreliable. For example, the modeling results in [231] show a wide range of variations due to the lack of reliable data, especially at the beginning of an outbreak.

#### Other issues

Comparing the public data available from different sources also reveals other issues like potential noise, bias and manipulation in some reported case data (e.g., due to their nonuniform statistic standards or manual statistical mistakes), missing values (e.g., unreported on weekends and in the early stage of outbreaks), different categorizations of cases and stages (e.g., some with susceptible and asymptomatic case numbers), misinformation, and lack of information and knowledge about the coronavirus resurgence and mutation.

##### Discussion

While increasing amounts of COVID-19 data are publicly available, they are in fact poor and limited in terms of quality, quantity, capability and capacity to discover deep insights about the hidden nature of COVID-19, the interactions between the virus and host and between them and external factors, and the influence and impact of COVID-19 on their affiliated areas. It is fundamental to acquire substantially larger and better-quality multisource and multimodal internal and external COVID-19 data. Meaningful and insightful COVID-19 modeling can only be robustly conducted and evaluated on such data with the hope of revealing intrinsic knowledge and insights about the disease and assisting effective pandemic control.

### 3.4 COVID-19 Modeling Complexities

The coronavirus and COVID-19 not only present significant challenges to health care, governments, society and the economy but also to the scientific and research communities. The aforementioned challenges have instigated and reshaped the foci of global scientific attention and agendas, and *COVID-19 studies* have emerged as the most important and active research area of recent years. The scientific research is comprehensive, spreading across almost every discipline from epidemiology to psychology and fostering new research areas and topics such as coronavirus epidemiology and genomics. In particular, the growing volume and variety of COVID-19 data form an intangible asset for evidence-based virus and disease understanding, fostering increasingly intensive global research interest and activities in modeling various COVID-19 problems. *COVID-19 modeling* has emerged as a major research direction in COVID-19 studies. It aims to quantitatively understand and characterize the virus and disease characteristics, estimate and predict COVID-19 transmission, and estimate cases, trends, intervention measures and their effects, and estimate their impacts on social, economic, psychological and political aspects. COVID-19 modeling plays an irreplaceable role in almost every aspect of the fight against the COVID-19 pandemic. However, modeling COVID-19 is not a trivial task. Below, we highlight a few challenges in modeling COVID-19.

#### Modeling the COVID-19 characteristics, disease complexities, and data complexities

First, the COVID-19 modeling challenges come from quantifying the diversified and complicated COVID-19 characteristics, disease complexities, and data complexities, discussed in Sections 3.1, 3.2 and 3.3 respectively. The unique characteristics and complexities of the COVID-19 disease and data discussed in Sections 3.2 and 3.3 challenge the existing modeling methods including deep neural learning. Examples are generalized modeling of quality and quantity-limited COVID-19 data from different countries and regions and over evolving time periods [269], robust modeling of short-range, small-size and incomplete-cycle data, and high-capacity modeling of mixture distributions with exponential growth [71], sub-exponential growth [172], discontinuous phase transition [269] and instant changes in case developments. It is challenging to undertake sound, robust, benchmarkable, and practically useful modeling on various characteristics and complexities of the multisource, multimodal and often poor-quality COVID-19 data.

#### Modeling the complexities of open complex COVID-19 ecosystems

The COVID-19 pandemic forms essentially open complex ecosystems when cross-domain and cross-disciplinary aspects are combined. Such ecosystems are associated with significant system complexities [41, 288]. Examples of system complexities are the hidden nature, strong uncertainty, self-organization, dynamics and evolution of the coronavirus, COVID-19 disease, and their continuous developments and mutations; their sophisticated interactions and relations to environments and contexts; the differentiated virus infections of individuals and communities in relation to different ethnic backgrounds; and the significant emergence of consequences and impacts on society and the economy in almost every part of the globe. However, often the related aspects and systems are loosely coupled without tight, consistent identification, connection, or structures as shown in a unified complex ecosystem. It is often difficult to generate a complete and sufficient characterization of the above complexities and intrinsic epidemiological attributes, transmission processes and cause-effect relations in a loosely coupled dynamic COVID ecosystem. The publicly available small, limited, low-quality and loosely-coupled COVID-19 data also do not explicitly support the exploration of COVID ecosystems. COVID-19 modeling has to ‘recover’ and ‘discover’ the hidden, implicit, inconsistent, and loose couplings and relations when multisource, multimodal data are involved.

#### Modeling complex problems with limited-to-no domain knowledge and ground truth

The weak-to-no firm knowledge and ground truth about COVID-19 and its medical confirmation and annotations and the often poor-quality data limit the capacity and richness of the problem hypotheses to be tested and modeled. It is not surprising that rather simple and classic analytical and learning models are predominantly applied by medical and biological scientists to verify specific hypotheses, e.g., various SIR models, time-series regression, and traditional machine learning methods [94, 58, 241]. Most AI and machine learning applications in COVID-19 also only involve simple methods and problem settings. Such simple applications occupy the top-80 keyword-based methods in the 200k WHO-collected references [44]. In contrast, statisticians and computer scientists tend to enforce overparameterized models, over-complicated hypotheses, or over-manipulated data, resulting in highly specific settings and over- or under-fitting results lacking practical applicability [169]. Without ground truth and domain knowledge, unsupervised modeling, multi-aspect modeling, cross-disciplinary modeling, cross-domain modeling, and multi-objective modeling become quite challenging.

#### Aiming for ambitious modeling objectives on low-quality, small COVID-19 data

As discussed in Section 4.2, many COVID-19 problems and objectives are expected to be addressed in modeling COVID-19. However, the complexities of COVID-19 characteristics, diseases and data may significantly limit this potential. Modelers have to carefully define learnable objectives, i.e., what can be learned from the data, acquire the most essential and feasible data, or leverage data poverty using more powerful modeling approaches. For example, when a model is trained on a country’s case numbers, its application to other countries may produce unfair results because of data inequalities. Another example is how to combine multisource but weakly connected data for multi-aspect analyses.

#### Characterizing complicated couplings, relations and interactions in weakly-coupled multi-aspect COVID-19 systems

COVID-19 is affiliated with many personal, social, health, medical, political and other factors, dispersedly reflected in explicitly or implicitly related multisource and cross-domain systems. The COVID-19 pandemic consists of and evolves as a dynamic social-technical process and the co-effects of multi-factor interplay. These multi-aspect factors are coupled strongly or weakly, locally or globally, explicitly or implicitly, subjectively or objectively, statically or dynamically, and essentially or accidentally during the virus and disease formation, development, influence, and evolution. Identifying and characterizing such sophisticated factor couplings and interactions is very challenging as they are not obvious or easily identifiable in their disperse observations. Therefore, modeling the COVID-19 ecosystem requires in-depth and transdisciplinary cooperation and collaborations between computer science, bioinformatics, virology, sociology and other related disciplines. A single factor or aspect cannot disclose the intrinsic and intricate nature of COVID-19, generate a complete picture of the COVID system, and explain the dynamics of this pandemic.

##### Discussion

The COVID-19 characteristics, disease and data complexities introduce significant challenges to their modeling. The challenges lie in the COVID cyber-physical-social-technical ecosystem, the complicated epidemic transmission mechanisms and processes, and the entanglement between epidemic factors (observations) and external objective (e.g., countermeasures) and subjective (e.g., people behavior changes) factors, etc. These determine that COVID-19 modeling goes beyond the transfer, transform and applications of powerful existing models, such as overparameterized deep neural networks, SIR variants, and hierarchical Bayesian networks on the limited, weakly-coupled and small volume of COVID-19 data. Careful modeling mechanisms and designs are essential to address specific COVID-19 characteristics and the complexities of the data and disease, avoid under-/over-fitting, and focus on modeling the complexities in relation to the underlying reality and insight. Complicating models does not necessarily contribute to better or more actionable knowledge and intelligence about the COVID-19 disease and data [28, 40, 258].

The discussion here on the COVID-19 ecosystem complexities informs the selection of the relevant modeling tasks and methods and their review in this paper. Accordingly, in Section 7, we will discuss domain-driven COVID-19 modeling from epidemic and medical and biomedical perspectives, while Section 9 will discuss simulation modeling. Section 8 will review the research on social-domain-related modeling. The mathematical modeling in Section 5 and data-driven modeling in Section 6 will address various technical developments for modeling COVID-19. These further lay the foundation of analyzing the gaps and directions of COVID-19 modeling research in Section 11.

## 4 COVID-19 Modeling Landscape

Here, we provide a brief summary and overview of the research on COVID-19 modeling. We start with an overview of the global research response to COVID-19, then summarize the main objectives of modeling COVID-19 in the literature, and further categorize the research on COVID-19 modeling. These research objectives and categorizations provide structural answers to how the modeling research addresses the aforementioned COVID-19 disease, problems, data and modeling complexities. Lastly, we summarize some global trends of COVID-19 modeling. These together form a high-level research landscape to categorize and connect the comprehensive objectives and techniques of modeling COVID-19.

### 4.1 Global Research on COVID-19

The coronavirus and COVID-19 have inspired increasingly significant research effort globally and in every discipline. There have been about 647k global studies, where about 524k are with full text, on the COVID-19 coronavirus disease collected by WHO between 2019 and 2023^13^. There are 553k articles, 67k preprints, 14k clinical trial registers, and 11k non-conventional documents collected from 124 resources, including Medline (293k), Scopus (53k), Web of Science (42k), ProQuest Central (35k), EMBASE (32k), medRxiv (18k), ICTRP (14k), WHO COVID (14k), and grey literature (11k). The top 10 publishing languages are English (569k), Spanish (14k), Chinese (6.3k), German (6k), Portuguese (5.8k), French (5.3k), Indonesian (4.7k), Russian (3.6k), Japanese (1.6k), and Italian (1.4k). Open access publications play a fundamental role in communicating the results.

The WHO-collected-literature covers a broad list of *subjects*, including 123k on COVID-19, 37k on coronavirus infections, 36k on viral pneumonia, 28k on the pandemic, 24k on SARS-CoV-2, 6k on vaccines, 2k on infection control, 1.8k on mental health, and 1.4 related to social media. The literature cover various *types of study*, including 141k on prognostic study, 119k on risk factors, 106k on random controlled trials, 60k on observational studies, 58k on diagnostic studies, 45k on etiology studies, 45k on qualitative research, 42k on clinical practice guides, and 30k on controlled clinical trials. There are 29k references on reviews and about 9k on systematic reviews.

In our literature review of how global scientists responded to COVID-19^14^ [44], we collected 346k references from major databases including Web of Science, Scopus, PubMed and PMC, the WHO collection on global research on COVID-19, and ResearchGate, as well as medRxiv and arXiv between 1 Jan 2020 and 9 Mar 2022, where about 51k are related to modeling COVID-19. About 176 first-authored countries and regions have contributed to COVID-19 research. The literature involves almost every discipline, from medical and biological science to computer science, engineering, economics, environment, and policy. The broad disciplines related to medical and health sciences contributed about 186k, in comparison to 14k by computer scientists, and 32k by social scientists. There has been a significant monthly increase in the number of publications over 2021 to 2022 in comparison with that in 2020.

In our collection [44], the top-10 first-authored countries are the US, China, Italy, the UK, India, Spain, Canada, Germany, Brazil, and France. Fig. 3 shows the publication distribution of the 175 first-authored countries.

**Fig. 3.**
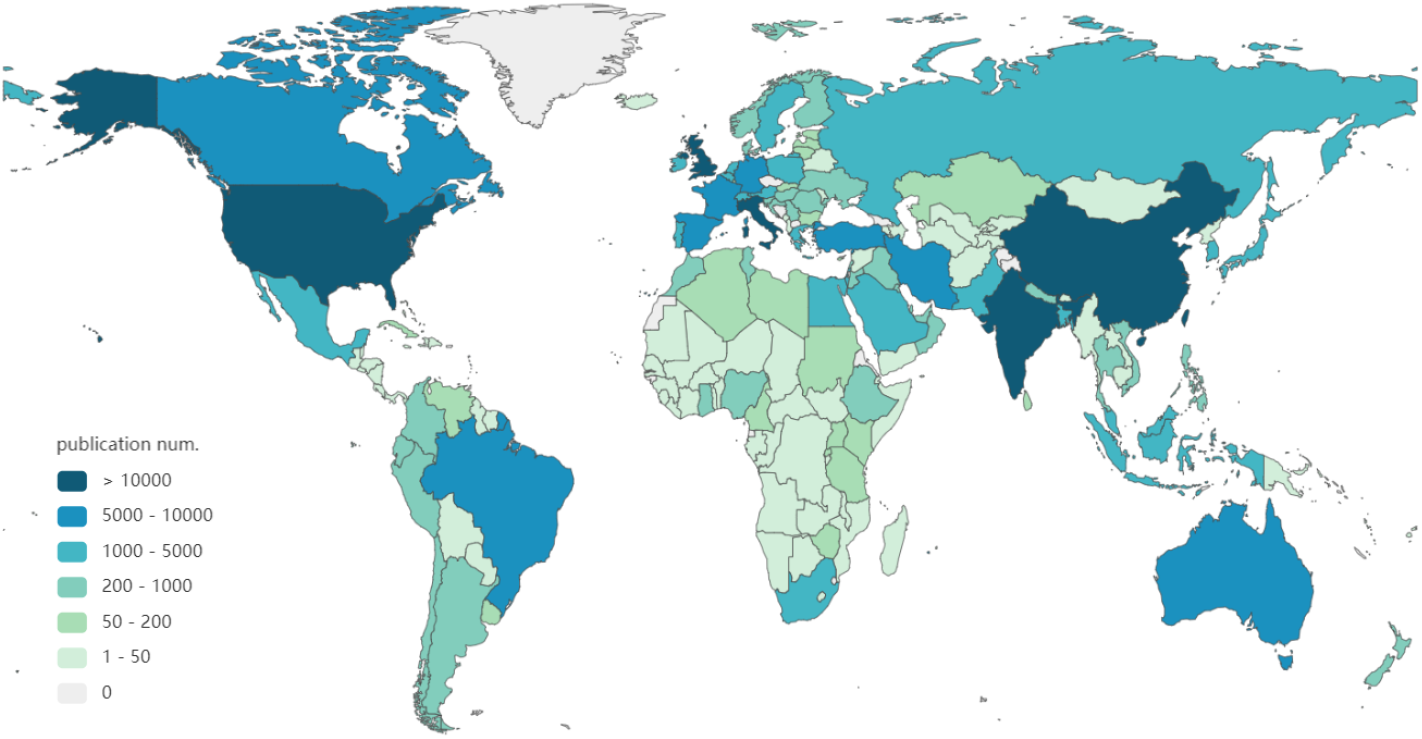
Global first-authored publication distribution

Further, the top keywords of global concern include pandemic (129k), patient (105k), infection (73k), risk (55k), treatment (43k), hospital (36k), measure (34k), symptom (32k), management (28k), and development (28k). Fig. 4 shows the word cloud of the top-200 words appearing in all publications.

**Fig. 4.**
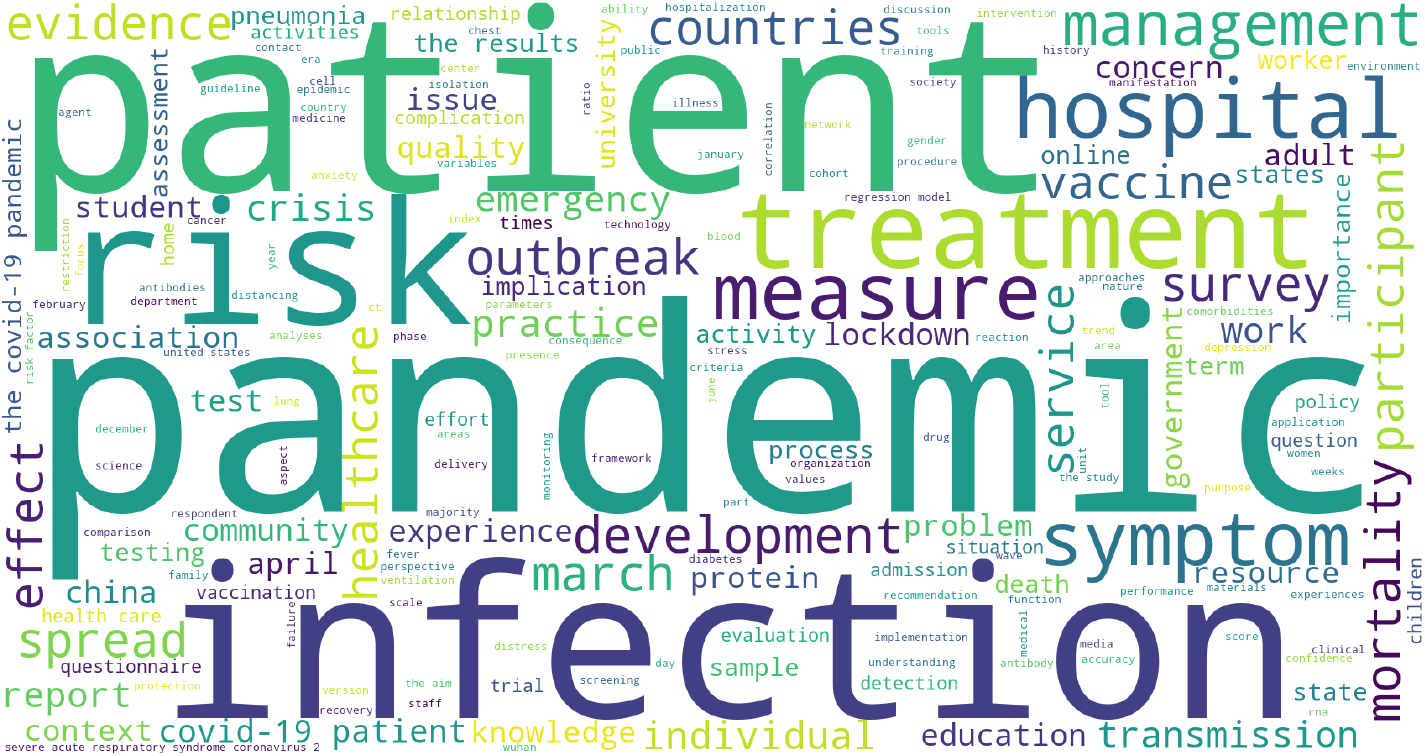
Global top-200 word cloud of all publications

**Fig. 5.**
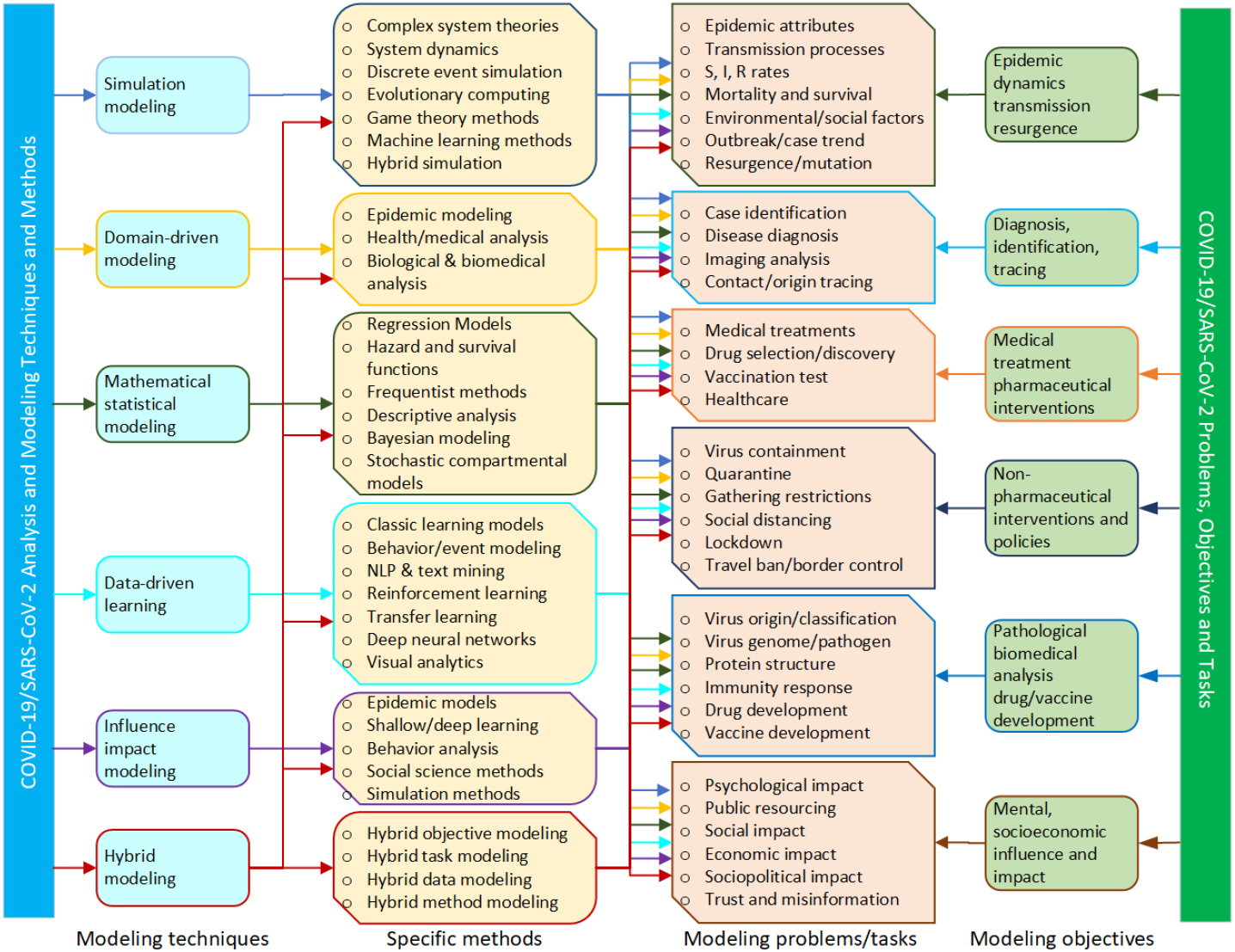
The transdisciplinary research landscape of COVID-19 modeling: From right: coronavirus problems and modeling objectives, to the left: modeling techniques and their representative methods.

### 4.2 Objectives of COVID-19 Modeling

Here, we summarize the main problems and objectives of modeling COVID-19 in the literature. The analysis of the WHO-collected literature [44] gives us a clear indication of the top problem terms in over 346k global references and those 51k modeling-focused ones. The keywords appearing most frequently in the 346k references are pandemic, patient, infection, risk, treatment, hospital, measure, symptom, management, development, outbreak, mortality, spread, healthcare, vaccine, transmission, crisis, and lockdown. The top-ranked problem keywords appearing in the 51k modeling publications include mental health, second wave, lockdown, vaccine, anxiety, risk factor, vaccination, X-ray, public health, depression, treatment, social media, and social distancing. In addition, the top keywords appearing in the WHO collected literature also indicate the key foci on infection, viral, pandemics, SARS-CoV-2, vaccine, telemedicine, antibody, disease control, infection control, mental health, quarantine, public health, healthcare, respiratory distress syndrome, hospitalization, testing, lung, social media, and vaccination.

The above keywords inform the key problems and objectives associated with modeling COVID-19. By further clustering the problems appearing in modeling publications, Table 1 categorizes and illustrates their representative modeling factors, modeling methods, and references. Below, we further elaborate on a few objectives widely explored in the literature on modeling COVID-19.

#### Characterizing and predicting COVID-19 epidemic dynamics and transmission

An important challenge is to understand the COVID-19 epidemic mechanisms, transmission processes, and their dynamics. It is also important to infer their epidemiological attributes and to understand how the coronavirus spreads spatially, temporally, and socially [35, 34]. In this regard, intensive COVID-19 modeling research focuses on exploring the source and spectrum of the COVID-19 infection, identifying clinical and epidemiological characteristics, tracking transmission routes, and forecasting case development trends and the peak values of infected cases and disease transmission [35, 34, 22, 50, 293, 294, 95, 125, 184]. The related work aim to understand the characteristics and dynamics of the coronavirus and COVID-19 disease to inform disease precaution, characterize the epidemiological attributes of coronavirus and COVID-19 and their resulting infections, mortality and patient statistics, quantifying the influence of virus containment and mitigation campaigns on epidemic dynamics and virus transmission, and measuring the influence of epidemic and infections on medical resource planning, etc. Typical modeling methods include mathematical and statistical models such as linear and nonlinear regression models [22, 321, 184], compartmental models including those incorporated with statistical settings and customization such as time and age dependence, SIR with Markov chain Monte Carlo [321], simulation methods [52, 125], and network modeling [95, 167].

#### Modeling the COVID-19 resurgence and mutation

SARS-CoV-2 has shown strong and uncertain dynamics, evolution and mutations with various strains emerging over time, including the recent increasingly infectious Omicron BA.5 [134]. Accordingly, most countries and regions have experienced multiple COVID-19 waves and resurgences. Both mutations and resurgences are highly uncertain, although they seem to become more transmissible and infectious [160] but less fatal [83, 99]. The WHO-identified five major variants of concern which show higher transmissibility, contagion and complexities^15 16^ [100, 101]. However, as our current understanding of the resurgence and mutation is still very limited, COVID-19 may become another epidemic disease which stays with humans for a long time. More research is expected to quantify the resurgence conditions, control potential resurgences after lifting certain restrictions and reactivating businesses and activities [163, 206], distinguish the characteristics and containment measures between waves and mutations [83, 99], predict future resurgences and mutations [101, 134], and prepare for their responsive countermeasures [12]. Various modeling methods are applied, including mathematical and statistical models such as regression models, ordinary differential equation and Bayesian inference [163], compartmental models [36, 206], simulation models, and various AI techniques including NLP and machine learning models [218].

#### Modeling the COVID-19 diagnosis, infection identification, and contact tracing

COVID-19 and its virus mutations present strong transmission and reproduction rates, high contagion, sophisticated transmission routes, and unexpected resurgences and cluster spreads. These make their diagnosis, infection identification and contact tracing difficult. It thus becomes crucial to immediately identify and confirm exposed cases and trace their origins and contacts to proactively implement quarantine measures and contain their potential spread and outbreak [189]. This is particularly important during the varying incubation periods and for those strains that are associated with high presymtomatic, asymptomatic to mildly symptomatic yet highly contagious infections. In addition to chemical and clinical approaches, identifying COVID-19 by analyzing the biomedical images, genomic sequences, symptoms, social activities, mobility, and media communications is also essential [263].

#### Modeling the efficacy of medical treatment and pharmaceutical interventions

The general practices of timely and proper COVID-19 medical treatments, drug selection and pharmaceutical measures, ICU and ventilation play a fundamental role in the mitigation of severe symptoms, and a reduction in the mortality rate of both the original and increasingly mutating virus strains. However, the lack of best practices and standardized protocols and specifications of medical and pharmaceutical treatments on the increasingly evolving virus variants, unexpected local and home clusters, and outbreaks in rural and disadvantaged areas affects the globalized standard treatment and control of the virus. Medical treatment may also be affected by the insufficient understanding of the couplings between virus infection and patient’s demographics and ethnic context. The wide dispersal of online misinformation of drug use and vaccination may also contribute to the global imbalance in treating COVID-19. Research is required to select and discover suitable drugs, analyze the drug-target interactions, and best match the patient’s diagnosis and ethnic contexts with their suitable medical treatments to control critical conditions and mortality and prioritize public health resources in a timely manner. [320, 293, 20].

#### Modeling the effect and impact of non-pharmaceutical intervention and policies

Various NPIs, such as travel bans, border control, business and school shutdowns, public and private gathering restrictions, mask-wearing, and social distancing, are often implemented to control the outbreak of COVID-19. NPIs are often combined in different ways and at different enforcement levels to contain outbreaks. Their easing and removal usually follows different policies, procedures and timeframes. Such customized NPI applications result in inconsistencies across countries and regions and different effects and impacts which are difficult to benchmark and compare. Limited research results have been reported to verify the effects and impacts of these control measures in different conditions and combinations on containing the virus spread and case number development, the balance between enforcement levels and containment results, and the response sensitivity of the restrictions in relation to the population’s ethnic context [257, 71, 30]. Robust results from investigations of these aspects can greatly complement medical and health treatments and inform public health policy-making on medication, business and societal management during COVID-19 outbreaks.

#### Pathological and biomedical analyses for drug and vaccine development

To develop drugs and vaccines for COVID-19, pathological and biomedical analyses analyze pathological test results, gene sequences, protein sequences, physical and chemical properties of SARS-CoV-2, the drug-target interaction and mapping, and the effect of candidate drugs and vaccines on people with varied backgrounds and conditions. Building on domain knowledge and techniques such as virology, pathogenesis, genomics and proteomics, it is essential to conduct domain- and data-driven analyses of drug-target interactions, identify the COVID-19 sensitive genomic and protein structures, and select and develop COVID-19-matched drugs and vaccines [132, 143]. It is also important to diagnose and identify the interactions and mapping between mutated virus strains and effective drugs and vaccines. [20, 227]. More research is needed on COVID-19 immunity responses, drug and vaccine development, and mutation intervention. In addition, limited results are available on the threshold and effects of various COVID-19 vaccines on different profiles of people and the resulting effect of herd immunity [271].

#### Modeling COVID-19 influence and impact

While the COVID-19 pandemic has changed the world and has had a significant and overwhelming influence on almost all aspects of our lives, society and the economy, quantifying its influence and impact has been insufficiently studied. Various objectives may be related to modeling the negative COVID-19 influence and impact, including (1) the economic impact on growth and restructuring [291]; (2) the social impact on people’s stress, psychology, emotions, behavior and mobility [207, 299]; and (3) the transformation of business processes and organizations, manufacturing, transport, logistics, and globalization [272, 238]. In addition, the COVID-19 pandemic also triggers various transformations and new requirements, for example, (1) enhancing the wellbeing and resilience of individuals, families and society and work-life balance [221]; (2) digitizing, virtualizing and transforming work, study, collaboration, entertainment, and shopping [250]; (3) restructuring the supply-demand relations and supply chains for better and immediate availability and to satisfy emerging demand [72]; (4) promoting research and innovation on intervening in global black-swan disasters such as on public health resources, transport and their impacts [316]; and (5) enhancing the public trust and developments in pandemic science, medicine, vaccination and hygiene [215]. Other impact modeling tasks include analyzing the relations between the COVID-19 containment effect and the socioeconomic level (e.g., income level particularly in relation to lower-income and disadvantaged groups), healthcare capacity and quality, government crisis management capabilities, citizen-government cooperation, and public health and hygiene habits.

### 4.3 Categorization of COVID-19 Modeling

Many techniques have been involved in addressing the objectives detailed in Section 4.2. The review of both the WHO-collected COVID-19 global literature on coronavirus disease and our analysis of global references on COVID-19 modeling [44] has resulted in the following categorization observations.

First, the COVID-19 modeling literature shows strong research features: (1) involving multi-disciplinary techniques, including mathematics and statistics, epidemiology, broad AI and data science including shallow and deep learning, and social science; (2) applying epidemiological methods to explore the COVID-19 pandemic problems; (3) involving domain-, model- and data-driven approaches from various families of disciplines and research areas; (4) demonstrating case studies and hypothesis tests with the results of particular methods highlighting specific modeling settings, scenarios, or data.

Second, the main COVID-19 modeling techniques consist of epidemiological modeling, mathematical and statistical modeling, artificial intelligence and data science, and simulation modeling. They have been applied to understanding, characterizing, simulating, analyzing, and predicting various COVID-19 problems and issues. Accordingly, we categorize the research landscape of COVID-19 modeling into six families: domain-driven modeling, mathematical and statistical modeling, data-driven learning, influence and impact modeling, simulation modeling, and hybrid methods in this review.

Further, Fig. 5 presents a research landscape of COVID-19 modeling. It summarizes the transdisciplinary research on modeling COVID-19 into six categories of modeling techniques and illustrates their representative modeling methods respectively. It also lists the main problems and their further decomposition commonly used in modeling COVID-19 linking to the modeling objectives in Section 4.2. The landscape further connects the modeling problems to their respective modeling techniques. These form an overall research map of the research on modeling COVID-19.

Lastly, below, we briefly summarize the main modeling tasks and methods associated with each category of the modeling techniques in Fig. 5.

- *COVID-19 mathematical and statistical modeling*: developing and applying mathematical and statistical models to estimate COVID-19 transmission processes, symptom identification, disease diagnosis and treatment, sentiment analysis, misinformation analysis, and resurgence and mutation, etc. Models include time-series analysis such as regression models, and hazard and survival functions, and statistical models such as descriptive analytics, statistical processes, latent factor models, temporal hierarchical Bayesian models, and stochastic compartmental models.
- *COVID-19 data-driven learning*: developing and applying data-driven analytics and learning methods to characterize, represent, classify, and predict COVID-19 problems on COVID-19 data. Typical methods include classic (e.g., tree models such as random forests and decision trees, kernel methods such as support vector machines (SVMs), NLP and text analysis, and classic reinforcement learning) and deep (e.g., deep neural networks, transfer learning, deep reinforcement learning, and variational deep neural models) analytics and learning methods. Problems include case development, mortality and survival forecasting, medical imaging analysis, NPI effect estimation, and genomic analysis.
- *COVID-19 domain-driven modeling*: developing and applying domain-specific models by incorporating domain knowledge and factors for quantifying COVID-19. Examples are epidemiological compartmental models to characterize the COVID-19 epidemic transmission processes, dynamics, transmission and risk; social science methods for estimating the influence of external factors on COVID-19 epidemics, resurgence and mutation; and medical, pathological and biomedical analyses for infection diagnosis, case identification, patient risk and prognosis analysis, medical imaging-based diagnosis, pathological and treatment analysis, and drug development.
- *COVID-19 simulation modeling*: developing and applying simulation models to simulate the COVID-19 epidemic evolution and the effect of interventions and policies on the COVID-19 epidemic. Examples are theories of complex systems, agent-based simulation, discrete event analysis, evolutionary learning, game theories, and Monte Carlo simulation.
- *COVID-19 influence and impact modeling*: developing and applying methods to estimate and forecast the influence and impact of SARS-COV-2 variations and COVID-19 diseases, as well as their interventions, treatments and vaccination. Areas influenced or impacted by these aspects include epidemic transmission dynamics, virus containment, disease treatment, human psychological health, behaviors and mobility, public resourcing (such as healthcare systems), social systems, and the economy.
- *COVID-19 hybrid modeling*: hybridizing and ensembling multiple models to tackle multiple coronavirus problems and objectives, multiple tasks, or multisource data, when those individual objectives, tasks, data or models cannot be better addressed by single aspects or approaches.

It is worth mentioning that each of the above modeling techniques and their corresponding methods may be applicable to address different and multiple coronavirus problems and modeling objectives, as shown in Fig. 5. In Sections 5 to 10, we review, categorize and comment on the progress of these six categories of COVID-19 modeling. Our foci are not on their specific references, rather on (1) introducing their typical representative techniques and methods, and (2) aligning the techniques with their typical applications in modeling diverse COVID-19 problems and issues. Each illustrated modeling technique is not from a specific reference, instead it aims for a representative approach usually integrating multiple designs or addressing various issues.

### 4.4 Global Trends in COVID-19 Modeling

Here, we present a snapshot of the global trends in modeling COVID-19. As reported in [44], of the 346k references published on COVID-19, over 51k are on COVID-19 modeling. In those modeling publications classifiable to disciplines, about 6k publications appeared in computer science venues. In contrast, over 21k publications on modeling appeared in medical science venues.

In computer science, AI and data science techniques are mostly applied. In particular, data-driven discovery [266, 42, 79], including both classic and deep analytical and machine learning methods are mostly applied. *COVID-19 data science* plays a major role in COVID-19 modeling, aiming to discover valuable knowledge and insights from various kinds of publicly available data, including daily cases, texts, biomedical images, mobility, and environmental factors. As further illustrated in the following sections, almost every modeling technique has been applied to COVID-19 in some way. For example, classic epidemic models were tailored for COVID-19 to model its macroscopic transmission and predict the trends of the spread of the virus. Generative models with Bayesian hierarchical structures were applied to capture the effects of NPIs. Deep natural language processing approaches were adopted to understand the growth, reality and spread of COVID-19 and people’s reactions based on the textual data from social media. Modeling also helps to understand and characterize every aspect of coronavirus and COVID-19 from its epidemiological characteristics to the underlying genomic reactions, virus mutations, and drug and vaccine development.

Below, we illustrate the trends of COVID-19 modeling in all modeling publications and in computer science and medical science respectively^17^. First, Fig. 6 shows the word cloud of the top-200 modeling keywords appearing in all modeling publications. The results show that the modeling methods are diversified, covering classic mathematical and statistical methods, epidemic modeling methods, simulation, and classic machine learning methods. Regression models, machine learning, simulation, linear regression, multivariate statistics, artificial intelligence, logistic regression, statistical models, deep learning, and CNNs rank in the top keywords in the literature on modeling COVID-19. This shows that classic regression methods dominate the publications in modeling COVID-19, followed by statistical models and machine learning. Deep learning has a strong presence in modeling COVID-19, in particular, using the most fundamental network CNNs in comparison with shallow machine learning models.

**Fig. 6.**
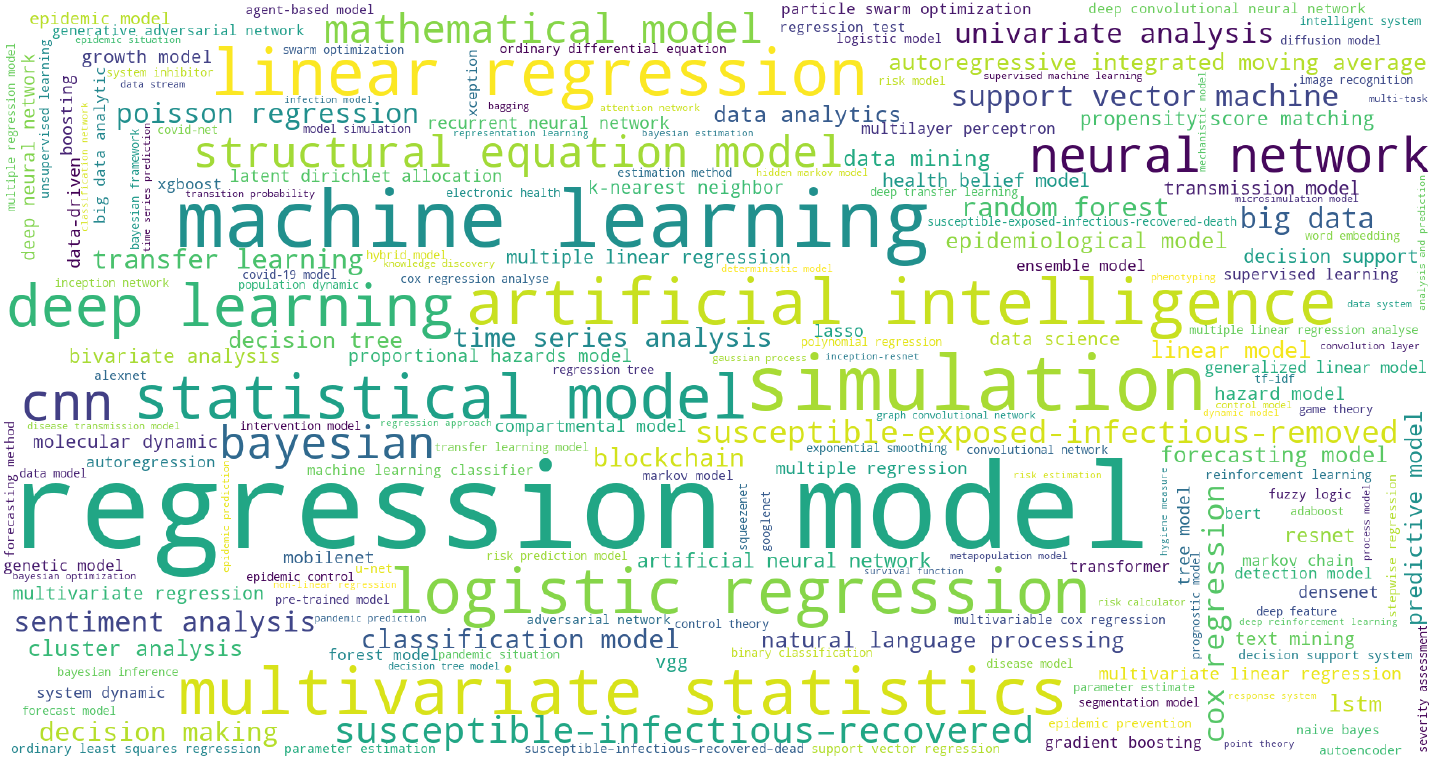
Keyword cloud of all modeling publications

In contrast, Fig. 7 shows the word cloud of the top-200 modeling words appearing in modeling publications which belong to the computer science category. The modeling publications from the computer science discipline mainly apply machine learning, deep learning, and AI methods, although mathematical models also appear very frequently in the publications. Fig. 8 shows the word cloud of the top-200 modeling keywords appearing in modeling publications which belong to the medical science category. Although the top-200 keywords shown in Fig. 8 largely overlap with those in the overall and computer science-based publications, publications related to medical science seem to favor classic mathematical and statistical methods than modern methods such as deep learning and machine learning, and their frequencies are also much higher than those in computer science.

**Fig. 7.**
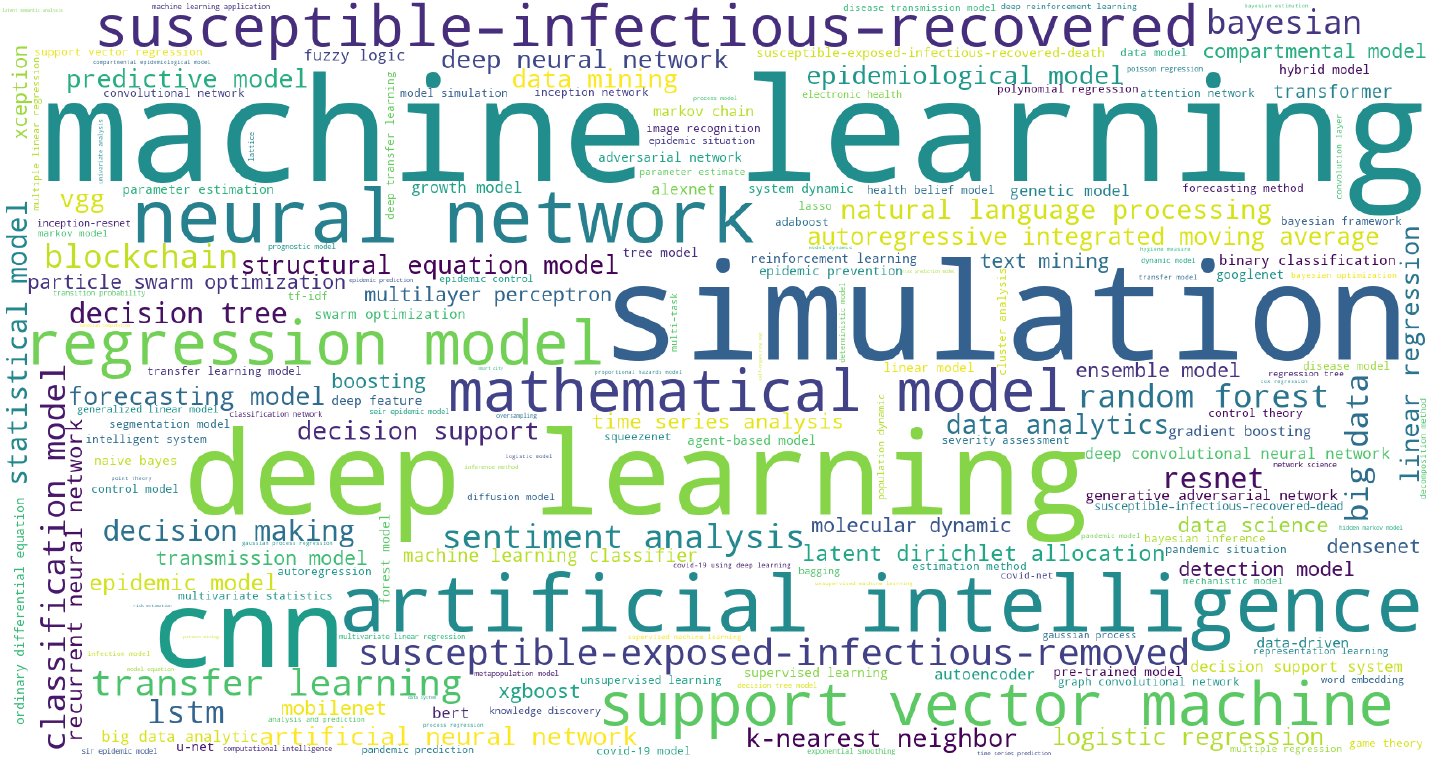
Keyword cloud of modeling publications in computer science

**Fig. 8.**
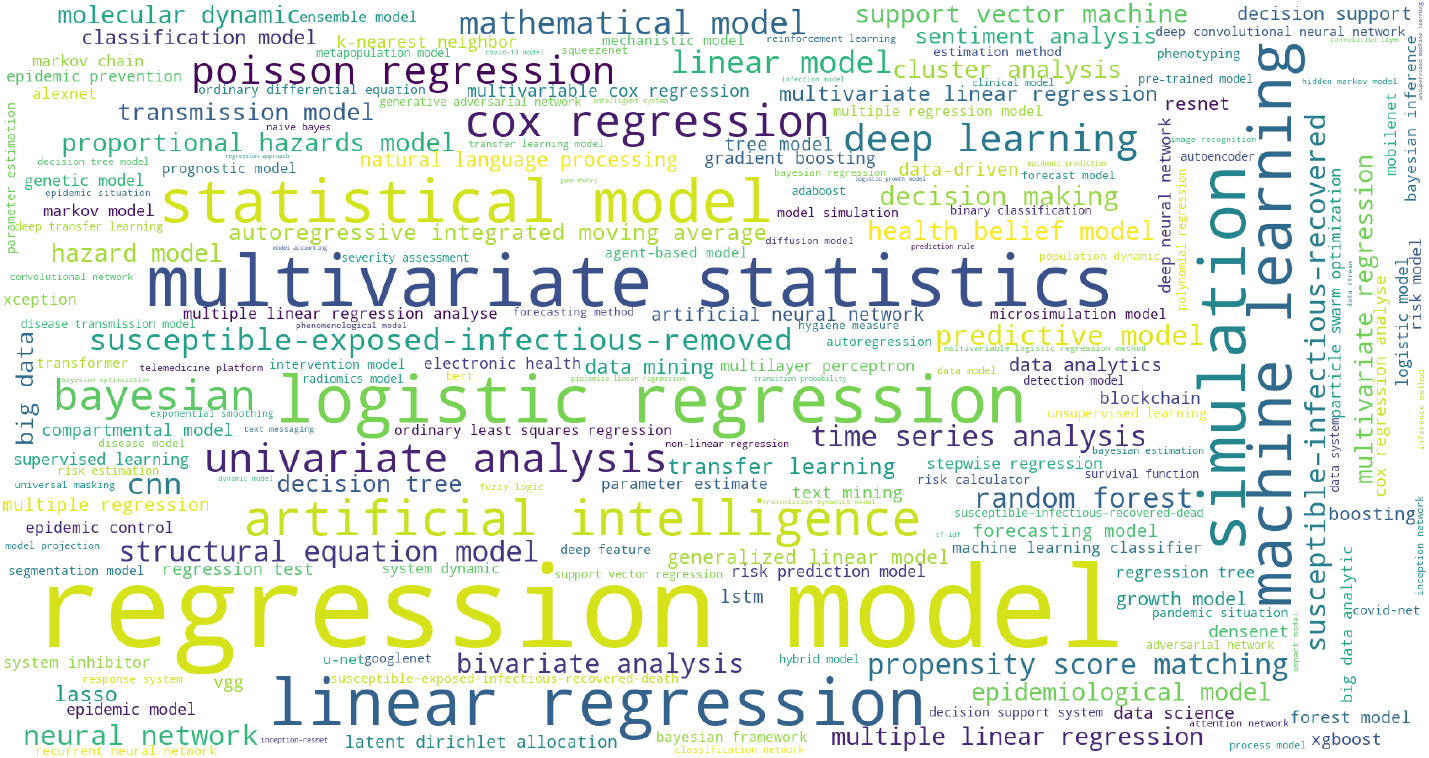
Keyword cloud of modeling publications in medical science

## 5 COVID-19 Mathematical Modeling

Mathematical and statistical models have been used extensively to estimate and predict the transmission dynamics of coronavirus and COVID-19 and to reveal the truth of the epidemic in a formal and quantitative manner. Accurate COVID-19 mathematical modeling is indispensable for quantifying COVID-19 systems and its epidemic forecasting and decision making. Here, we review two sets of mathematical methods: time-series analysis and statistical modeling, which are the most commonly applied in COVID-19 modeling references across the body of literature.

### 5.1 COVID-19 Time-series Analysis

Here, we first briefly introduce the time-series models commonly applied in COVID-19 modeling. Then, we summarize the related work on applying such time-series models to quantify COVID-19 problems.

#### 5.1.1 Time-series models

We here briefly introduce two typical time-series models which have been predominantly customized for modeling COVID-19: regression models, and hazard and survival functions.

##### Typical regression models

Typical regression models such as logistic regression and auto-regressive integrated moving average (ARIMA) variants have been widely used in epidemic and COVID-19 modeling. *Logistic growth models* estimate the number of COVID-19 infected cases [283]. Assume a COVID-19 outbreak is associated with a population of *S*_*t*_ with the infection rate *β* at time *t*, the growth scale of infected case number *I*_*t*_ can be modeled by

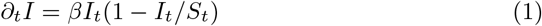

over time *t*. Accordingly, with historical COVID-19 cases at the bursting point at time period *T*, an S-shaped curve will be derived to describe and forecast the growth distributions of COVID-19 infections and the infected peak number by adjusting the constant rate *β* (assuming it is constant; in the case where the infection rate is evolving, we would have *β*_*t*_).

Although such logistic models can be widely seen in the COVID-19 modeling literature, they are weak or even incapable of modeling the complex transmission states and dynamics of a COVID-19 outbreak. Many other challenges, such as the nonstationary characteristics discussed in Section 3.3 and the disease complexities in Section 3.2, need to be better modeled by more sophisticated regression models or other advanced methods.

Alternatively, *ARIMA* and its variants have been widely applied to model the temporal movement of COVID-19 case numbers with more flexibility than the logistic models. One example of applying ARIMA for modeling COVID-19 is as follows. Given the number *I*_*t*_ of infected cases at time *t*, it can be modeled by model *ARIMA*(*p, d, q*) which factorizes the number *I*_*t*_ into consecutive past numbers {*I*_*t−p*_, …} with errors *ε*:

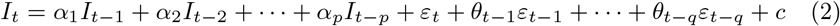

over the number of time lags (order) of autoregression *p*, the order of moving average *q*, the degree of differencing *d*, and time *t* with a constant *c. I*_*t−p*_ refers to the infected cases at time *t*−*p* with weight *α*_*p*_, *ε*_*t−q*_ refers to the error between *I*_*t−q*_ and *I*_*t−q−* 1_ with weight *θ*_*t−q*_. Adjusting parameters like *p, d* and *q* can simulate and capture some of the time-series characteristics. Examples are to model the process and trend of the COVID-19 infection series by *p* and *q*, seasonality by *d*, and volatile movement by the distribution of error terms. Similarly, ARIMA models can be used to simulate and forecast the evolving number of recoveries and deaths in relation to COVID-19.

In addition, ARIMA and its variants can be integrated into other modeling methods to characterize multiple aspects of the COVID-19 time-series. For example, the wavelet decomposition of frequency-based nonstationary factors can model the oscillatory error terms in the ARIMA-based modeling of COVID-19 infected cases [48]). Another typical example is to combine the decision tree model with regression to form a regression tree and identify mortality-sensitive COVID-19 factors [49].

##### Hazard and survival functions

Hazard functions and survival functions have been used to model the mortality and survival (recovery) rates of patients using time-to-event analysis. A hazard function models the mortality probability *h*(*t*|**x**) of a COVID-19 patient with factor vector **x** (∈ ℛ^*d*^) of dying at discrete time *t* :

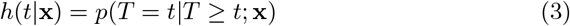

On the contrary, the survival function models the probability *S*(*t*|**x**) of surviving until time *t* :

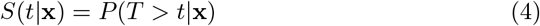

In discrete time, 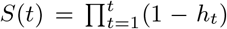 where *h*_*t*_ is the mortality probability at time *t*. In continuous time, *S*(*t*|**x**) = 1 − *F* (*t*|**x**) where *F* (·) is the cumulative distribution function until time *t*. For covariates **x** with their relations represented by either a linear or nonlinear function *f* (·; *δ*) with parameters *δ*, the mortality rate of COVID-19 can be modeled by a Cox proportional hazard model [224]:

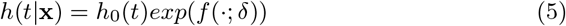

where *h*_0_(*t*) is the baseline hazard function, function *f* (·; *δ*) can be implemented by a linear function such as a linear transform or a nonlinear function such as a deep convolutional network. For example, in [237], *f* (·; *δ*) was implemented as a shallow neural network with a leaky rectified linear unit-based activation of the input and then another tangent transformation. In the case of time-varying covariates **x**_*t*_, the above hazard, survival and transform functions should be time sensitive as well.

##### Other time-series models

In addition, other commonly used time-series models include linear regression models such as ARIMA and GARCH [247, 253], logistic growth regression [283], Cox regression [237], multivariate and polynomial regression [6, 55, 105], generalized linear model and visual analysis [185], support vector regression (SVR) [102, 229], regression trees [49], hazard and survival functions [237], and more modern LSTM networks [119]. In addition, temporal interpolation methods such as best fit cubic, exponential decay and Lagrange interpolation, spatial interpolation methods such as inverse distance weighting, smoothing methods such as moving average, and spatio-temporal interpolation have also been applied to fit and forecast COVID-19 case time-series. Typically, the performance of mathematical COVID-19 modeling is measured by metrics such as mean absolute error (MAE), root mean square error (RMSE), the improvement percentage index (IP), and symmetric mean absolute percentage error (sMAPE) in terms of certain levels of confidence intervals.

#### 5.1.2 COVID-19 time-series modeling

Time-series analysis comprises the most commonly used methods (about 13k references appearing in the top-10 most commonly used modeling methods over the 30k modeling publications) for COVID-19 modeling [44]. Regression models, linear regression, and logistic regression are most commonly applied in COVID-19 modeling. Many linear and nonlinear, univariate, bivariate and multivariate analysis methods have been intensively applied for the regression and trend forecasting of new, susceptible, infectious, recovered and death case numbers of COVID-19. Below, we illustrate a few tasks commonly addressed in the literature: COVID-19 epidemic distributions, case number and trend forecasting, COVID-19 factor and risk analysis, and the correlation analysis between COVID-19 epidemic dynamics and external factors.

##### Forecasting COVID-19 case number, trend and epidemic distributions

Regression-centered time-series analysis has been widely applied to forecast case number developments and trends. For example, Singh et al. [247] applied ARIMA to predict the COVID-19 spread trajectories for the top 15 countries with confirmed cases and concluded that ARIMA with a weight to adjust the past case numbers and the errors has the ability to correct model prediction and is better than regression and exponential models. However, ARIMA lacks flexible support for the volatility and in-between changes during the prediction periods [247]. Gupal et al. [105] adopted polynomial regression to predict the number of confirmed cases in India. Almeshal et al. [9] utilized logistic growth regression to fit the actual infected cases and the growth of infections per day. Wang et al. [283] modeled the cap value of the epidemic trend of COVID-19 case data using a logistic model. With the cap value, they derived the epidemic curve by adapting time-series prediction. To find the best regression model for case forecasting, Ribeiro et al. [229] explored and compared the predictive capacity of the most widely used regression models including ARIMA, cubist regression (CUBIST), random forest, ridge regression, SVR, and stacking-ensemble learning models. They concluded that SVR and stacking ensemble are the most suitable for short-term COVID-19 case forecasting in Brazil. In addition, linear regression with the Shannon diversity index and Lloyd’s index are applied to analyze the relations between the meta-population crowdedness in city and rural areas and the epidemic length and attack rate [225].

##### COVID-19-specific factor and risk analysis

Time-series analysis is also used to (1) analyze the influence of specific and contextual factors on COVID-19 infections and COVID-19 epidemic developments, including infection, transmission, outbreak, hospitalization, and COVID-19 survival, mortality and recovery; and (2) analyze the influence and impact of external and contextual factors of the COVID-19 outbreak on the population, health, society and the economy, as well as case developments and containment. For example, to investigate the potential risk factors associated with fatal outcomes from COVID-19, Schwab et al. [237] presented an early warning system assessing COVID-19-related mortality risk with a variation of the Cox proportional hazard regression model. Chen et al. [57] adapted the Cox regression model to analyze the clinical features and laboratory findings of hospitalized patients. Charkraborty et al. [49] designed the wavelet transform optimal regression tree (RT) model, which combines various factors including case estimates, epidemiological characteristics and healthcare facilities to assess the risk of COVID-19. The advantage of RT is that it has a built-in variable selection mechanism from high dimensional variable space and can model arbitrary decision boundaries.

##### Correlation analysis between COVID-19 epidemic dynamics and external factors

Much research has been conducted on analyzing the relationships between COVID-19 transmission and dynamics and external and contextual factors. For example, Cox proportional hazard regression models are used to analyze high-risk sociodemographic factors such as gender, individual income, education level, and marital status that may be associated with a patient’s death [76]. Logistic regression models are applied to analyze the relations between COVID-19 (or SARI with unknown aetiology) and socioeconomic status (per-capita income) [70]. To reveal the impact of meteorological factors, Chen et al. [55] examined the relationships between meteorological variables (i.e., temperature, humidity, wind speed, and visibility) and the severity of the outbreak indicated by the confirmed case numbers using the polynomial regression method. Liu et al. [157] fit the generalized linear models (GLM) with a negative binomial distribution to estimate the city-specific effects of meteorological factors on confirmed case counts. In [216], Loess regression does not show an obvious relation between the COVID-19 reproduction number, weather factors (humidity and temperature), and human mobility. Lastly, linear models including linear regression, Lasso regression, ridge regression, elastic net, least angle regression, Lasso least angle regression, orthogonal matching pursuit, Bayesian ridge, automatic relevance determination, passive aggressive regressor, random sample consensus, TheilSen regressor, and Huber regressor are applied to analyze the potential influence of weather conditions on the spread of coronavirus [173].

###### Discussion

Time-series methods excel at characterizing sequential transmission processes and temporal case movements and trends of short and small COVID19 case data. However, they lack the capability to involve other multi-source, multimodal factors and heterogeneous external factors. Their capacity in disclosing deep insights into why case numbers evolve in a certain way and how to intervene in the infection, treatment and recovery is also limited.

### 5.2 COVID-19 Statistical Modeling

Statistical learning, in particular Bayesian modeling, plays a critical role in stochastic epidemic and infectious disease modeling [29]. Typically, generative stochastic processes are assumed to model epidemic contagion for epidemic modeling [10, 202]. In contrast to compartmental models (Section 7.1), statistical models incorporate prior knowledge about an epidemic disease. Their results have confidence levels corresponding to distinct assumptions (i.e., possible mitigation strategies), which better interpret and more flexibly model COVID-19 complexities. Below, we summarize typical statistical models and their applications in COVID-19 statistical modeling.

#### 5.2.1 Statistical models

Typical statistical methods include descriptive analytics, Bayesian hierarchical models, probabilistic compartmental models, and probabilistic deep learning encompassing shallow to deep analytics. Below, we introduce some common statistical settings and their corresponding statistical models in both frequentist and Bayesian families that are often applied in COVID-19 statistical modeling.

##### Hierarchical Bayesian modelling

Taking a stochastic (vs. deterministic) state transition assumption, various statistical processes can be assumed to simulate and estimate the state-specific counts (case numbers) and the probability of state transitions (e.g., between infections and deaths) during the COVID-19 spread. The stochastic processes and states (e.g., its infection and mortality) of a COVID-19 outbreak are influenced by various explicit and latent factors. Examples of explicit (observable) factors include a person’s demographics (e.g., age and race), health conditions (e.g., disease history and hygienic conditions), social activities (e.g., working environment and social contacts), and containment actions (e.g., quarantined or not) taken on each person. Latent factors may include a person’s psychological attitude toward cooperation (or conflict) with containment, health resilience strength to coronavirus, and the containment influence on the outcome (e.g., infected or deceased).

Fig. 9(a) illustrates a general graphical model of the temporal hierarchical Bayesian modeling of COVID-19 case numbers for estimation and forecasting. The reported case number *Y*_*t*_ (e.g., death toll or infected cases) at time *t* can be estimated by 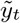, which is inferred from the documented (declared) infections 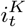 and removed (e.g., recovered and deceased) rate *κ*_*t*_. The documented infection number 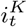 is inferred from infected population *i*_*t*_ and test rate *ρ*_*t*_. *i*_*t*_ is inferred from exposed population *e*_*t*_ and infection rate *β*_*t*_, *e*_*t*_ is determined by its exposed rate *E*_*t*_. Further, we assume the removed rate *κ*_*t*_ is influenced by various medical treatments *α*, determined by auxiliary variables including socioeconomic condition *λ*_2_, the treatment effectiveness *ψ*, and the public health quality *ω*. Infection rate *β*_*t*_ is determined by NPIs *ζ*, which is further influenced by the NPI execution rate *τ* and the socioeconomic factor *λ*_1_. The priors of the corresponding parameters are *a, b, c, d*_1_, *d*_2_, *f, g, h* and *j*, which may follow specific assumptions.

**Fig. 9.**
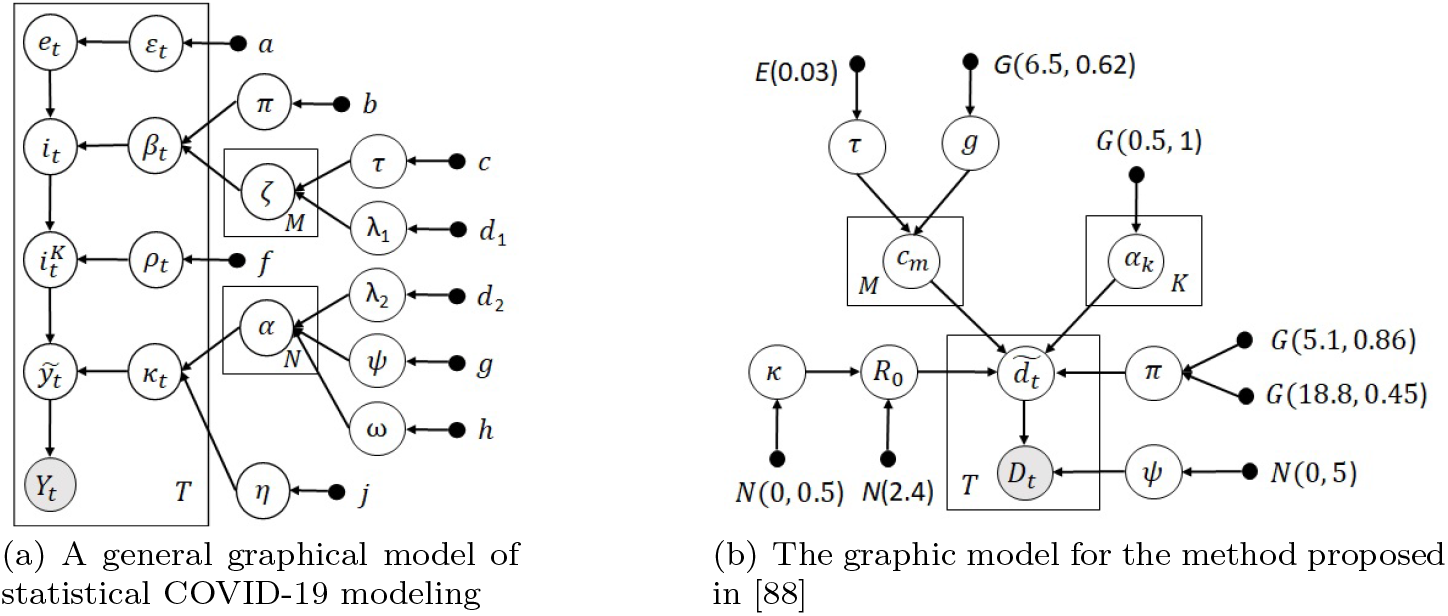
A general and a specific hierarchical Bayesian model for COVID-19.

For the statistical settings and hypotheses, typical statistical distributions of COVID-19 state-specific counts are applied to (1) *infection modeling*, e.g., by assuming a Bernoulli process (*B*(*n, ε*) with the probability *ε* of exposure to infections over *n* contacts) and then a Poisson process at points of infections with exponentially-distributed infectious periods (*Pois*(*β*) with rate *β* referring to the infection rate within the infectious period); (2) *mortality modeling*, e.g., by assuming a negative binomial distribution (*NB*(*μ, σ*)) or a Poisson distribution (*Pois*(*γ*)) with rate *γ* parameterized on mortality rate *γ*_*D*_ and population *N*). Further, basic reproduction number *R*_0_ may be estimated by *Nεβ/*(*β* + *γ*), the infections will be under control if *R*_0_ is less than a given threshold (e.g., 1).

The above hierarchical statistical model in Fig. 9(a) captures a general framework of COVID-19 statistical modeling. It can be customized to estimate and forecast COVID-19 case numbers in terms of specific hypotheses, settings, and conditions. For example, Fig. 9(b) shows the graphic model for the hierarchical model proposed in [88] to estimate death number *D* from its inferred variable 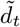 and inferred from auxiliary variable *ψ*. The inferred death number 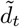 is sampled from the basic reproduction number *R*_0_ with a normally-distributed prior *Normal*(2.4, |*κ*|) parameterized by its variance variable *κ* and the probability of infected death *π* determined by two Gamma priors. In addition, 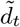 is also influenced by the number of new infections *c* with two latent variables, the distribution rate of the exponential distribution *τ* and the daily serial interval *g*, and a variable *α* as a parameter of the reproduction rate. Fig. 9(b) also shows the prior distributions of the auxiliary variables, for example, assuming the variable describing the time from infection to death *τ* following an exponential prior *Exponential*(0.03).

The hierarchical statistical model in Fig. 9(b) to estimate the death number can be described by the following equations.

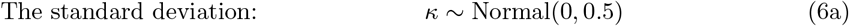

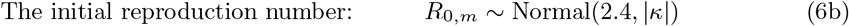

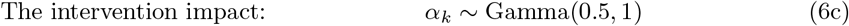

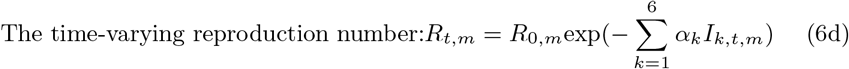

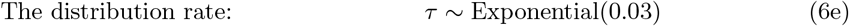

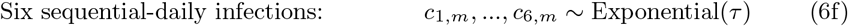

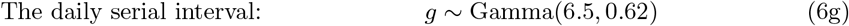

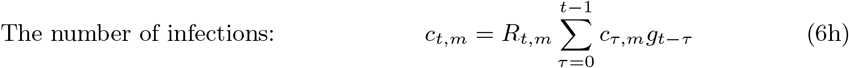

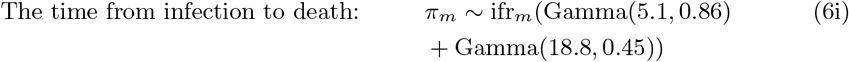

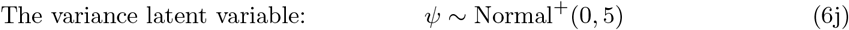

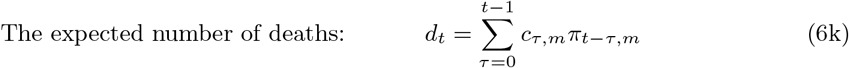

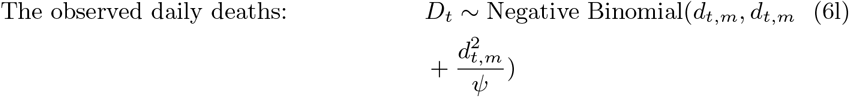

##### Other statistical models

Another major set of COVID-19 statistical modeling incorporates statistical hypotheses and settings into epidemic models such as compartmental models to approximate some state distributions or estimate some parameters. A typical application reformulates SIR-based models as a system of stochastic differential equations, e.g., by assuming the Gamma-distributed probability density of exposed *E* and infected *I* states in Section 7.1. In addition, modeling the influence of mitigation strategies on the COVID-19 case numbers (including asymptomatic infections) is also a typical statistical modeling problem [158] (this model also expands the general framework in Fig. 9(a)).

#### 5.2.2 COVID-19 statistical analysis

The COVID-19 pandemic has proven to be highly complex and uncertain. Statistical or probabilistic modeling naturally captures the uncertainties around epidemics better than other models. Statistical models have been widely applied to COVID-19 modeling. Typical tasks include (1) simulating and validating the state distributions and transitions of COVID-19 infected individuals over time; (2) modeling latent and random factors affiliated with the COVID-19 epidemic processes, movements and interactions; (3) forecasting short-to-long-term transmission dynamics; (4) evaluating the effect of NPIs; and (5) estimating the impact of COVID-19 such as on socioeconomic aspects. Below, we summarize the relevant applications of descriptive analytics, Bayesian statistical modeling, and stochastic compartmental modeling of the COVID-19 epidemic statistics, epidemic processes, and the influence of external factors such as NPIs on the epidemic. Table 2 further summarizes such applications of COVID-19 statistical modeling.

**Table 2.**
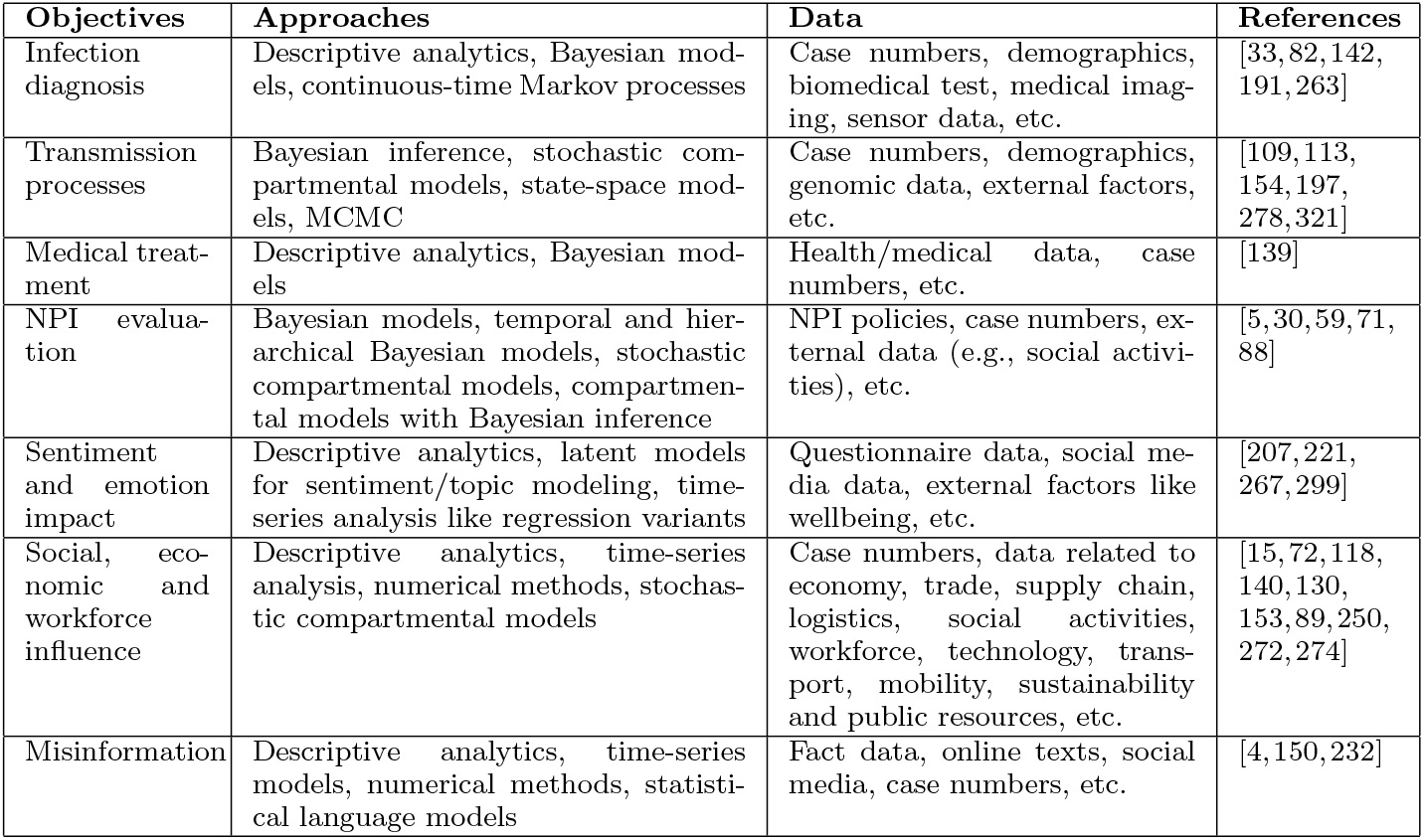
Summary and Examples of COVID-19 Mathematical and Statistical Learning.

| Objectives | Approaches | Data | References |
| --- | --- | --- | --- |
| Infection diagnosis | Descriptive analytics, Bayesian models, continuous-time Markov processes | Case numbers, demographics, biomedical test, medical imaging, sensor data, etc. | [33, 82, 142, 191, 263] |
| Transmission processes | Bayesian inference, stochastic compartmental models, state-space models, MCMC | Case numbers, demographics, genomic data, external factors, etc. | [109, 113, 154, 197, 278, 321] |
| Medical treatment | Descriptive analytics, Bayesian models | Health/medical data, case numbers, etc. | [139] |
| NPI evaluation | Bayesian models, temporal and hierarchical Bayesian models, stochastic compartmental models, compartmental models with Bayesian inference | NPI policies, case numbers, external data (e.g., social activities), etc. | [5, 30, 59, 71, 88] |
| Sentiment and emotion impact | Descriptive analytics, latent models for sentiment/topic modeling, time-series analysis like regression variants | Questionnaire data, social media data, external factors like wellbeing, etc. | [207, 221, 267, 299] |
| Social, economic and workforce influence | Descriptive analytics, time-series analysis, numerical methods, stochastic compartmental models | Case numbers, data related to economy, trade, supply chain, logistics, social activities, workforce, technology, transport, mobility, sustainability and public resources, etc. | [15, 72, 118, 140, 130, 153, 89, 250, 272, 274] |
| Misinformation | Descriptive analytics, time-series models, numerical methods, statistical language models | Fact data, online texts, social media, case numbers, etc. | [4, 150, 232] |

##### COVID-19 descriptive analytics

Descriptive analytics serve as the entrance to COVID-19 statistical analysis and are typically seen in non-modeling-focused references and communities. Typically, simple statistics such as the mean, deviation, trend and change of COVID-19 case numbers are calculated and compared. For example, the statistics of asymptomatic infectives are reported in [142]. In [22], change point analysis detects a change of the exponential rise of infected cases and the Pearson’s correlation between the change and lockdown implemented across risky zones. In addition, case statistics can be calculated in terms of specific scenarios, e.g., a population’s mobility [118] or workplace [15].

##### Bayesian statistical modeling of COVID-19 epidemic processes

Bayesian models have been favored by epidemiologists and health researchers for their capability of estimating uncertainty and involving factor dependence and prior knowledge [32], for example, modeling conventional epidemic contagion [10, 202]. In Bayesian analysis, MCMC has been particularly focused and widely applied in stochastic epidemic modeling [107]. MCMC is capable of simulating and estimating health and epidemic processes with a strong stochastic Markovian assumption, approximate model fitting, and parameter inference to complex settings. In COVID-19, Bayesian analysis and MCMC have been applied to model its stochastic epidemic processes, specific factors that may influence the COVID-19 epidemic process, causality, partially observed data (e.g., under-reported infections or deaths), and other uncertainties. For example, Niehus et al. [191] applied a Bayesian statistical model to estimate the relative capacity of detecting imported cases of COVID-19 by assuming the observed case count to follow a Poisson distribution and the expected case count to be linearly proportional to daily air travel volume. To capture the complex relations in the COVID-19 pandemic, the causal relationships in the coronavirus transmission process are modeled by hierarchical Bayesian distributions [191, 88]. In [82], a partially observable pure birth process following the continuous-time Markov population process assumes a binomial distribution of partial observations of infected cases and estimates the future actual values of infections and the unreported percentage of infections in the population.

##### Bayesian statistical modeling of external factors on influencing the COVID-19 epidemic

Another important application is to model the influence of external factors on the COVID-19 epidemic dynamics. For example, Flaxman et al. [88] inferred the impact of NPIs including case isolation, educational institution closure, banning mass gatherings, public events and social distancing (including local and national lockdowns) in 11 European countries. They estimated the course of COVID-19 by back-calculating the infections from observed deaths and fitting a semi-mechanistic Bayesian hierarchical model with an infection-to-onset distribution and an onset-to-death distribution. In addition, with case numbers, especially deaths, their model also jointly estimates the effect sizes of interventions.

##### Stochastic compartmental modeling of the COVID-19 epidemic

Stochastic compartmental models integrate statistical modeling with compartmental models (see Section 7.1). They can simulate the stochastic hypotheses of specific aspects (e.g., probability of a state-based population or of a state transition) of the COVID-19 epidemiological process and the stochastic influence of external interventions on the COVID-19 epidemic process. Such probabilistic compartmental models integrate the transmission mechanisms of epidemics with the characteristics of observed case data [321, 71, 197, 109]. For example, in [278], a COVID-19 transmission tree is sampled from the genomic data with MCMC-based Bayesian inference under an epidemiological model. The parameters of the offspring distribution in this transmission tree are then inferred. The model thus infers the person-to-person transmission in an early outbreak. Based on the probabilistic compartmental modeling, Zhou et al. [321] developed a semiparametric Bayesian probabilistic extension of the classical SIR model, called BaySIR. It involves time-varying epidemiological parameters to infer the COVID-19 transmission dynamics by considering the undocumented and documented infections and estimates the disease transmission rate by a Gaussian process prior and the removal rate by a gamma prior. To estimate the all-cause mortality effect of the COVID-19 pandemic, Kontis et al. [139] applied an ensemble of 16 statistical models (autoregressive with holiday and seasonal terms) on the vital statistics data for a comparable quantification of the weekly mortality effects of the first wave of COVID-19 and an estimation of the expected deaths in the absence of the pandemic. Other similar stochastic SIR models can also be found in the COVID-19 literature. Examples are assuming a Poisson time-dependent process on infection and reproduction [113], a beta distribution of infected and removed cases [281], and a Poisson distribution of susceptible, exposed, documented infected and undocumented infected populations in a city [154].

##### Statistical influence modeling of COVID-19 interventions and policies

Apart from modeling the COVID-19 transmission dynamics or forecasting case counts, Bayesian statistical models are also applied in other areas, e.g., to estimate the state transition distributions by applying certain assumptions such as of the susceptible-to-infected (i.e., the infection rate) or infected-to-death (mortality rate) transition. For example, Cheng et al. [59] used a Bayesian dynamic item-response theory model to produce a statistically valid index for tracking the government response to COVID-19 policies. Dehning et al. [71] combined the established SIR model with Bayesian parameter inference using MCMC sampling to analyze the time dependence of the effective growth rate of new infections and to reveal the effectiveness of interventions. With the inferred central epidemiological parameters, they sample from the parameter distribution to evolve the SIR model equations and thus forecast future disease development. In [281], a basic SIR model is incorporated with different types of time-varying quarantine strategies, such as government-imposed mass isolation policies and micro-inspection measures at the community level, to establish a method of calibrating the cases of under-reported infections.

###### Discussion

Statistical modeling and a Bayesian statistical framework offer various advantages in modeling COVID-19 uncertainties [158]. They can elicit informative priors for hidden parameters that are difficult to estimate due to the lack of data reflecting the clinical characteristics of COVID-19, offer coherent uncertainty quantification of parameter estimates, and capture nonlinear and non-monotonic relationships without the need for specific parametric assumptions [321]. Compared with compartmental models, statistical models usually converge at different confidence levels for different assumptions (i.e., possible mitigation strategies), providing better interpretability and flexibility for characterizing the COVID-19 characteristics and complexities discussed in Section 3. However, the related work on deep and comprehensive COVID-19 statistical modeling is still limited in terms of addressing COVID-19-specific characteristics and complexities. Open issues and opportunities include modeling asymptomatic effect, the couplings between mitigation measures and case numbers, and the time-evolving and nonstationary case movement of COVID-19 clusters.

## 6 COVID-19 Data-driven Learning

Here, we review the related work on data-driven discovery in COVID-19. Data-driven COVID-19 modeling covers classic (shallow) and deep machine learning methods and various AI and data science techniques on COVID-19 problems and data. They discover interesting knowledge and insights by characterizing, representing, analyzing, classifying and predicting COVID-19 problems.

### 6.1 Shallow and Deep Learning

Both classic (shallow) machine learning models and deep neural networks have been intensively applied in modeling COVID-19, as shown by the commonly used keywords in the literature [44] (also see Figs. 6, 7 and 8). Below, we summarize the relevant machine learning models customized for COVID-19 modeling.

#### Shallow learning models

Classic (shallow) machine learning methods have been intensively applied in COVID-19 classification, prediction and simulation, as shown in [44]. Typical shallow learning methods include artificial neural networks (ANN), SVM, decision trees, Markov chain models, random forest, reinforcement learning, and transfer learning. These tools are easy to understand and implement and are more applicable than other sophisticated methods (e.g., deep models and complex compartmental models) for the often-small COVID-19 data. They are well explained in the relevant literature (e.g., [56]). Interested readers can refer to them and other textbooks for technical details [122, 25, 42].

Though different machine learning methods may be built on their respective learning paradigms [42], and therefore differ from each other, their main learning tasks and processes for modeling COVID-19 are actually similar. They (1) select discriminative features **x**, (2) design a model *f* (e.g., a random forest classifier) to predict the target 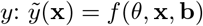 with parameters *θ* and bias term **b**, and then (3) optimize the model to fit the COVID-19 data by defining and optimizing an objective function 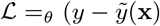 for the goodness of fit between expected 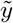 and actual *y* target (e.g., infective or diseased case numbers).

#### COVID-19 deep sequential interaction modelling

As discussed in Section 3.3, COVID-19 involves various sources of data. COVID-19 case numbers including infected and death cases are temporal. External factors such as NPIs, vaccination, mobility and environmental factors may be temporal and sequential as well. The case series and external factors interact with each other over time. A critical challenge in modeling COVID-19 (see Section 3.4) is to model the temporal and sequential interactions between multiple series over time. Few deep models serve this purpose, thus we highlight this important matter here.

Typical approaches for COVID-19 deep sequential interaction modeling can be characterized by a general deep interaction and prediction framework as follows. They model (1) temporal dependencies over sequential *case* (**x**, which may consist of categories of case numbers *s, i* and *r* or their rates) evolution, (2) the interactions and influence between external containment *actions* (**a**, which may consist of various control measures such as masking and social distancing) and case developments, and (3) the influence of personal *context* (**c**, which may consist of demographic, health circumstances and symptomatic features on COVID-19 infections) over time *t*. As COVID-19 case developments are sequential, stochastic and influenced by many external factors, such a framework can combine (1) autoencoders for representing the influence of unknown and stochastic asymptomatic and unreported case dynamics on reported numbers **x**; (2) RNNs for capturing the sequential evolution of case numbers, control measures and personal context; and (3) the contextual attention network for capturing the exterior containment strategies applied on case control. These DNN networks jointly model complex interactions between various sources of underlying and control factors in COVID-19 sequential developments.

Fig. 10 illustrates a deep sequential case-action-context interaction network for implementing the above framework. The formulation of the key variables and their interactions are shown below. In practice, networks **h** can be based on an LSTM, RNN, Transformer or other deep networks. The gating function unit *g* can be implemented by a gated recurrent unit (GRU) or other gating functions to determine the influence of control actions/context on case number movement. The transformation from the input case vector **x** to the action **a** and context **c** adjusted case representation **e** through network *q*_*θ*_ can be treated as an encoder. The estimation (reconstruction) of **x**_*t*_ from **e** by network *p*_*φ*_ is a decoding or prediction process.

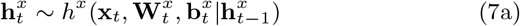

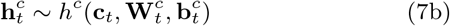

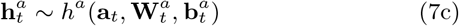

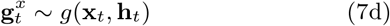

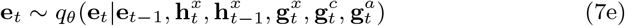

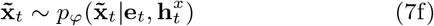

**Fig. 10.**
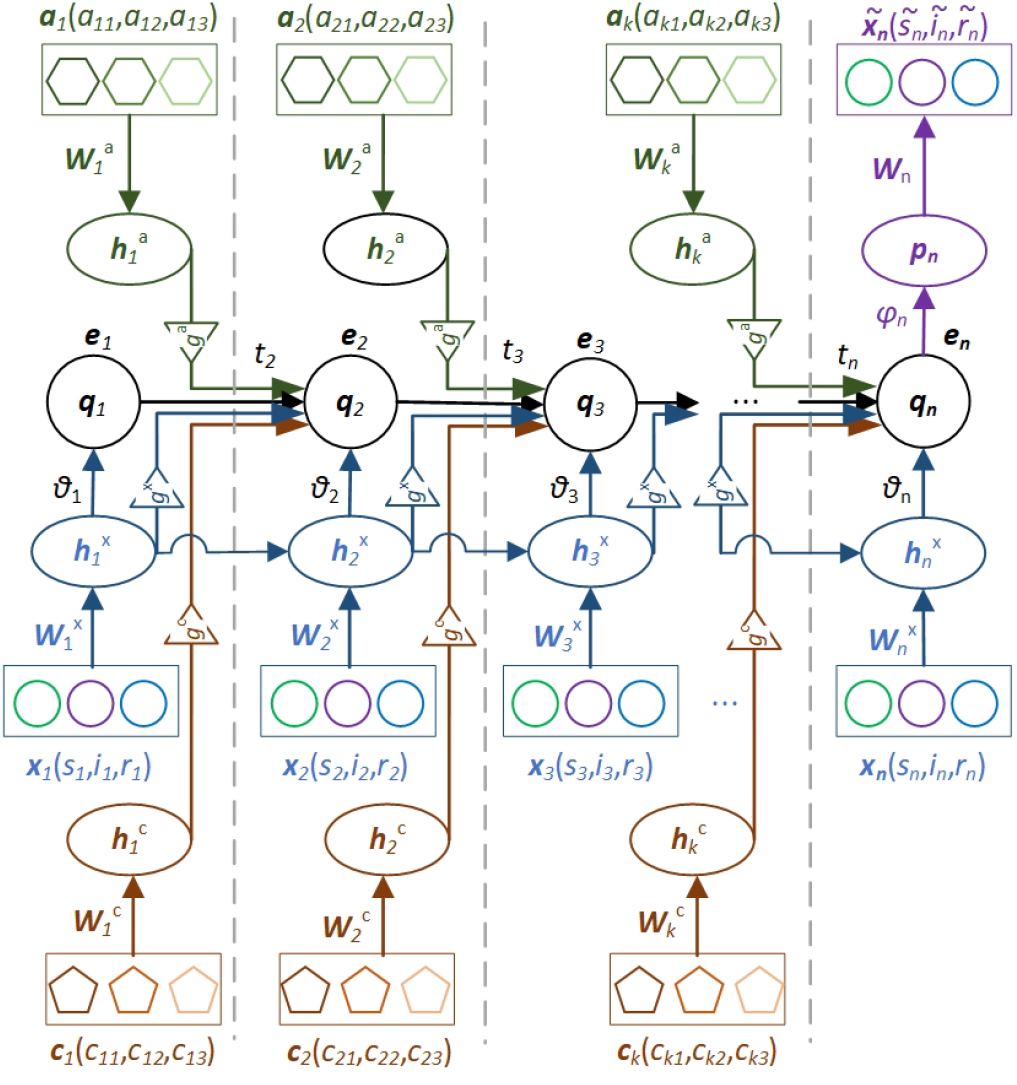
A time-varying case-action-context neural interaction network for COVID-19 deep sequential modeling.

The interaction and prediction network for COVID-19 in Fig. 10 can be implemented in terms of an autoencoder (where *q* and *p* refer to encoding and decoding networks, e.g., [119]) or LSTM/RNN-based prediction (with *q* for representation and *p* for estimating the next input, e.g., [240]) framework. Accordingly, the objective function can be defined in terms of the discrepancy 𝒥 between **x**_*t*_ and 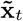 (i.e., 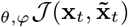) or the KL-divergence (𝒟) with loss ℒ (where **h**_*t*_ and **h**_*t−* 1_ refer to the representations of input **x** interacting with actions **a** under the context **c** through gating *g* integration).

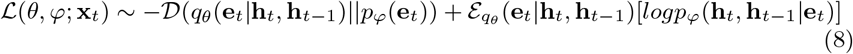

#### Other deep learning models

Deep learning [96] as represented by DNNs serves as a more advanced approach to modeling COVID-19, typically favored by computing researchers. Pretrained DNNs are widely applied in modeling COVID-19 data, in particular, X-ray medical images, texts, news, and social media. Numerous references can be found in the literature [44] which apply DNNs in addressing various COVID-19 problems, in particular, medical imaging and textual data. Typical DNNs applied in COVID-19 modeling include (1) convolutional neural networks (CNNs) and their extensions such as ImageNet and ResNet in particular for COVID-19 images; (2) sequential networks such as LSTM, recurrent neural networks (RNNs), memory networks, Transformer and their variants for COVID-19 case series; (3) textual neural networks such as BIRT, Transformer and their variants for COVID-19 texts and news; (4) unsupervised neural networks such as autoencoders and generative adversarial networks (GANs) for COVID-19 case estimation; and (5) other neural learning mechanisms such as attention networks for incorporating contextual factors.

### 6.2 COVID-19 Shallow Learning

Here, *COVID-19 shallow learning* refers to applying general analytics methods and classic machine learning models in the analytics and modeling of COVID-19 problems and data. In the literature, about 8.5k of 44k modeling references are on COVID-19 shallow learning, which forms the second most commonly used set of modeling methods. COVID-19 shallow learning has been widely applied in studying broad-reaching COVID-19 problems by medical, biomedical, computing and social scientists [241, 182, 228, 242, 145, 304]. The analytics and learning methods most applied include ANN, tree models such as decision trees and random forest, clustering methods, classification methods, kernel methods like SVM, transfer learning, federated learning, NLP and text mining methods. In addition, evolutionary computing like genetic algorithms and fuzzy set, and reinforcement learning have also been involved in COVID-19.

Shallow learning methods have been applied to address various COVID-19 problems and tasks. Examples are COVID-19 outbreak, case forecasting, medical diagnostics, contact tracing, risk, transmission, uncertainty, anomalies, complexities, classification, variation, prediction, and drug development [44]. Below, we briefly illustrate their applications in several common COVID-19 problems and tasks: COVID-19 outbreak prediction and risk assessment, COVID-19 diagnosis on clinical attributes, COVID-19 diagnosis on respiratory data, COVID-19 diagnosis on medical imaging, COVID-19 diagnosis on latent features, modeling the influence of external factors on COVID-19, and COVID-19 drug and vaccine development. Table 3 further summarizes and illustrates various applications of COVID-19 shallow learning.

**Table 3.** Summary and Examples of COVID-19 Shallow and Deep Learning^*a*^.

| Objectives | Approaches | Data | References |
| --- | --- | --- | --- |
| COVID-19 transmission & external factor impact on transmission | Shallow learners like SVM, ANN, decision tree, random forest and ensemble methods; evolutionary computing methods such as particle swarm optimization; DNN variants such as LSTM, GRU, VAE, GAN and BiLSTM, etc. | Epidemic case numbers, external data such as meteorological data, environmental data (e.g., humidity), social activity, and mobility data, etc. | [128,190, 234,261, 74,240,138, 210,312] |
| COVID-19 infection diagnosis | Shallow learners; CNN and RNN variants like LSTM and GRU; pretrained CNN-based image nets like ResNet, MobileNetV2, and SqueezeNet; and text analysis models | Pathological and clinical records, respiratory signals (e.g. coughing and breathing signals and patterns in ultrasound or thermal video), computed tomography (CT), and CXR images, etc. | [31,31,33, 51,85,259, 262,123, 135,171, 301] |
| COVID-19 mortality and survival analysis | Shallow learners like SVM, ANN, decision tree, regression tree, and random forest; ensemble methods like XGBoost; CNNs, pretrained CNN-based image nets, RNN variants like LSTM and GRU | Medical imaging including CT and CXR images, clinical records, patient demographics, case numbers, external data, etc. | [237,49, 237,74,240, 313] |
| COVID-19 medical treatment | Shallow machine learning methods, and DNNs, etc. | Health and medical records, pharmaceutical treatments, ICU data, etc. | [320,293, 20] |
| COVID-19 genomic and protein analysis, drug and vaccine development | Shallow classifiers like SVM and ensembles, frequent pattern and sequence analysis methods, CNN variants, RNN variants, attention networks, GAN, autoencoders, reinforcement learning, NLP models like Transformer, etc. | Genomic data, proteomic data, drug-target interactions, molecular reactions, etc. | [132,177, 20,317,176, 187,8] |
| COVID-19 resurgence and mutation | Shallow learners like linear discriminant, SVM, KNN and subspace discriminant, classifiers combined with compartmental models, DNN variants, sequence analysis<sup>b</sup>, etc. | Resurgence case numbers, virus strain genome and protein sequences, NPI data, external data, etc. | [227,12] |
| COVID-19 NPI evaluation | Various Bayesian models<sup>c</sup>, compartmental models combined with classifiers or estimators, DNNs<sup>b</sup>, etc. | Case numbers, NPI policies, external data, etc. | [84,92] |
| COVID-19 sentimental and emotional impact | NLP models like LDA and topic models, DNN variants like BERT and Transformer variants, etc. | Social media data, news feeds, Q/A data, external factors, etc. | [78,286, 188,181] |
| COVID-19 socioeconomic influence | Relation (e.g., correlation and causality) analysis<sup>b</sup>, topic modeling by NLP models | Social, economic and workforce activities, case numbers, etc. | [316,26] |
| COVID-19 misinformation analysis | Classic NLP models, correlation analysis, shallow learners, outlier detectors, DNN variants like BERT and Transformer mutations | Social media, online texts, Q/A data, news feeds, etc. | [178,150] |

<sup>a</sup> See Table 6 for deep COVID-19 medical imaging analysis; <sup>b</sup> such methods are applicable, however, not much work has been reported in the literature; <sup>c</sup> See Table 2 for the NPI effect modeling.

#### Machine learning for COVID-19 outbreak prediction and risk assessment

Typical classifiers like ANN, SVM, decision trees, random forest, regression trees, least absolute shrinkage and selection operator (LASSO), and self-organizing maps have been applied to forecast COVID-19 spread and outbreak and their coverage, patterns, growth, and trends; estimate and forecast the confirmed, recovered and death case numbers, and the transmission and mortality rates; and cluster infected cases and groups, etc. For example, in [128], logistic regression, decision trees, random forest and SVM are applied to estimate the growth trend and containment sign on the data consisting of factors about health infrastructure, environment, intervention policies and infection cases with accuracy between 76.2% and 92.9%. Evolutionary computing such as genetic algorithms, particle swarm optimization, and gray wolf optimizer forecast COVID-19 infections [190, 234, 261].

#### Machine learning for COVID-19 diagnosis on clinical attributes

The machine learning of COVID-19 clinical reports such as blood test results is applied to assist in COVID-19 diagnosis. For example, in [135], clinical attributes and patient demographic data were extracted by term frequency/inverse document frequency (TF/IDF), bag of words (BOW) and report length from textual clinical reports. The extracted features were then classified in terms of COVID, acute respiratory distress syndrome (ARDS), SARS, and both COVID and ARDS by SVM, multinomial naive Bayes, logistic regression, decision trees, random forest, bagging, Adaboost, and stochastic gradient boosting. It reports an accuracy of 96.2% using multinomial naive Bayes and logistic regression. In [31], hematochemical values were extracted from routine blood exam-based clinical attributes, which were then classified into positive or negative COVID-19 infections by decision trees, extremely randomized trees, KNN, logistic regression, naive Bayes, random forest, and SVM. It reports an accuracy of 82% to 86%. The work in [31] applied decision tree to identify and explain suspicious COVID-19 cases.

#### Machine learning for COVID-19 diagnosis on respiratory data

Machine learning is applied to COVID-19 patient respiratory data, such as lung ultrasound waves and breathing and coughing signals, to extract respiratory behavioral patterns and anomalies. For example, logistic regression, gradient boosting trees and SVMs were used to distinguish COVID-19 infections from asthmatic or healthy people with 80% AUC on the Android app-based collection of coughs and breathing sounds and symptoms in [33].

#### Machine learning for COVID-19 diagnosis on medical imaging

A very intensive application of classic machine learning methods is to screen COVID-19 infections on CT, chest X-ray (CXR) or PET images. For example, in [51], the majority voting-based ensemble of SVM, decision trees, KNN, naive Bayes, and ANN was applied to classify normal, pneumonia and COVID-19-infected patients on CXR images. It reports an accuracy of 98% and AUC of 97.7%. In [85], the simple applications of SVM, naive Bayes, random forest, and JRip on CT images screen COVID-19 diseases. It reports 96.07% accuracy by naive Bayes combined with random forest and JRip, in comparison with 94.11% by CNN.

#### Machine learning for COVID-19 diagnosis on latent features

Shallow learners have also been applied to detect and diagnose COVID-19 infections on latent features learned by shallow to deep representation models on COVID-19 medical images. For example, in [127], ANN-based latent representation learning captures latent features from gray, texture, histogram, number, intensity, surface and volume features in CT images. Classifiers including SVM, logistic regression, Gaussian naive Bayes, KNN, and ANN then differentiate COVID-19 infections from community-acquired pneumonia with 95.5% accuracy reported. In [199], latent features were extracted from CXR and CT images to form a gray level co-occurrence matrix (GLCM), local binary gray level co-occurrence matrix (LBGLCM), gray level-run length matrix (GLRLM), and segmentation-based fractal texture analysis (SFTA)-based features. These features were then oversampled by the synthetic minority over-sampling technique (SMOTE) and further selected by a stacked autoencoder (sAE) and principal component analysis (PCA), before SVM was applied to achieve 94.23% accuracy.

#### Modeling the influence of external factors on COVID-19

Various machine learning tasks have been undertaken to analyze the relation and influence of external and contextual factors on COVID-19 epidemic attributes. For example, ensemble methods including random forest, extra trees regressor, AdaBoost, gradient boosting regressor, extreme gradient boosting (XGBoost), light gradient boosting machine (LightGBM), CatBoost regressor, kernel ridge, SVM, KNN, MLP, and decision trees were used to estimate the potential association between COVID-19 mortality and weather data in [173].

#### Machine learning-driven COVID-19 drug and vaccine development

Machine learning methods have also been applied to analyze the drug-target interactions, drug selection, and the effectiveness of drugs and vaccines on containing COVID-19. For example, machine learning methods including XGBoost, random forest, MLP, SVM and logistic regression were used to screen thousands of hypothetical antibody sequences and select nine stable antibodies that potentially inhibit SARS-CoV-2 in [169, 132].

### 6.3 COVID-19 Deep Learning

Deep learning has been intensively applied to modeling COVID-19 as discussed in Section 6.1. As reported in [44], about 4.6k of 4k modeling publications are associated with deep learning of COVID-19 [110, 121]. Typical applications involve COVID-19 data on daily infection case numbers, health and clinic records, hospital transactions, medical imaging, respiratory signals, genomic and protein sequences, and exterior data such as infective demographics, social media communications, news and textual information, etc. Below, we first discuss the broad deep COVID-19 learning, and then illustrate two of the most commonly used applications of deep learning for COVID-19 epidemic forecasting and image analysis.

#### Broad deep COVID-19 learning

By reviewing the related literature, we summarize and highlight the following typical application areas of COVID-19 deep learning.

- Characterizing the symptoms of coronavirus infections and COVID-19, e.g., by pretrained neural networks, e.g., [236]; with more discussion in Section 7.2.1;
- Analyzing health and medical records, blood sample-based test reports, and respiratory sounds and signals for COVID-19 diagnosis and treatment, e.g., by CNN, LSTM, and GRU [228]; with more discussion in Section 7.2.2;
- Analyzing medical imaging for COVID-19 diagnosis, quarantine and treatment, e.g., by CNNs such as ImageNet and ResNet, GANs, and their mutations [121, 228]; with more discussion in Section 7.2.3;
- Analyzing COVID-19 genomic and protein sequences and interactions, e.g., by RNNs, CNNs and their variants, for drug and vaccine development, infection source tracing, and virus structure and evolution analysis; with more details in Section 7.2.4;
- Repurposing and developing COVID-19 drugs and vaccines, e.g., by generative autoencoders, generative tensorial reinforcement learning, and GANs [317] for generative chemistry discovery;
- Analyzing the COVID-19 impact on sentiment and emotion, e.g., by RNN, Transformer-based NLP neural models, and their derivatives [78];
- Characterizing the COVID-19 infodemic, e.g., by NLP and text mining including misinformation identification [93, 243, 178, 19], enhancing epidemic modeling using social media data [137], and analyzing COVID-19 research progress and topic evolution [316];
- Other tasks such as analyzing the influence and effect of countermeasures, e.g., the effect of quarantine policies on outbreak using DNNs; with more discussion in Section 8.

Further, Table 3 illustrates some typical applications of deep learning methods for modeling COVID-19.

#### Deep learning of the COVID-19 epidemic

Deep neural networks have been intensively applied to characterize and forecast COVID-19 epidemic outbreak, dynamics and transmission. Examples are predicting the peak confirmed numbers and peak occurrence dates, forecasting daily confirmed, diseased and recovered case numbers, and forecasting the next *N* -day (e.g., *N* = {7, 14, 10, 30, 60 *days*}) infected, confirmed, recovered and death case numbers or their transmission/mortality rates. In such problems, short-range temporal dependencies in case numbers can be modeled by LSTM, stacked LSTM, bi-LSTM, convolutional LSTM-like RNNs, and GRU [74, 240]. Other work models the transmission dynamics and predicts the daily COVID-19 infections using a variational autoencoder (VAE), encoder-decoder LSTM, LSTM with encoder and Transformer [138], modified autoencoder [210], GAN and their variants. In [312], a comparative analysis shows that VAE outperforms simple RNN, LSTM, BiLSTM and GRU in forecasting COVID-19 new and recovered cases. In addition, DNNs are also used to track the COVID-19 outbreak, predict the outbreak size by encoding quarantine policies as the strength function in a deep neural network, estimate global transmission dynamics using a modified autoencoder [117], predict epidemic size and lasting time, and combine medical information with local weather data to predict the risk level of a country using a shallow LSTM model [203].

#### Deep learning of COVID-19 medical imaging

COVID-19 medical imaging analysis is an area mostly explored by deep learning methods. Typical COVID-19 data includes CXR, CT, ultrasound, and multimodal data. Problems related to COVID-19 medical imaging analysis include diagnosing and detecting COVID-19 and differentiating COVID-19 infections from viral and bacterial pneumonia in COVID-19 images, such as lung ultrasound images, CT and CXR images; localizing and segmenting COVID infections; representing what the coronavirus infection on the lungs looks like; and visualizing COVID infections. DNNs applied on COVID-19 ultrasound imaging include LSTM, VGG variants, LSTM and CNN combinations, and CNNs. Deep models applied on CXR images include various CNNs, such as AlexNet, CheXNet, COVIDX-Net, DenseNet201, DenseNet121, GoogleNet, InceptionV3, InceptionResNetV2 (Inception-ResNetV2), MobileNetV2, NASNet, ResNetV2, ResNet18, ResNet50, ResNet101, ShuffleNet, SqueezeNet, VGG16, VGG19, Xception, and XceptionNet. For example, the COVID-Net [279] pretrained on ImageNet detects COVID-19 on CXR images and achieves 93.3% test accuracy. In [259], MobileNetV2 and SqueezeNet extract features from CXR images, which are then processed by social mimic optimization to classify coronavirus, pneumonia, and normal images with 99.27% accuracy by SVM. More discussion on COVID-19 deep learning can be found in Section 7.2 for COVID-19 medical and biomedical analysis.

##### Discussion

Most existing studies on shallow and deep COVID-19 modeling directly apply existing shallow machine learning methods and pretrained deep neural networks on COVID-19 data, as shown in reviews like [156, 239, 279]. Our literature review also shows that DNNs have been widely applied to COVID-19 modeling. However, they are unnecessarily applicable to any possibilities and do not always significantly outperform shallow models such as time-series forecasters and shallow machine learners. In fact, sometimes, deep models may even lose their advantage over traditional modelers such as ensembles, as shown in Table 3, and Table 6. This may be caused by the often small but complex COVID-19 data, as discussed in Section 3.3, making deep models incapable of sufficient fitting.

**Table 6.** Summary and Examples of Deep COVID-19 Medical Imaging Analysis.

| Method | Task | Data | Performance |
| --- | --- | --- | --- |
| Domain extension transfer learning with pretrained CNNs [18] | Diagnosis | Italian Society of Medical Radiology and Interventional (25 cases)<sup>a</sup>; Radiopaedia.org (20 cases)<sup>b</sup>; COVID-19 images (180 cases) [64]; A Spanish hospital (80 cases)<sup>c</sup> | Overall accuracy 90.13% ± 0.14 |
| Shallow CNN [183] | Diagnosis | COVID-19 images [64] (321 cases); Kaggle non-COVID-19 CXR images (5856 cases)<sup>d</sup> | Highest accuracy 99.69%, sensitivity 1.0, AUC 0.9995 |
| CNN-based truncated Inception-Net [69] | Diagnosis | COVID-19 images (162 cases) [64]; Kaggle CXR images (5863 cases); Tuberculosis CXR images<sup>e</sup> | Accuracy 99.96% (AUC of 1.0) in classifying COVID-19 cases combining pneumonia and healthy cases; accuracy 99.92% (AUC of 0.99) in classifying COVID-19 cases combining pneumonia, tuberculosis and healthy CXRs |
| DarkCovidNet [200] built on Darknet-19 | Diagnosis | COVID-19 images (127 cases) [64]; CXR images [287] | Accuracy 98.08% for binary classes and 87.02% for multi-class cases |
| CoroNet built on Xception pretrained on ImageNet [133] | Diagnosis | COVID-19 images [64]; Kaggle CXR images | Accuracy 89.6%, precision 93% and recall 98.2% for COVID vs bacterial pneumonia, viral pneumonia, and normal |
| COVID-CAPS based on Capsule net [2] | Diagnosis | COVID-19 images [64], Kaggle CXR images | Accuracy 95.7%, sensitivity 90%, specificity 95.8%, and AUC 0.97 |
| VGG16 and transfer learning [192] | Diagnosis | COVID-19 images [64], RSNA Pneumonia Detection Challenge data<sup>f</sup> | Accuracy 83.6% for COVID-19 pneumonia vs non-COVID-19 pneumonia and healthy; sensitivity 90% for COVID-19 pneumonia |
| COVID-Net [279] | Diagnosis | COVID-19 images [64], COVID-19 CXR Dataset Initiative<sup>g</sup>, ActualMed COVID-19 CXR Dataset Initiative<sup>h</sup>, RSNA data, COVID-19 radiography data<sup>i</sup> | Accuracy 93.3%, sensitivity 91.0%, positive predictive value 98.9% |
| Inf-Net with decoder and attention [64] | Segment | COVID-19 CT Segmentation<sup>j</sup>, COVID-19 CT Collection | Dice similarity coefficient 0.739, sensitivity 0.725, specificity 0.960 |
| DeepPneumonia built on ResNet-50 [249] | Diagnosis | Private data | Sensitivity 0.93, AUC 0.99; AUC 0.95 and sensitivity 0.96 for COVID-19 vs. bacteria pneumonia-infections |

<sup>a</sup>https://www.sirm.org/category/senza-categoria/covid-19/;
<sup>b</sup>[https://radiopaedia.org/search?utf8=%E2%9C%93&q=covid&scope=all&lang=us](https://radiopaedia.org/search?utf8=%E2%9C%93&q=covid&scope=all&lang=us;);
<sup>c</sup>https://twitter.com/ChestImaging/status/1243928581983670272;
<sup>d</sup>https://www.kaggle.com/paultimothymooney/chest-xray-pneumonia;
<sup>e</sup>https://ceb.nlm.nih.gov/tuberculosis-chest-X-ray-image-data-sets/;
<sup>f</sup>https://www.kaggle.com/c/rsna-pneumonia-detection-challenge;
<sup>g</sup>https://github.com/agchung/figure1-COVID-chest-xray-dataset;
<sup>h</sup>https://github.com/agchung/Actualmed-COVID-chest-xray-dataset;
<sup>i</sup>https://www.kaggle.com/tawsifurrahman/covid19-radiography-database;
<sup>j</sup>https://medicalsegmentation.com/covid19/.

## 7 COVID-19 Domain-driven Modeling

To address the physical-technical nature of COVID-19, here, we focus on the two most relevant physical-technical domains of COVID-19: epidemic modeling, and medical and biomedical analysis.

### 7.1 COVID-19 Epidemic Modeling

Epidemic modeling is a research area intensively studied for modeling the epidemic systems, infection, transmission and dynamics of infectious diseases, typically by combining epidemiology and mathematical modeling. Here, we introduce the relevant epidemiological compartment models and their applications in modeling COVID-19.

#### 7.1.1 Epidemiological compartmental models

Epidemiological modeling [10, 202, 245, 32, 107] portrays the state-space, interaction processes and dynamics of an epidemic in terms of its macroscopic population, states and behaviors. Compartmental models [58, 201, 158] have been widely used in characterizing COVID-19 epidemiology by incorporating epidemic knowledge and compartmental hypotheses into imitating the multi-state COVID-19 population transitions. An individual in the COVID-19 epidemic sits at one state (compartment) at a time-point and may transit this state to another at a state transmission rate. The individuals of the closed population are respectively labeled per their compartments and migrate across compartments during the COVID-19 epidemic process, which are modeled by (ordinary) differential equations.

By consolidating various COVID-19 epidemiological characteristics and hypotheses, Fig. 11 illustrates a general and typical COVID-19 state-space and evolution compartmental system with major (thick) and minor (thin) states and state transition paths. This compartmental modeling framework decomposes the COVID-19 epidemic process into four major sequential states: Susceptible (S), Exposed (E), Infective (I), and Removed (R).

**Fig. 11.**
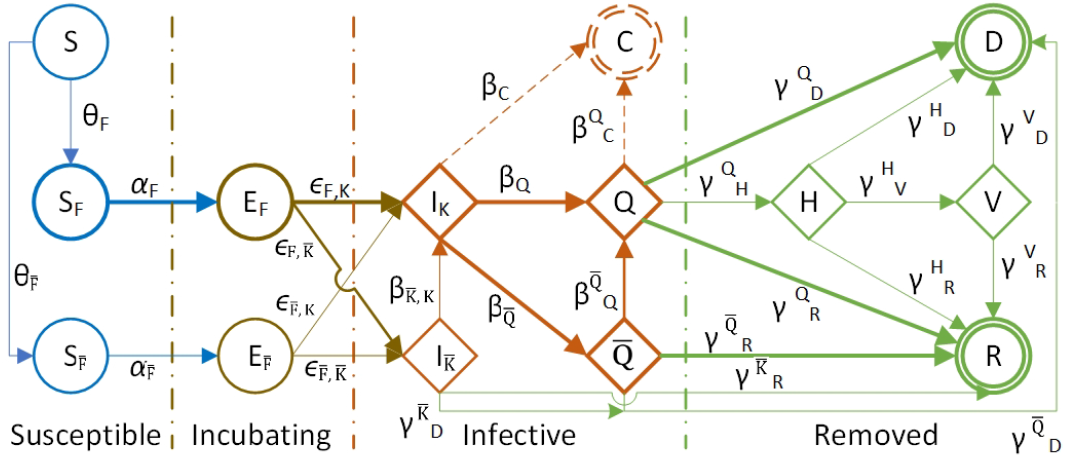
A general framework of COVID-19 epidemiological compartmental modeling. The COVID-19 epidemiological process can be categorized into four major states: susceptible (S), exposed/incubating (E), infective (I) and removed (R), and other optional states. A *circle* denotes the initial states, a *diamond* denotes the optional states, and a *double circle* denotes the final states. *Blue* indicates the noninfectious states, *red* for the infectious states, and *green* for the states with patients removed or to be removed from the infection. The *thick* lines denote the main states and state transition paths while the *thin* lines are minor ones which may be ignored in modeling.

- Susceptible (S): Individuals (*S*) are susceptible to infection under free (uncontained or unrestrained, *S*_*F*_ at uncontained rate *θ*_*F*_) or contained (restrained, 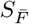 at containment rate 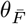) conditions at a respective transmission rate *α*_*F*_ or 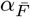;
- Exposed (E): Free or contained susceptibles are exposed to infection from those who are infected but in the incubation period (which could be as long as 14 days), and may be noninfectious and free (*E*_*F*_) or infectious and contained 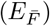 at a respective exposure rate *ε*_*F*_ or 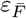;
- Infective (I): Those exposed become infectious and may be detected (registered/documented and known to medical authorities, *I*_*K*_) or undetected (unreported/undocumented and unknown to medical management, 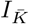); also, some initially undetected infectives may be further detected and converted to detected infectives at rate 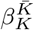; some documented infectives may be symptomatic and quarantined (*Q*) at quarantine rate *β*_*Q*_ while others may be asymptomatic and unquarantined 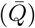 at unquarantined rate 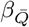; there may be some rare cases (at rate *β*_*C*_) who carry the virus and infection for a long time with or without symptoms, called lasting *carriers* (*C*); in addition, some initially asymptomatic cases may transfer to symptomatic and quarantined at rate 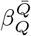;
- Removed (R): Unquarantined infectives may recover (R) at recovery rate 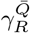 or die (D) at mortality rate 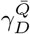 respectively; at rate 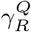 or 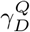 for the quarantined infectives; and at rate 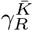 or 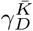 for unknown/undetected infectives; some quarantined infectives may present acute symptoms even with life threats, who are then hospitalized (H) at rate 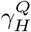 or even further ventilated (V) at rate 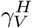; hospitalized infectives may recover or die at rate 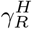 or 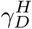, and at rate 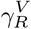 or 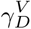 for ventilated.

As noted in Fig. 11, this general epidemic modeling framework further decomposes the SEIR states into specific conditional states: noninfectious and free exposed state (*E*_*F*_) vs infectious and contained state 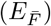, detected infectious state (*I*_*K*_) vs undetected infectious state 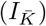 quarantined infective state (*Q*) vs unquarantined infective state 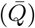. The framework also incorporates a few optional states: *carriers* (*C*), recovery (*R*), deceased (*D*), hospitalized (*H*), or ventilated (*V*).

In practice, the above comprehensive COVID-19 state-space system is often instantiated into specific SEIR models in the literature, where not all states are characterizable by their available data. Accordingly, the focus in COVID-19 epidemic modeling is on those main epidemiological compartments and their transitions when the corresponding data is available. Examples are the susceptible, exposed, infectious (which consists of both detected and undetected), recovered, and diseased states [58].

Below, we illustrate the general COVID-19 epidemic modeling framework in Fig. 11 in terms of the differential equations with states 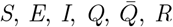 and *D*, which are the main states of a closed COVID-19 population. Here, (1) 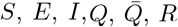 and *D* represent the fraction of the population at each state; (2) *S* and *E* are impacted by containment measures at the containment rate *θ* and *I* at *θ*_*I*_ who are contained and recovered; and (3) the state transitions take place at their respective rates.

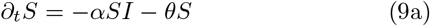

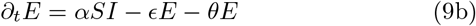

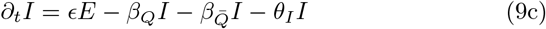

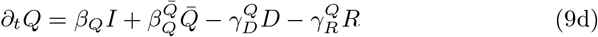

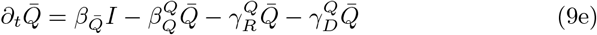

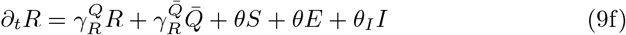

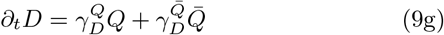

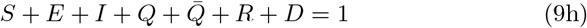

Both the COVID-19 epidemic framework in Fig. 11 and the above differential equations can be further customized and connected to many compartmental models in the literature [44, 305, 58].

#### 7.1.2 COVID-19 epidemiological modeling

Here, building on the above discussion on epidemic modeling in Section 7.1.1, we summarize and discuss the related work on the major tasks of COVID-19 epidemiological modeling, and highlight the related work on modeling COVID-19 epidemic transmission processes, dynamics, external factor influence, and resurgence and mutation.

##### COVID-19 epidemiological modeling tasks

Epidemiological models dominate COVID-19 modeling (about 5.5k publications of the 4k reported in the literature analysis [44]). Epidemic researchers and computing scientists expand or hybridize epidemic models with other modeling methods such as statistical modeling and machine learning for a more powerful characterization of COVID-19 complexities. In general, COVID-19 compartmental modeling aims to address several epidemiological problems: (1) quantifying the growth (spread) of COVID-19 and its case number movements at different epidemiological states to forecast case numbers in the next days or periods; (2) quantifying the basic reproduction rate *R*_0_ that informs the contagion and transmission level and control strategies for COVID-19; quantifying the sensitivity and effect of control measures on COVID-19 infection containment and case movements; and (4) quantifying the sensitivity and effect of strategies for COVID-19 herd immunity and mass vaccination.

Accordingly, various compartmental models have been customized to cater for specific assumptions, settings and conditions in modeling COVID-19, as discussed in Section 7.1.2. To address the above objective (1), with historical case numbers of a country or region and the initial settings of hyperparameters, epidemic modeling can estimate the parameters and further predict numbers over time, e.g., the number of infections and deaths in a country or city. Regarding (2), with the state-space shown in Fig. 11, by resolving their differential equations, we can first obtain the population projection matrix *A* corresponding to all states and their transition probabilities. The projection matrix *A* can be converted to a state transition matrix *T*, where each element *T*_*ij*_ is the probability of an individual transfer from state *i* at time *t* to state *j* at time *j* + 1, and a fertility (reproductive) matrix *F*, where an element *F*_*ij*_ refers to the reproduced number of *i*-state offsprings of an individual at state *j*, i.e., *A* = *T* + *F*. Further, we can calculate the fundamental matrix *N* : *N* = (*I* − *T*)^*−* 1^ with the identity matrix *I* to represent the expected time spent in each state and the time to death. Then, we can obtain another matrix *R*: *R* = *FN* with each entry referring to the expected lifetime production number of *i*-state offspring by an individual at stage *j* [47, 245]. Its dominant eigenvalue is the net reproduction rate *R*_0_. With regard to (3), since control measures such as social distancing and lockdown may influence the growth of case numbers and reproduction and transmission rates, we can analyze the sensitivity of adjusting related parameters on the case numbers and rates. To explore the opportunities for herd immunity and mass vaccination in (4), the herd immunity rate and vaccination rate are expected to be greater than 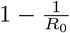 to eradicate the disease.

These major objectives may be further connected to other COVID-19 modeling tasks, applying epidemiological modeling to modeling COVID-19 transmission (which is also the most explored area) and resurgence and mutation (which is a recent challenge).

##### Modeling COVID-19 epidemic transmission processes

The studies on modeling COVID-19 epidemic transmission mainly focus on evaluating the epidemiological attributes (e.g., infection rate, recovery rate, mortality, reproduction number, etc.), predicting the infection and death counts, and revealing the transmission, spread and outbreak trends under experimental or real-world scenarios [201, 158]. As illustrated in Table 4, various compartmental models are available to characterize COVID-19. For example, the SIDARTHE compartmental model considers eight stages of infection: susceptible (*S*), infected (*I*), diagnosed (*D*), ailing (*A*), recognized (*R*), threatened (*T*), healed (*H*) and extinct (*E*) to predict the course of the epidemic and to plan an effective control strategy [94]. In [172], a new compartment is introduced to the classic SIR model to quantify those who are symptomatic, quarantined infected. Further, a stochastic SHARUCD model framework contains seven compartments: susceptible (*S*), severe cases prone to hospitalization (*H*), mild, sub-clinical or asymptomatic (*A*), recovered (*R*), patients admitted to intensive care units (*U*), and the recorded cumulative positive cases (*C*), which include all new positive cases for each class of *H, A, U, R*, and deceased (*D*) [5]. In addition, several models involve new compartments to represent asymptomatic features to mild symptoms [5, 290] and undocumented cases [154]. For the recent Omicron variant, Khan et al. [134] separate the infected compartment as asymptomatic individuals *I*_*α*_, symptomatic individuals *I*_*s*_, infected with Omicron variant *I*_*o*_ since people infected with Omicron may further infect other people, so it does not matter whether they have been vaccinated or not.

**Table 4.** Summary and Examples of COVID-19 Epidemiological Modeling.

| Objectives | Factors and Settings | Data | References |
| --- | --- | --- | --- |
| COVID-19 epidemic transition and spread | SIR variants like SIDARTHE with eight phases: susceptible, infected, diagnosed, ailing, recognized, threatened, healed and extinct as well as severe symptoms; variational mode decomposition with shallow regressors, etc. | Case numbers, reporting information, demographics, test results, external data, etc. | [94,68] |
| COVID-19 epidemic dynamics | Time-dependent compartment transmissions, time-varying state transition rates | Case numbers, reporting information, etc. | [58,209] |
| COVID-19 asymptomatic transmission | Asymptomatic to mild symptoms, differing mild and asymptomatic from severe infections, undocumented cases, epidemiological interventions with serological tests, age-dependent and asymptomatic settings, etc. | Case numbers, reporting information, test results, symptoms, demographics, etc. | [290,5,154,17,158,290] |
| External factor influence on COVID-19 epidemic | Age-sensitive or age-structured SIR models such as SEIRD and with social contacts, public monitoring and detection policies, ethnicity, public health conditions, social contacts, environmental factors | Case numbers, demographics, health conditions, social activities, environmental factors, etc. | [60,246,193,305,159,298,282] |
| COVID-19 NPI influence | Lockdown, social distancing, quarantine, symptomatic and quarantined infected cases, self-protection, etc. | Case numbers, NPIs, health conditions, test results, demographics, etc. | [5,58,172,209] |
| COVID-19 resurgence | Second waves, wave difference, reopening business and social activities, time-decaying immunity, easing lockdown and social distancing, age sensitivity, NPI implementation information, travel ban, and virus importation | Case numbers, multi-wave data, NPIs, and external data, etc. | [12,83,99,206,163,12,151] |
| COVID-19 herd immunity | Compartmental model for simulating 'shield immunity' in a population | Case numbers, serological tests, etc. | [290] |

##### Modeling the COVID-19 epidemic dynamics, transmission, and risk

Classic compartmental models typically assume constant transmission and recovery rates between state transitions for the past epidemic and infectious diseases. This assumption has been taken in many SIR variants tailored for COVID-19, which, however, cannot capture the COVID-19 disease characteristics in Section 3.2. To cater for COVID-19-specific characteristics, especially when mitigation measures are involved, the classic SIR [131] and SEIR models [13], which were applied to modeling other epidemics like measles and Ebola, have to be tailored for COVID-19. Since COVID-19 transmission may involve more states, especially with interventions, SIR and SEIR models have been extended by adding customized compartments such as quarantined, protected, asymptomatic and immune [94, 290, 5, 172].

Accordingly, to capture the evolving COVID-19 epidemiological attributes including time-variant infection, mortality and recovery rates, time-dependent compartmental models have been proposed. For example, a time-dependent SIR model adapts the change of infectious disease control and prevention laws as city lockdowns and traffic halt are imposed, reflected in the control parameters infection rate *β* and recovery rate *γ*, which are modeled as time-variant variables [58]. Dynamical modeling is also considered in temporal SIR models with temporal susceptible, insusceptible, exposed, infectious, quarantined, recovered and closed (or death) cases in [209]. In [305], an early-stage study of a dynamic SEIR model estimates the epidemic peak and size, and an LSTM further forecasts its trend after taking into account public monitoring and detection policies. In [97], a multistrain, stochastic, compartmental epidemic model identifies the relative transmissibility and immune escape of the Omicron variant with respect to both naturally acquired immunity and vaccines.

##### Modeling the COVID-19 resurgence and mutation

Our current understanding of the COVID-19 resurgence and mutation is still very limited. The reported British, South African, Indian, and other newly emergent mutations of coronavirus exhibit higher contagion and complexities [100, 101]. COVID-19 may indeed become another epidemic disease which remains with humans for a long time. Quantifying the virus mutation and disease resurgence conditions, forecasting and controlling their potential resurgences and future waves after lifting certain mitigation restrictions and reactivating businesses and social activities are important modeling tasks [163, 206]. Other tasks include distinguishing the epidemiological characteristics, age sensitivity, and intervention and containment measures between waves [99], comparing the epidemiological wave patterns between countries experiencing mutations and resurgences, comparing COVID-19 wave patterns with influenza wave patterns [83], predicting resurgences and mutations (e.g., by estimating the daily confirmed case growth when relaxing interstate movement, mobility and contact restrictions and social distancing by SEIR-expanded modeling), and preparing countermeasures for future waves [12].

Limited research results are available in the literature on the above broad issues. For example, a comparative analysis in [27] shows the differences in the second COVID-19 wave in Italy and the strategies taken in implementing facemasks, social distancing, business closures and reopenings. In [37], building on fitting the first wave data, an epidemic renormalisation group approach further simulates the dynamics of disease transmission and spread across European countries over weeks by modeling the European border control effects and social distancing in each country. In [151], an SIR model estimates the scenarios of incurring a potential second wave in China and the potential case fatality rate if containment measures such as travel bans and viral reintroduction from overseas importation are relaxed for certain durations in a population with a certain epidemic effect size and cumulative count after the first wave. In [206], an SEIR model incorporates social distancing to model the mechanism (closure releasing) of forming the second wave, the epidemiological conditions (ranges of transmission rate and the inverse of the average infectious duration) for triggering the second and third waves, and the socioeconomic (economic loss due to lockdown) and intervention (novel social behavior spread) factors on case numbers. In [163], a revised stochastic SEIR model estimates different resurgence scenarios reflected on infections when applying time-decaying immunity, lockdown release, or increasing the implementation of social distancing and other individual NPIs.

##### Modeling the influence of external factors including NPIs on the COVID-19 epidemic

COVID-19 epidemic dynamics reflect the time-varying states, state transition rates, and their vulnerability to contextual and external factors such as a person’s ethnicity, public health conditions, and social contacts and networking [159]. To depict the influence of external factors, compartmental models can involve relevant side information (e.g., NPIs, demographic features such as age stratification and heterogeneity, and social activities such as population mobility) in the state transitions of COVID-19 population during outbreak. For example, an age-sensitive SIR model in [60] integrates known age-interaction contact patterns into the examination of potential effects of age-heterogeneous mitigation on an epidemic in a COVID-19-like parameter regime. An age-structured SIR model involves social contact matrices and Bayesian imputation in [246]. An age-structured SEIRD model identifies no significant susceptibility difference between age groups in [193]. More discussion on modeling the NPI effect on COVID-19 transmission and the epidemic is in Section 8.1.

In addition, environmental factors, especially humidity and temperature, may also affect COVID-19 virus survival and the epidemic’s transmission [298, 282]. For example, in [68], variational mode decomposition decomposes COVID-19 case time-series into multiple components and then a Bayesian regression neural network, cubist regression, KNN, quantile random forest, and support vector regression (SVR) are combined to forecast six-day-ahead case movements by involving climatic exogenous variables.

###### Discussion

COVID-19 compartmental models excel at modeling epidemiological hypotheses, processes and factors with domain knowledge and interpretation of infectious diseases. Table 4 illustrates various applications of COVID-19 epidemic modeling. The existing models often assume constant state-space transitions, capture average behaviors and their contagion of a closed population, and are sensitive to initial states and parameters. Challenges and opportunities exist in expanding their traditional frameworks to address the specific COVID-19 complexities and challenges in Section 3. Examples are time-varying, non-IID dynamics, and complex couplings between interior and exterior factors related to COVID-19 populations, management groups, and contexts. Other important issues include understanding how vaccination and specific vaccines affect coronavirus mutation, discovering the relationships between interior and exterior factors during their interactions with the COVID-19 ecosystem, and how internal and external COVID-19 factors jointly affect resurgence and mutations.

### 7.2 COVID-19 Medical and Biomedical Analyses

COVID-19 medical and biomedical analyses reveal the intrinsic and intricate characteristics, patterns and outliernesses of SARS-CoV-2 viruses and COVID-19 diseases. A wide range of research issues have been explored, including but not limited to: COVID-19 infection diagnosis, prognosis and treatment, virology and pathogenesis analysis, potential therapeutics development (e.g., drug repurposing and vaccine development), genomic similarity analysis and sourcing, and contact tracing. In this section, we summarize the medical and biomedical modeling of COVID-19 infection diagnosis and case identification, risk and prognosis analysis, medical imaging analysis, pathological and treatment analysis, and drug development.

#### 7.2.1 COVID-19 infection diagnosis, test and case identification

COVID-19 and its coronavirus strains such as Delta and Omicron have shown high transmission and reproduction rates, high contagion, and sophisticated and unclear transmission routes. It is crucial to identify and confirm exposed cases, test infections, identify infected virus variant types, and trace their origins and contacts as early as possible so as to timely and proactively implement appropriate quarantine measures and contain their potential spread and outbreak [189]. This is particularly important during the varying incubation periods which are often asymptomatic to mildly symptomatic yet highly contagious virus variants, particularly for the increasing evolving new virus strains.

The SARS-CoV-2 diagnosis and test methods include (1) chemical and clinical methods, typically nucleic acid-based molecular diagnosis, and antibody-based serological detection; (2) medical imaging-driven analysis, such as symptom inspection from CXR and CT images; (3) clinical diagnoses and tests like respiratory signal analysis, such as on the abnormal patterns of infected lung ultrasound waves and coughing and breathing signals; and (4) other noninvasive methods such as by involving SARS-CoV-2 and its disease data and external data [54, 56].

Data-driven discovery also plays an increasingly important role in improving COVID-19 diagnosis. Due to the virus and disease complexities, alternative and complementary to the chemical and clinical diagnosis approaches, COVID-19 identification [263] can benefit by analyzing biomedical images, genomic analysis, symptom identification and discrimination, and external data including social contacts, social activities, mobility and media communications, etc. by data-driven discovery [42].

Below, we summarize and illustrate the above research for COVID-19 infection diagnosis, test and case identification.

- *Nucleic acid-based diagnosis test* (NAT) [87, 3, 307] refers to various molecular diagnosis test methods, including the non-isothermal amplification (e.g., the real-time reverse transcription polymerase chain reaction (RT-PCR) test, which is the gold standard of COVID-19 diagnosis), isothermal amplification (e.g., CRISPR-based), and sequencing-based tests. Such methods may benefit from modeling techniques including gene and protein sequence analysis and drug-target and virus-host interaction analysis. It is highly sensitive and usable for large-scale operations, but it is expensive as typically it is done using specific test materials and in labs, which is less accurate as it is subject to the varied quality and quantity of specimen collections. The challenges are to reduce its false-negative and false-positive rates supplemented by other diagnosis tools and develop scalable fast test tools.
- *Antibody-based serological diagnosis* [155, 148, 208] is to detect anti-SARS-CoV-2 immunoglobulins, i.e., the antibodies produced in response to COVID-19 infections by validating the specificity and sensitivity of chemiluminescent immunoassays, enzyme-linked immunosorbent assays and lateral flow immunoassays against SARS-CoV-2. It is an alternative or complement to NATs for acute infection diagnosis with easier and cheaper operations at any time. It, however, may produce poor-performing results which are unreliable for decision-making. It may take time to get the results and it might be unsuitable for early large-scale diagnosis. There is an urgent need to develop more accurate serological test methods and tools. Machine learning methods such as CNNs could improve test performance, e.g., analyzing the test results, involving external data on patient demographics and clinical results, and integrating various test results [175].
- *Clinical diagnosis and analysis* involves clinical reports, domain knowledge, and clinicians in identifying COVID-19-specific symptoms, indications, and infections; differentiating them from other similar diseases such as influenza; and confirming positive, negative, severe, or fatal conditions. Such diagnoses are conducted by blood tests, cough sound judgment, breathing pattern detection, and external factors by involving external data, etc. AI, machine learning and analytics methods have been increasingly used to classify COVID-19 from other diseases, predict infections, recovery and mortality rates, numbers or timing, etc. [31, 135, 33]. For further discussion, see Section 6.2.
- *Clinical medical imaging analysis* for the COVID-19 inspection on COVID-19-sensitive medical images, typically using DNN-based image analysis, can complement the aforementioned chemical and medical methods. It can detect abnormal and discriminative symptoms and patterns sensitive to COVID-19 in patient’s CXR and CT images. Both typical deep and shallow learning methods have been widely applied, which may also bring about inconsistencies and biases in their applications, experiments, results and actionability [230]. For further discussion, see Section 7.2.3.
- *Data-driven prediction* is conducted on COVID-19 related data, such as blood test results, respiratory signals, and external data that may indicate symptoms, patterns or anomalies of COVID-19 infections. Shallow and deep learning and mathematical modeling methods have been applied to classify symptom types, differentiate COVID-19 infections from other diseases, and detect outliers that may indicate COVID-19 infections. For example, in [31], a random forest classifier estimates the likely positive or negative COVID-19 patients on hematochemical values from routine blood exams. In [236], computer audition is used to recognize COVID-19 patients under different semantics such as breathing, dry and wet coughing or sneezing, and speech during colds, etc. AI4COVID-19 [120] combines the deep domain knowledge of medical experts with smart phones to record cough and sound signals as the input data to identify suspect COVID-19 infections with 92.8% accuracy reported. In [183], a shallow LSTM model combines medical information and local weather data to predict the risk level of the country.

##### Discussion

A comparison of the diagnosis methods is shown in Table 5. As commented in various reviews [268, 24, 254, 155, 179], COVID-19 diagnosis and tests still suffer from various limitations and challenges. The issues include concerns about the result quality, implementation scalability, actionability for determining isolation and quarantine strategies, and trustfulness of accepting medical findings as general clinical specifications. An increasing number of studies appear promising by incorporating advanced data science and AI techniques into complementing medical and chemical test approaches and tools. They integratively enhance pre-analytical and postanalytical test results, and strengthen the interpretability and actionability of the results for clinicians, microbiological staff, and public health authorities.

**Table 5.** Comparison of COVID-19 Infection Diagnosis Methods.

| Methods | Pros | Cons | Data | References |
| --- | --- | --- | --- | --- |
| Nucleic acid-based diagnosis test | High sensitivity, suitable for large-scale operation | Preliminary assessment by technicians, professional data analysis, expensive, less accurate, false-negative or false-positive results | Nasal, nasopharyngeal or oropharyngeal swab, aspiration, saliva or wash specimens | [307] |
| Serological diagnosis | Easy and cheap to implement, no requirement of experts | Unstable performance, time-inefficient, unscalable for early diagnosis | Serum or plasma samples | [155] |
| Clinical diagnosis analysis | Diagnosis from mixed clinical reports and tests, on-demand, verifiable by domain experts | Require professional tools and domain knowledge | Blood and respiratory test samples, etc. | [31] |
| Medical imaging inspection | Fast and automated detection, data-driven analysis | Need trained experts, costly in labeling and early detection, train data scarcity | CT and CXR images | [230] |
| Data-driven prediction | Algorithmic prediction by data-driven analytics and learning on data relevant to the COVID-19 diagnosis | Biases from data and predictors | Any relevant data including clinical test results and genomic/protein sequences | [56] |

#### 7.2.2 COVID-19 patient risk and prognosis analyses

COVID-19 patient risk assessment identifies the risk factors and parameters associated with patient infections, disease severity, and recovery or fatality to support accurate and efficient prognosis, resource planning, treatment planning, and intensive care prediction. This is crucial for early interventions before patients progress to more severe illness stages. Moreover, the risk and prognosis prediction for patients can inform effective health and medical resource allocation when intense monitoring, such as that involving ICU and ventilation, and more urgent medical interventions are needed and prioritized. Machine learning models and data-driven discovery can also play a vital role in such risk factor analyses, scoring, prediction, prioritization, and planning of prognostic and hospitalization resources and facilities, treatment, and discharge planning. They can also conduct the influence and relation analyses between COVID-19 infection, disease conditions, and the external environment and context (e.g., weather conditions and socioeconomic statuses).

Techniques including mathematical models, and shallow and deep learners are applicable on health records, medical images, and external data. For example, LightGBM and Cox proportional-hazard (CoxPH) regression models incorporate quantitative lung-lesion features and clinical parameters (e.g., age, albumin, blood oxygen saturation, and CRP) for prognosis prediction [315]. Their results show that lesion features are the most significant contributors in clinical prognosis estimation. Supervised classifiers like XGBoost are applied on electronic health records to predict the survival and mortality rates of severe COVID-19 infectious patients [302, 237] for the detection, early intervention, and potential reduction of mortality of high-risk patients. In [223], logistic regression and random forest are used to model CT radiomics on features extracted from pneumonia lesions to predict feasible and accurate COVID-19 patient hospital stay, which can be treated as one of the prognostic indicators. Further, shallow and deep machine learning methods are applied to screen COVID-19 infections on respiratory data, including lung ultrasound waves, and coughing and breathing signals. For example, in [123], a bidirectional GRU network with attention differentiates COVID-19 infections from normal on face-based videos captured by RGB-infrared sensors with 83.69% accuracy. Lastly, external data can be involved in risk analysis. For example, the work in [173] analyzes the association between weather conditions and COVID-19 confirmed cases and mortality.

#### 7.2.3 COVID-19 medical imaging analyses

COVID-19 medical services generate typical medical imaging data, including the CXR and CT images of a COVID-19 patient’s lung (lobes or segments), lesion, trachea, or bronchus. Building on shallow and deep learning methods, especially various pretrained CNN-based image nets, a rapidly growing number of research studies are available on COVID-19 medical imaging processing. They handle various COVID-19 medical imaging tasks, including their feature extraction, the region of interest (ROI) segmentation, infectious region or object detection, and disease and symptom diagnosis and classification. The most commonly used DNNs are pretrained and customized CNNs, GANs, VGGs, Inception, Xception, ResNet, DenseNet, and their variants [121, 242]. Their principles are similar to other applications in computer vision and pattern recognition.

Further, CNN-based transfer learning models and deep transfer learning are applied on CXR images to detect COVID-19 pneumonia, segment the pneumonia, and detect their severity [63, 180]. On chest CT images, CNNs like ResNet, DenseNet, VGG and the Inception transfer networks are applied to classify COVID-19 infected patients, detect COVID-19 pneumonia, and localize infection regions [318, 217, 11, 284, 249].

It is reported that the application of various DNNs for COVID-19 medical imaging analyses shows significant performance advantages in comparison with shallow learners. For example, several references report close-to-perfect prediction performance of pretrained DNNs on CXR images, such as achieving 100% accuracy and F-score 1.0 in [161], AUC 1.0 in [222] and AUC 0.9997 in [170], and 98% accuracy and F-score 0.98 in [186]). Lower performance is achieved by customized networks on CT images, e.g., obtaining 99.68% accuracy in [108], 0.994 AUC in [85], and 0.94 F-score in [85, 249]). These reported results show the highly promising potential of deep learning for COVID-19 medical imaging analytics, especially when the number of COVID-19 medical imaging is sufficient for DNN fitting.

Further, Table 6 illustrates various DNNs applied on medical imaging for COVID-19 screening and abnormal infection region segmentation, etc. For example, CNNs such as shallow CNNs, truncated InceptionNet, VGG19, MobileNet v2, Xception, ResNet18, ResNet50, SqueezeNet, DenseNet-121, COVIDX-Net, GoogleNet, AlexNet and capsule networks [69, 183, 18, 111, 2, 279] have been applied to analyze CXR images. They are used to screen COVID-19 patients, assist in their diagnosis, quarantine and treatments, and differentiate COVID-19 infections from normal, pneumonia-bacterial, and pneumonia-viral infections.

Please note that the reported results in the literature are mainly data-driven using pattern and exception analyses. They result from their specific designs, settings, data manipulation, fitting, and evaluation on their specific data. Their clinical applications should be subject to further methodological checks, validation, practical tests [230, 295], and domain-driven actionability checks [42, 45], etc.

#### 7.2.4 COVID-19 pathological and treatment analyses and drug development

The modeling of COVID-19 pathology and treatment aims to characterize virus origin and spread, infection sources, pathological findings, immune responses, and drug and vaccine development, etc. The formulation of coronavirus molecular mechanisms and pathological characteristics underlying the viral infection can inform the development of specific anti-coronavirus therapeutics and prophylactics, and disclose the structures, functions and antigenicity of SARS-CoV-2 spike glycoprotein [275]. The pathological findings can pave the way to design vaccines against coronavirus and its mutations. For example, a higher capacity of membrane fusion of SARS-CoV-2 compared with SARS-CoV is shown in [296], suggesting the fusion machinery of SARS-CoV-2 as an important target of developing coronavirus fusion inhibitors. Further, human angiotensin coverting enzyme 2 (hACE2) may be the receptor for SARS-CoV-2 [198] informing drug and vaccine development for SARS-Cov-2. In [276], a structural framework for understanding coronavirus neutralization by human antibodies can help understand the human immune response upon coronavirus infection and activate coronavirus membrane fusion. The kinetics of immune responses to mild-to-moderate COVID-19 discloses clinical and virological features [256].

Data-driven analytics have also been applied in COVID-19 virology, pathogenesis, genomics, and proteomics. They analyze the pathological testing results, gene sequences, protein sequences, physical and chemical properties of SARS-CoV-2, drug information and its effect, together with their domain knowledge. Such approaches play an important role in discovering and exploring feasible drugs and treatments, enabling drug discovery and repurposing, and correlating drugs with protein structures for COVID-19 drug selection and development. For example, a pretrained MT-DTI (molecule transformer-drug target interaction) deep learning model based on the self-attention mechanism identifies commercially available antiviral drugs by finding useful information in drug-target interaction tasks [20]. In [317], 28 machine learning methods including generative autoencoders, generative adversarial networks, genetic algorithms, and language models generate molecular structures and representations on top of generative chemistry pipelines and optimize them with reinforcement learning to design novel drug-like inhibitors of SARS-CoV-2. Further, multitask DNNs screen candidate biological products. In [176, 177], CNN-enabled CRISPR-based surveillance supports a rapid design of nucleic acid detection assays.

For genome and protein analysis, frequent sequential pattern mining identifies the frequent patterns of nucleotide bases, predicts nucleotide base(s) from their previous ones, and identifies the genome sequence locations where nucleotide bases have changed [187]. In [8], a bidirectional RNN classifies and predicts the interactions between COVID-19 non-structural proteins and between the SARS-COV-2 virus proteins and other human proteins with an accuracy of 97.76%.

Classic and deep machine learning methods such as classifiers SVM and XG-Boost, sequence analysis, multi-task learning, deep RNNs, reinforcement learning such as deep Q-learning network, and NLP models such as Transformer are applied to SARS-COV-2 therapy discovery, drug discovery, and vaccine discovery [132]. Examples are the rule-based filtering and selection of COVID-19 molecular mechanisms and targets; virtual screening of protein-based repurposed drug combinations; identifying the links between human proteins and SARS-COV-2 proteins; developing new broad-spectrum antivirals and molecular docking; identifying functional RNA structural elements; discovering vaccines such as predicting potential epitopes for SARS-COV-2 and vaccine peptides by LSTM and RNNs; and analyzing protein interactions, and molecular reactions by neural NLP models such as Transformer variants.

Table 7 briefly illustrates the applications of COVID-19 modeling in supporting COVID-19 treatments and drug and vaccine development.

**Table 7.** Summary and Examples of Modeling for COVID-19 Treatment and Drug and Vaccine Development.

| Objectives | Approaches | Data | References |
| --- | --- | --- | --- |
| Treatment | Data-driven diagnosis-informed treatment, e.g., pathological analysis, medical imaging analysis, immune reaction, genomic and proteomic analysis | Pathological, clinical, virological, genomic, proteomic data | [121, 315, 121, 242] |
| Drug development | Correlating drugs with protein structures and molecule transformer for drug-target interactions, DNNs like GANs and multitask DNNs for drug discovery, machine learning and language models to generate molecular structures and drug-like inhibitors | Virological, genomic, proteomic data | [20, 317] |
| Vaccine development | Sequence analysis and sequential modeling like LSTM and RNN variants and NLP models like Transformer variants for functional RNA structures, vaccine epitopes and peptides, protein interactions and molecular reactions | Genomic and proteomic data | [132] |

##### Discussion

In this review, we illustrate some results of COVID-19 medical and biomedical analytics, which have been published by peer-reviewed venues, associated with advanced methods, or achieving impressive performance. We did not validate them independently, nor check their design, implementation and evaluation. We also did not check their reproducibility and actionability. They may not be directly applicable to practice. It would not be surprising if their models and results are not reproducible and nonactionable when deployed. In fact, as concerned in domain-driven actionable knowledge discovery [45], data and AI ethics [126], and broadly in fairness, bias, and code of conduct of applying AI models in practice [174, 73], it is often the case that data-driven and machine learning findings in the literature are not actionable for decision-making. This may be due to various reasons, such as their over-focus on data and parameter fitting, ignorance of COVID-19 characteristics and complexities, and mismatch of pretrained deep networks with the COVID-19 reality. There may also be gaps in characterizing COVID-19 characteristics and involving COVID-19 knowledge into modeling. In particular, deep neural networks tend to report close-to-perfect results on simple data, which may not be the case when they are applied on usually small, highly inconsistent, and quality-limited COVID-19 data.

It is thus worthy of noting that the reported results should not be directly used as evidence of implementing the corresponding COVID-19 case confirmation, medical treatment, hospitalization, resource planning, and quarantine, etc. Their practical test and deployment require the involvement of medical and clinical context, knowledge, expertise, code of conduct, and regulation. Independent and external validation are also suggested to further verify the soundness, applicability, and actionability [45, 42] of their methodologies, processes, models, data manipulation, business and technical evaluation, and result conditions. It is also suggested to undertake a careful check of potential methodological flaws, design biases, learning and evaluation fairness, and faulty manipulation of features and labels, which could cause their uselessness in practice [230, 295].

## 8 COVID-19 Influence and Impact Modeling

COVID-19 has had an unprecedented and overwhelming influence and impact on all aspects of our life, society, and the economy. It has posed significant health, economic, environmental and social challenges to the entire world and all mankind [48, 104, 1, 136]. The literature analysis in [44] reports that over 3k references of the 44k modeling references involve the topic of COVID-19 influence and impact modeling. Here, we review and summarize the modeling and analysis methods and illustrate some results on several broad and important areas affected by SARS-CoV-2 and COVID-19. They include modeling the effect of COVID-19-sensitive NPIs and the COVID-19 healthcare, psychological, economic, and social influence and impact. Table 8 further summarizes and illustrates various areas and their subareas influenced by the COVID-19 pandemic and their relevant research.

**Table 8.** Summary and Examples of COVID-19 Influence and Impact Modeling.

| Aspects | Objectives | Approaches | Data | References |
| --- | --- | --- | --- | --- |
| NPI effect & influence | on epidemic dynamics | SIR model variants, statistical models, Bayesian hierarchical models, etc. | COVID-19 case data, case reporting data, NPIs | [257, 71, 30, 209, 144, 88, 219] |
|  | on public resources | SIR model variants, statistical models, polynomial regressors, etc. | Case data, NPIs, public resource (incl. healthcare) data | [84, 94, 5] |
|  | on human behaviors | SIR model variants, statistical models e.g. MCMC, relation modeling, behavior and event analysis, etc. | Case data, NPIs, human activities (incl. mobile phone data), mobility, etc. | [140, 130, 92, 98, 290, 314, 86, 146, 77] |
| Psychological and mental influence | on individual mental health | Psychology, systematic reviews, classic and neural NLP models e.g. BOW, LDA, SciBERT, Transformer variants, etc. | Clinical data, personal healthcare records, identity, social media data, news feeds, Q/A, surveys, instant messaging, behaviors, NPIs, etc. | [299, 286, 188, 267] |
|  | on public mental health | Psychology, systematic reviews, classic and neural NLP models e.g. BOW, LDA, SciBERT, Transformer variants, etc. | Healthcare data, social media data, news feeds, Q/A, surveys, instant messaging, public emotion, activities and events, NPIs, etc. | [299, 286, 188, 270, 114] |
|  | on disadvantaged and vulnerable people's mental health | Psychology, systematic reviews, meta-analyses, classic and neural NLP models, statistics, etc. | Healthcare data, disadvantaged and vulnerable people information, social media data, questionnaires, instant messaging, behavior and events, NPIs, etc. | [299, 106] |
| Economic and financial impact | on economic growth | Time-series analysis, descriptive analytics, macroeconomic models, relational models, etc. | Economic data, financial data, case data, NPIs, etc. | [48, 214, 274, 26] |
|  | on work-force and sustainability | Descriptive analytics, time-series analysis, relational models, NLP models, Q/A analysis, etc. | Work and sustainability-related data, performance, activities, unemployment, surveys, questionnaires, social media data, social welfare data, etc. | [15, 89, 153] |
| Social, public and political impact | on human behaviors | Descriptive analytics, pattern analysis, sequence analysis, behavior informatics, social media and network analysis, NLP models, etc. | Public, online, and household activities, gathering, mobility, entertaining, mobile phone data, social media data, etc. | [118, 98, 114] |
|  | on public health systems | Descriptive analytics, relational models, machine learning models, etc. | Public health and medical data, healthcare data, public health resources, public hygiene data, case data, A/Q, surveys, etc. | [80, 271] |
|  | on misinformation and information disorder | Classifiers, classic and neural NLP models, social media and network analysis, sentiment and topic modeling, time-series analysis, outlier detection, etc. | Social media data, news feeds, Q/A, cross references, fact-check, etc. | [19, 62, 4, 93, 150, 178, 243] |
|  | on socio-political systems | Descriptive analytics, sociopolitical methods, survey analysis, etc. | Social and political data, case data, surveys, questionnaires, sociopolitical events, etc. | [141, 244] |

### 8.1 Modeling COVID-19 Intervention and Policy Effects

On one hand, pharmaceutical measures and drug and vaccine development play a fundamental role in containing COVID-19 [320]. On the other, governments adopt various NPIs, such as travel restrictions, border control, business and school shutdown, public and private gathering restrictions, mask-wearing, and social distancing, to control outbreaks of COVID-19 and its further influence on various aspects. For example, travel bans and lockdowns are issued to decrease cross-border population movement; social distancing and shutdowns minimize contacts and community spread; school closures and teleworking reduce indoor gatherings and workplace infections. Although these control measures flatten the curve, they also undoubtedly change the regular mobility and activities of the population, normal business and economic operations, and the usual practices of our daily businesses.

A critical modeling issue is to characterize, estimate and predict how such NPIs influence the COVID-19 epidemic dynamics, infection spread, case development, population structure including deceased people, medical resource and treatment allocation, and human, economic and business activities. Accordingly, various modeling tasks apply epidemiological, statistical and social science modeling methods and their hybridization (e.g., stochastic compartmental models) to evaluate and estimate their effects. Typically, such modeling aligns NPIs with the case numbers of a country or region for their correlation and dependency modeling. Below, we summarize and illustrate the modeling of NPI influence on COVID-10 epidemic dynamics, public resource allocation including healthcare systems, and human activities.

#### Modeling the NPI influence on COVID-19 epidemic dynamics

This typically models the correlations between COVID-19 cases and NPIs, the NPI influence on COVID-19 epidemic factors including transmission rate and case numbers, and the NPI influence on improving recovery rates and lowering death rates. Various SIR and statistical models have been applied to evaluate the effects of such control measures and the NPI combinations on containing virus spread and controlling infection transmission (e.g., per the transmission rate) and to estimate the corresponding scenarios (distributions) of case number development [257, 71, 30]. For example, in [209], a generalized SEIR model involves the self-protection and quarantine measures to interpret the publicly released case numbers and forecast their trend in China. In [257], the effect of control measures, including city lockdowns and travel bans implemented in the first 50 days in Wuhan, and their effect on controlling the COVID-19 outbreak across China in terms of infection case numbers were estimated by an SEIR model before and after the controls.

Often, various NPIs are jointly implemented to contain a COVID-19 epidemic. It is thus essential to estimate how multiple NPIs cooperatively reduce the epidemic effective reproduction number [30, 144, 88, 219]. In [30], a temporal Bayesian hierarchical model incorporates auxiliary variables describing the temporal implementation of NPIs to infer the effectiveness of individually (estimated 13% to 42% reduction of reproduction number) and conjunctionally (77% reduction of reproduction number) implementing NPIs such as staying-at-home, business closures, shutting down educational institutions, and limiting gathering sizes in terms of their influence on the reproduction number. In [88], a hierarchical Bayesian model infers the impact and effectiveness of NPIs (including case isolation, school closure, mass gathering ban, and social distancing) on the infections, reproduction number *R*_0_, effect sizes of population, and the death toll in 11 European countries and suggests continued interventions to keep the epidemic under control.

#### Modeling the NPI influence on public resources including healthcare systems

The implementation of NPIs affects the demand, priority, and effectiveness of anti-pandemic public health resourcing and the planning and operations of healthcare systems. For example, in [84], an SEIR model and a polynomial regressor simulate the effect of early detection, isolation, treatment, adequate medical supplies, hospitalization, and therapeutic strategy on COVID-19 transmission, in addition to estimating the reproductive number and confirmed case dynamics. The SIDARTHE model [94] simulates the possible scenarios and the necessity of implementing countermeasures such as lockdowns and social distancing together with population-wide testing and contact tracing to rapidly control the pandemic. The SHARUCD model [5] predicts the COVID-19 transmission response (in terms of infection cases, growth rate, and reproduction number) to the control measures including partial lockdown, social distancing, and home quarantining and differentiates asymptomatic and mild-symptomatic from severe infections. The results are assumed useful for informing the prioritization of healthcare supplies and resources.

#### Modeling the NPI influence on human activities

This explores the relations between COVID-19 NPIs and human mobility, travel, social and online activities. For example, in [140], the alignment between human mobility and case number development in Wuhan and China presents the effect of travel restrictions on case reduction and COVID-19 spread. In [130], a simple SEIR model analyzes contact tracing in the UK’s social network data, estimates the scenarios of COVID-19 infection control and subsequent untraced cases and infections, and shows the efficacy of close contact tracing in identifying secondary infections. In [92], the MCMC parameter estimation and a meta-community SEIR-like disease transmission model show the need for planning emergency containment measures such as restrictions on human mobility and interactions to control COVID-19 outbreak (by 42% to 49% transmission reduction). In [98], mobile phone data was collected and analyzed to inform epidemiologically relevant COVID-19 behaviors and responses to interventions. Weitz et al. [290] developed and analyzed an epidemiological intervention model that leverages serological tests to identify and deploy recovered individuals as focal points for sustaining safer interactions by interaction substitution, developing the so-called ‘shield immunity’ at the population scale. In addition, it is shown that the change of contact patterns could dramatically decrease the probability of infections and reduce the transmission rate of COVID-19 [314, 86, 146].

##### Discussion

The diverse SIR-based modeling of COVID-19 invention and policy effects enables an epidemiological explanation. Such methods assume each NPI independently acts on the case movement. This leaves various gaps and open issues. One is to characterize the effectiveness of individual NPIs by assuming they are coupled with each other and cooperatively contribute to flatten the curves. The other is to explore the interactions between NPIs, case development, and external factors such as people’s behaviors and environmental factors without disentangling them (opposite to the method of DNN-based decoupled, homogeneous, and independent representations and learning).

### 8.2 Modeling COVID-19 Psychological and Mental Impact

The literature analysis shows that the influence of COVID-19 on individual and public psychological and mental health is the most commonly investigated impact of concern [44, 299]. To quantify such impact, typical tasks are to characterize, classify and predict social-media-based individual and public emotion, sentiment and their mental health issues. The COVID-19 psychological and mental impact modeling may involve data and aspects of COVID-19 outbreak, health and medical mitigation, NPI measures, government governance, public healthcare system performance, vaccines, resurgence, coronavirus mutations, and the ‘new normal’ situations including working from home, home schooling, and online education. Other commonly used data and explored tasks are from social media and networks such as Twitter, Facebook, Wechat, Weibo, YouTube, Instagram, and Reddit; online news feeds, discussion boards, blogs and Q/A; and instant messaging such as mobile messaging and apps.

The COVID-19 psychological and mental impact analyses, characterizes, clusters, classifies or identifies negative sentiments [286, 188], opinion and topic trends, online hate speech [270], psychological stress, men’s and women’s worries [267], responsive emotions [114], behaviors and events on short and long texts by applying NLP and neural language modeling techniques. Examples are extracting TF-IDF and part-of-speech features and then applying NLP and text analysis models. Typical models include shallow NLP and text analysis models such as bag-of-words and latent Dirichlet allocation (LDA), and neural text modelers including BioBERT, SciBERT, Transformer and their variants on the word, sentence or corpus level. For example, in [299], the preferred reporting items for systematic reviews and meta-analyses guidelines are used to review the COVID-19 impact on public mental health, disclosing the extent of symptoms and risk factors associated with anxiety (6.33% to 50.9%), depression (14.6% to 48.3%), posttraumatic stress disorder (7% to 53.8%), psychological distress (34.43% to 38%), and stress (8.1% to 81.9%) in the surveyed population of eight countries.

#### Discussion

The existing modeling of COVID-sensitive psychological and mental influence often misses psychological knowledge because it is purely driven by data. The analytical results are based on a cohort of infected people with anonymous identification. No work is available on fusing various sources of data including online misinformation to infer the predominant drivers of specific mental stress such as vaccination hesitation and the targeted analysis of specific mental issues in vulnerable groups such as COVID-driven teenage suicide and racism.

### 8.3 Modeling COVID-19 Economic Impact

The COVID-19 pandemic has had an overwhelming and devastating impact on regional and global economy and business activities, including trade, tourism, education exchange, logistics, supply chains, workforce, and employment. It seems that no economy on the interconnected globe is immune to the negative consequences of COVID-19 [61, 129, 21, 26]. Typical tasks of modeling COVID-19 economic impact include identifying and quantifying indications and insights for (1) how COVID-19 influences various aspects of the economy and businesses; (2) how to manage and balance COVID-19 control measures (including NPIs and vaccination rollouts) and government relief and recovery programs, and (3) how to sustain and recover business and economic activities without seriously suffering from uncontrollable outbreaks and resurgences for better sustainability in the COVID new normal. Below, we illustrate two sets of impact on economic growth, the workforce, and sustainability.

#### Modeling the COVID-19 impact on economic growth

A rapidly growing body of research investigates the heterogeneous, non-linear and uncertain macroeconomic effects of COVID-19 across regions and sectors in individual countries and on a global scale. It is estimated that COVID-19 and SARS-CoV-2 may have caused over 2% monthly GDP loss and a 50% to 70% decline in tourism in 2020 [48]. In [214], a sectoral macroeconomic model analyzes the short-term effects of intervention measures, such as lockdown, social distancing, and business reopening, on economic outcomes. Examples of economic outcomes include affecting the production network, supply and demand, inventory dynamics, unemployment, and consumption and estimating their influence on the relations between reproduction number and GDP. The study in [274] illustrates the relations between a country’s income levels, public healthcare availability and capacity, and the COVID-19 infected patient’s demography and social patterns in low-to middle-income countries.

#### Modeling the COVID-19 impact on workforce and sustainability

COVID-19 drives the new normal of work practices, including a hybrid work mode, cloud-based enterprise operations, the shift from centralized infrastructures (including IT) to cloud-based ICT and home-based workplaces, and gender inequality in unemployment. COVID-19 also triggers new ways of ensuring sustainability, including engaging and supporting clients through online operations and services and AI-enabled cost-effective planning, production, logistics and services. In [15], the descriptive statistics of the daily activities of Baidu developers show the positive and negative impacts of working from home on developer productivity, particularly on large and collaborative projects. The meta-analysis and review in [89] estimate the health, social, and economic effects of the COVID-19 pandemic on gender equality and identify that women more likely suffered from unemployment at the early stage of the COVID-19 pandemic. The analysis in [153] in Australia shows the impact of government welfare support responses to COVID-19-infected people and businesses on mitigating potential unemployment, poverty, income inequality, and the sustainability of such support measures.

##### Discussion

The existing objectives, tasks and methods of modeling COVID-19 economic impact are highly preliminary, specific and limited. Expected opportunities include macro-, meso- and micro-level modeling of economic impact by involving their economic-financial variables and activities, contrastive analysis with similar historical events and periods, and data-driven insight discovery for a sustainable tradeoff between mitigation and economic growth in the new normal, to name a few.

### 8.4 Modeling COVID-19 Social Impact

The COVID-19 pandemic has had a significant impact on public health, welfare, and social, political and cultural systems. It has reshaped our global society, including restricting human activities and mobility, affecting people’s well-being, causing an overwhelming burden on public health systems, restructuring sociopolitical systems, and disturbing social regularity and order such as incurring online information disorder. This section reviews the relevant modeling work on such social impacts.

#### Modeling the COVID-19 influence on human behaviors

In addition to the COVID-19-sensitive NPI influence on human activities as discussed in Section 8.1, SARS-CoV-2 and COVID-19 have fundamentally reshaped people’s social activities, mobility, and habits [66, 303]. For example, Baidu-based daily transportation behaviors and simple statistics show COVID-specific high-level mobility patterns such as visiting venues, origins, destinations, distances, and transport time during the COVID-19 epidemic in China [118]. In [98], large-scale mobile phone data such as call detail records, GPS locations, Bluetooth data, and contact tracing apps were collected and analyzed by off-the-shelf tools to extract statistic metrics and patterns of behaviors, mobility, and interactions. Their results indicate population behaviors, individual contacts, movement paths, mobility patterns, and networking affected by the COVID-19 epidemic. In [114], social media data from Sina Weibo, the Baidu search engine, and 29 Ali e-commerce marketplaces were collected and analyzed using keyword-based linguistic inquiries and statistics like word frequencies and Spearman’s rank correlation coefficient analysis. Keywords extracted show people’s behavioral responses to COVID-19 outbreaks, public awareness and attention to COVID-19 protection measures, concerns about misinformation and rumors about ineffective treatments, and the correlation between risk perception and negative emotions.

#### Modeling the COVID-19 influence on public health systems

The sudden COVID-19 endemic to pandemic and its mysterious, wide and unexpected resurgences have resulted in significant impact on public health systems globally [101, 297, 251]. They increase the imperative, nonscheduled and overwhelming rationing demand on healthcare and medical professionals, public health and medical resources and supplies. The pandemic has caused a global shortage of health resources, including oxygen, hospital beds, hospital facilities, ICU facilities, ventilators, medical waste processing equipment, hygiene protection equipment such as medical masks and sanitization chemicals, and intervention materials and devices. The pandemic also triggers the research demand on how to better plan, prioritize, ration, and manage these resources, assess their supply and demand, and evaluate global supply chain systems. Other research studies the effects of different strategies and priorities in particular of prioritizing the emergent resources for COVID hotspots and in-demand regions and optimizing their reorganization per local and global needs. Another important topic is to assess and improve the population-wide well-being during the pandemic by identifying disadvantaged groups, aged and vulnerable people, people and communities hit hard by the pandemic, and those experiencing poverty. For example, in [80], recommendations are made to allocate medical resources to both COVID-19 and non-COVID-19 patients to maximize benefits, prioritize health workers, avoid a first-come, first-served approach, in a way that is evidence-based and involves science and research findings. People from different classes have different affordabilities and capacities, and the COVID-19 impact on them also presents different patterns and extents, including on vaccine production, affordability, allocation, and deployment [292] and on people’s vaccination intention [271].

#### Modeling the COVID-19 influence on sociopolitical systems

The COVID-19 influence on social and political systems is unprecedented. COVID-19 has reshaped confidence and trust in the existing sociopolitical systems, including public and moral values, national interests, social welfare systems, human services, political relations, globalization, scientific exchange and collaborations, science-driven epidemic mitigation policies and strategies, and social governance and disaster management. For example, an identity fusion theory-based online sampling and a moral foundations theory-based computer simulation show the correlations between nationalism, religiosity, and anti-immigrant sentiment from a socio-cognitive perspective during the COVID-19 pandemic in Europe. The surveys undertaken in [141] show that COVID-19 affects the scientific uncertainty and the public and political trust in science-based policy making in the US and suggest more careful scientific communications. The work in [244] evaluates the impact of COVID-19 on globalization and global health. It shows that the COVID-19 pandemic has affected mobility, trade, travel, event management, food, and agriculture. A pandemic vulnerability index quantitatively measures the potential impact on global health and those countries most impacted.

#### Modeling misinformation and disorder in the COVID-19 infodemic

The COVID-19 infodemic has been accompanied by a large volume of misinformation (partially or entirely inaccurate or misleading information), biased (polarized), questionable, and unverified information, rumor, and propaganda [196, 93, 243]. Such information is harmful for correctly understanding, recognizing, intervening, and preventing the COVID-19 pandemic. However, their diffusion is usually fast, their spread is often wide, and their impact is typically devastating. Modeling such COVID-19 misinformation and information disorder is thus imperative. COVID-19 infodemic modeling tasks include detecting and ranking misinformation, classifying them, undertaking fact checks and cross-references, tracing their sources and transmission paths, discovering their diffusion and propagation networks and paths, and estimating their effects on the COVID-19 epidemic spread and control.

For example, in [62], skip-gram was used to represent the words collected from Twitter, Instagram, YouTube, Reddit, and Gab. The converted vector representations were then clustered by partitioning them around medoids and cosine distance-based similarity analysis to extract the topics of concern. An SIR model was then applied to estimate the basic reproduction number of the social media-based COVID-19 infodemic. A comparative analysis then estimates and compares the platform-dependent interaction patterns, information spread (w.r.t. reproduction rate), questionable and reliable information sourcing and differentiation, and rumor amplification across the above platforms. In [4], bivariate (ANOVA) and multivariate logistic regression identifies similar belief profiles of political orientation, religious commitment, and trust in science in survey-based narratives and compares the profiles of those who are disinformed or conspiratorial with scientific narratives. Further, the statistics on Weibo tweets show the COVID-19 misinformation evolution related to topics and events such as city lockdowns, cures, preventive measures, school reopening, foreign countries, the bias involving cures and preventive methods, and sentiment evolution such as fear of specific topics [150]. The work in [178] applied SVM, logistic regression, and BERT to classify COVID-19 misinformation and counter-misinformation tweets, characterizes the type, spread and textual properties of counter-misinformation, and extracts the user characteristics of the citizens involved. Further, in [19], the authors applied word embeddings and time-series analyses to analyze the spread of diverse conspiracy theories with real-life impact on both individual and group levels.

##### Discussion

Typical research on COVID-19 influence and impact modeling only involves local and regional COVID-19 case data and their affected objects. Hence, the resultant conclusions are often limited in their ability to indicate their applicability to general practices and broad pandemic control. More robust results are expected to inform medical and public health policy-making on medication, reinhabitating and reactivating businesses and society, and reboosting the economy. In addition, no-to-rare outcomes are available on how NPIs influence the threshold and effects of COVID-19 vaccinations and herd immunity and on how to balance NPIs and economic and social revivification. It is also difficult to find actionable evidence and guidelines on what policies should be implemented and what tradeoff is appropriate in balancing a COVID-19 outbreak, containing their resurgence, and promoting economic and social business recovery.

## 9 COVID-19 Simulation Modeling

Simulation has been widely applied in understanding, intervening, and managing SARS-CoV-2 and COVID-19 and their impact [67, 165]. Here, we summarize and illustrate the broad COVID-19 simulation research, simulating the COVID-19 epidemic transmission and evolution, and simulating the effect of interventions and policies on COVID-19 epidemic development. Table 9 summarizes and illustrates COVID-19 simulation modeling.

**Table 9.** Summary and Examples of COVID-19 Simulation Modeling.

| Objectives | Approaches | Data | References |
| --- | --- | --- | --- |
| COVID-19 epidemic and transmission | Markov chain model, Monte Carlo model, agent-based simulation, stochastic SIR models, mathematical models, Bayesian models, etc. | Case data, reporting information, demographics, mobility, location, etc. | [300, 7, 321, 95] |
| COVID-19 influence and impact | Monte Carlo simulation, statistical models, nonlinear analysis, machine learning methods, agent-based modeling, etc. | Case data, reporting information, business data, economic data, social activities, social media data, news, etc. | [67, 306] |
| External factor impact on COVID-19 | Mathematical models such as regression, Bayesian models, machine learning models, deep neural networks, behavior models, interaction modeling, discrete event modeling, etc. | NPIs, vaccination, drug and medical treatment data, case data, etc. | [52, 49, 76, 55, 216] |
| Treatment and intervention effect | Risk factor analysis, molecular dynamics simulation for virus-drug interactions and drug-target interactions, what-if analysis, agent-based modeling, Monte Carlo simulation, and hybrid methods, etc. | Gene sequences, molecular data, viral response, NPIs, vaccines, drugs, clinical data, case data, etc. | [149, 165, 90, 52, 57] |

### Broad COVID-19 simulation research

Despite being a small focus in COVID-19 modeling (over 3k of the 44k publications on modeling), simulation has been an essential means to understand, imitate, replicate and test SARS-CoV-2 and COVID-19 and their transmission, containment, and influence [149]. The COVID-19 simulation research can be categorized into simulating (1) the transmission and infection of SARS-CoV-2 and its COVID-19, including their infection mechanisms, epidemic transmission processes, evolution, and mutation under certain conditions, such as airborne transmission, asymptomatic transmission, indoor transport and exposure; (2) the influence and effect of external factors on SARS-CoV-2 and COVID-19, such as NPIs, the interactions and self-organization between COVID and external factors, and the effect of mitigation measures and various interior and contextual factors: (3) the virus-drug and virus-vaccine interactions, the effect of drugs and vaccines on SARS-CoV-2 and COVID-19, immune response, herd immunity, such as by molecular dynamics; and (4) the influence and impact of SARS-CoV-2 and COVID-19, such as on healthcare resource allocation, public health resource planning and optimization, vaccine rollout, production and distribution, unemployment, and the global economy.

Typical simulation methods include dynamic systems, state-space modeling, discrete event simulation, agent-based modeling, reinforcement learning, Monte Carlo simulation, molecular dynamics, evolutionary computing such as swarm intelligence, what-if analysis, Gompertz model, nonlinear dynamics such as the reaction–diffusion model and fractional-order model, and hybrid simulation [67, 165, 300, 7].

### Simulating COVID-19 epidemic transmission and evolution

One important but unclear question is how SARS-CoV-2 and COVID-19 evolved over time in the community, indoors, by airborne methods, and in contained environments. Typical simulation models include SIR model variants, statistical and mathematical models, agent-based simulation, and Monte Carlo simulation. For example, what-if analyses can be applied to estimate COVID-19 infection case numbers and their evolution under various hypotheses tests [321]. In [90], composite Monte Carlo simulation enables the what-if analysis of future COVID-19 epidemic development possibilities on top of the estimation made by a polynomial neural network on COVID-19 cases. Fuzzy rule induction then generates decision rules to inform epidemic growth and control. In [95], an agent-based simulation system simulates a COVID-19 patient’s demographic, mobility, and infectious disease state (susceptible, exposed, seriously infected, critically infected, recovered, immune, and dead), and their dynamic interactions (agents, i.e., people in epidemiology) in certain environments (home, public transport stations, and other places of interest). They further evaluate the effect of adjusting individual and social distancing (separation) on epidemics (e.g., numbers of each state).

### Simulating the relevant policy effect on the COVID-19 epidemic

Another important task is to simulate how interventions, interior and external factors, and other policies and control measures of interest influence the dynamics and their containment of the COVID-19 epidemic. For example, a discrete-time and stochastic agent-based simulation system (Australian Census-based Epidemic Model) [52] incorporates 24 million software agents. Each agent mimics an Australian individual in terms of their demographics, occupation, immunity and susceptibility to COVID-19, contact rates in their social contexts, interactions, commuting and mobility patterns, and other aspects, which are informed by the census data from the Australian Government. The system evaluates various scenarios by adjusting the level of restrictions on case isolation, home quarantine, international air travel, social distancing, school closures, and their effects on COVID-19 pandemic consequences in terms of the virus reproductive number, the generation period, the growth rate of cumulative cases, and the infection rate for children. The simulation provides evidence and an indication to help the government understand how COVID-19 is transmitted and what policies should be implemented to control COVID-19 in Australia.

A typical simulation tool is to introduce control-measure-sensitive variables into SIR models to estimate their effects on virus infections, reproduction numbers, transmission rate, and outbreak control after implementing or relaxing certain interventions. For example, in [172], an SIR model is implemented which incorporates variables reflecting symptomatic infections and the quarantine of susceptibles, which then estimates the case development distribution as subexponential after implementing the quarantine. In [306], an attributed heterogeneous information network incorporates the representations of external information about COVID-19 disease features, the population’s demographic features, the mobility and public perception of sentiment into a GAN model, which then assesses the hierarchical community-level risks of COVID-19 to inform interventions and minimize disruptions.

#### Discussion

Many aspects and questions related to COVID-19 could be (better) addressed by simulation, while the relevant research is still very limited. In addition to the major effort made to understand the COVID-19 epidemic dynamics, other tasks and opportunities include simulating the mutation and resurgence of the coronavirus and COVID-19 in communities with different social, ethnic, and economic conditions; the influence of individual and compound COVID-19-sensitive policies on social, economic, and psychological aspects; and the tradeoff between the strength and width of mitigation strategies and their impact on society and the economy.

## 10 COVID-19 Hybrid Modeling

Hybrid modeling dominates the literature on COVID-19 modeling. It involves various hybrid scenarios, such as multi-objective modeling, multi-task modeling, multisource and multimodal data modeling, and multi-method modeling. Hybrid COVID-19 modeling applies cross-disciplinary, cross-domain, cross-scenario, and cross-data approaches and settings. Below, we summarize and illustrate the broad-reaching COVID-19 hybrid modeling, multi-objective and multi-task modeling, multisource and multimodal data modeling, and hybrid methods for COVID-19 modeling. Table 10 summarizes and illustrates COVID-19 hybrid modeling.

**Table 10.** Summary and Examples of COVID-19 Hybrid Modeling.

| Objectives | Approaches | Data | References |
| --- | --- | --- | --- |
| Hybrid objective modeling | Multivariate analysis, probabilistic compartmental models, Bayesian SIR models, ensemble learning, deep neural networks, evolutionary computing, etc. | Case data, NPIs, external factors, demographics, clinical data, vaccinations, drug and medical treatment, etc. | [321,139,113,281,154] |
| Hybrid task modeling | Multivariate analysis, probabilistic compartmental models, Bayesian SIR models, ensemble learning, deep neural networks, transfer learning, multi-task learning, etc. | Case data, NPIs, external factors, demographics, clinical data, vaccinations, drug and medical treatment, etc. | [158,71,53,104,233,136,271,299,285] |
| Hybrid data modeling | Probabilistic compartmental models, Bayesian SIR models, multimodal analysis, event and behavior analysis, ensemble learning, neural compartmental models, deep neural networks, etc. | Case data, NPIs, external factors, demographics, clinical data, vaccinations, drug and medical treatment, mobility, business activities, news, social media, Q/A, surveys, etc. | [127,119,173,259,2,77,140,118,98] |
| Hybrid method modeling | Ensemble learning, fuzzy neural networks, probabilistic compartmental models, Bayesian SIR models, hybrid deep neural networks, etc. | Case data, NPIs, external factors, demographics, clinical data, vaccinations, drug and medical treatment, etc. | [252,310,305,158,1,49,255,103] |

### Broad COVID-19 hybrid modelling

Hybrid COVID-19 modeling can be categorized into the following families: (1) multi-objective modeling: to address multiple problems and multiple modeling objectives at the same time, such as jointly understanding the COVID-19 epidemic dynamics and their corresponding effective NPI policies; (2) multi-task modeling: to handle multiple modeling tasks, e.g., simultaneously forecasting daily confirmed, death and recovered case numbers; (3) multisource and multimodal data modeling: to involve multiple sources and modalities of internal and external COVID-19 data for modeling, e.g., supplementing environmental and demographic data with case numbers and complementing case numbers with medical imaging and social mobility data for COVID-19 infection identification and diagnosis; (4) hybrid methods for COVID-19 modeling: typically by sequentializing (i.e., multi-phase) or parallelizing multiple tasks or methods from different disciplines and areas, e.g., integrating statistical methods, shallow or deep learning methods, and evolutionary computing methods into compartmental models; and (5) complex modeling by hybridizing multi-methods from various disciplines for multi-objective or multi-task modeling on multisource or multimodal COVID-19 data.

COVID-19 hybrid modeling involves various hybrid methods across different disciplines and research areas. Examples are combining epidemic models (e.g., SIR models) with statistical models to form statistical compartmental models; integrating simulation methods (e.g., agent-based modeling) with SIR models to simulate COVID-19 infections and transmission; integrating multiple data-driven models, e.g., ensemble learning by integrating various machine learning models or deep neural networks; integrating evolutionary computing (e.g., fuzzy rules) with machine learning methods (e.g., ANN); and integrating deep learning with transfer learning.

*COVID-19 multi-objective and multi-task modeling* is commonly seen in COVID-19 modeling, as shown in Sections 5-9. Multiple COVID-19 problems and modeling objectives are involved in one task or setting. Typical problems and scenarios include forecasting COVID-19 transmission and its sensitivity to external factors, such as patients’ age groups, hygiene habits, and environmental factors; modeling the influence of NPIs and people’s ethnic conditions on case movements; modeling the influence of NPIs on both case trends and public psychological health; and estimating the survival and mortality rates and their relations to dependent factors such as patients’ health conditions.

Typical methods include multivariate analysis, probabilistic compartmental models, simulation systems, multi-objective evolutionary learning methods, and DNN variants. For example, in [216], a regression model estimates the relations between reproduction number, environment factors, and human movements. In [71], the Bayesian inference of an SIR model infers the effect of various interventions on new infections. In [206], an SEIR models the relations between case trends, epidemic conditions, socioeconomic effect, and interventions. In [299], systematic reviews and meta-analyses review the work on the relations between COVID-19 symptom severity, risk factors, and public emotions. In addition, in [158], a Susceptible-Undocumented infected-Documented infected-Recovered (SUDR) model integrates the probabilistic density function and Bayesian inference to characterize undocumented infections during unknown transmission processes with temporal transmission and social reinforcement during the COVID-19 contagion.

*COVID-19 multisource and multimodal data modeling* can serve various purposes for COVID-19 [75]. Typical applications include predicting the COVID-19 epidemic spread, transmission, medical diagnosis, treatment, government and community interventions by combining data from respective modalities or sources. Examples of multisource and multimodal data are combining COVID-19 case numbers with NPI data; integrating people’s demographics, health conditions, mobility, social and business activities with their social networking and media information; integrating health and medical records, diagnosis information, treatments, pharmaceutical interventions, and pathological tests; integrating medical imaging such as CXR, CT and ultrasound for diagnosis; combining social and public activities, events, economic data, and sociopolitical data; and integrating online, social media, mobile app-based messaging, news, Q/A, and discussion groups.

Typical methods include data fusion-based learning, mixed representation-based learning, and integrating clustering and classification on mixed data types, integrating DNN variants [127]. For example, a novel variational-LSTM autoen-coder model in [119] predicts the coronavirus spread in various countries by integrating historical confirmed case numbers with urban factors about location, urban population, population density, and fertility rate and governmental measures and responses about school, workplace and public transport closures, public event cancellation, contact tracing, public information campaigns, international travel controls, fiscal measures, and investment in health care and vaccines. In [173], COVID-19 case numbers and weather data are combined to analyze the correlation between COVID-19 confirmed cases, mortality, and weather factors. NLP methods are applied to extract and analyze the related news, which are then input to LSTM networks to update the infection rate in a susceptible–infected epidemic model [319], outperforming the susceptible–infected epidemic model and its combination with LSTM. Other examples are to couple LSTM with SIR epidemic models to forecast the COVID-19 spread using case data, population density, and mobility.

*Hybrid methods for COVID-19 modeling* integrate various methods for single or multiple-objective, multi-task or multisource COVID-19 learning [252, 310], as partially illustrated above. Typically, ensemble learning integrates the results from multiple learners such as ensemble trees and XGBoost for COVID-19 learning [51]. Further, multiple methods are sequentially involved to learn specific tasks or data over phases of COVID-19. Other common tasks are to integrate compartmental models with methods like statistical models, classifiers, and DNNs to improve the forecasting of COVID-19 epidemic dynamics and attributes [305, 158]. For example, in [313], a hybrid model predicts the infected and death cases by integrating a genetic algorithm and LSTM into a modified susceptible-infected-quarantined-recovered (SIQR) epidemic model to optimize infection rates and modeling parameters. In [49], a regression tree combined with wavelet transform predicts COVID-19 outbreak and assesses its risk in terms of case numbers. In [16], a baseline method generates a granular ranking (discrimination) of severe respiratory infection or sepsis on the medical records of the general population. A decision-tree-based gradient boosting model then adjusts the former predicted results in subpopulations by aligning it with the published aggregate fatality rates. In addition, other methods and tasks are applied for innovative estimation of COVID-19 pandemic responses. Examples are automated primary care tools to alleviate the shortage of healthcare workers [255], expert systems and chatbots for symptom detection and lessening the mental health burden [181], IoT and smart connecting tools to prevent outbreaks, remotely monitoring patients, and prompting the enforcement of guidelines and administrative orders to contain future outbreaks [103].

#### Discussion

Although hybrid COVID-19 modeling demonstrates an enhanced capability and capacity to characterize COVID-19, the relevant research is still not systematic, comprehensive, or substantial. The gaps and opportunities of novel and effective hybrid COVID-19 modeling apply to hybrid problems, hybrid tasks, hybrid data, and hybrid methods. These may provide extra capacity to address the multiple characteristics and challenges of both the coronavirus and COVID disease, as discussed in Sections 3.3, 3.2 and 3.4.

## 11 Discussion and Opportunities

This review also shows that there are many open challenges, gaps, and opportunities in modeling COVID-19. First, publicly available COVID-19 data is limited, partial, inconsistent, and may contain erroneous, biased, noisy and uncertain observations and statistics due to the limited, imbalanced and non-universal test ability and non-unified reporting standards and statistical errors, especially at the beginning of the epidemic and in underdeveloped regions and countries. Second, the aforementioned long incubation period from infection to the onset of symptoms and the large number of asymptomatic to mild infections make correct and instant reporting difficult and lead to a significant number of undetected and unreported cases, degrading data quality and trust. Third, coronavirus and COVID-19 exhibit unique complexities, which differ from existing epidemics in terms of its transmission, infectivity in ethnic populations, external NPIs, people’s NPI reactions and behavioral changes as a result of COVID-19 mitigation policies, and the rapid and mysterious mutation and spread of coronavirus. Lastly, the modeled problems and areas are fragmented, divided, and evolving, although the modeling techniques and results are highly comprehensive.

These brief observations indicate the critical need to model COVID-19 and the urgency of forming a comprehensive understanding of the progress being made in COVID-19 modeling, the research gaps, and the open issues. This overview is crucial not only for furthering COVID-19 modeling research but also for informing insights on scientific and public strategies and actions to better battle this pandemic and future pandemics.

In the above review of each category of the COVID-19 modeling techniques, a brief discussion has been provided on the main limitations, gaps and opportunities in those areas. Below, we further expand this specific discussion to broad major gaps in the research on modeling COVID-19. In addition, we discuss various open issues and opportunities to discover more informative insights for epidemic, medical, biomedical, political and social management and the further research of COVID-19 and other similar crises.

### 11.1 COVID-19 Modeling Gap Analyses

Here, we highlight two major aspects of modeling gaps. They are the gaps in understanding the virus and disease nature and the gaps in modeling their complexities.

#### 11.1.1 Gaps in understanding the COVID-19 problem nature

Since the coronavirus is new and unique, our knowledge is limited in terms of all aspects of the SARS-CoV-2 virus and COVID-19, such as virus characteristics, epidemiological attributes and dynamics, socioeconomic influence, virus mutations, and so on. Specifically, our poor understanding of the intrinsic and intricate pathological, biomedical and epidemiological attributes of the evolving SARS-CoV-2 and COVID-19 systems limits the modeling attempts and contributions. As a result, our understanding of the virus and disease is still (1) *insufficient* without substantial knowledge and comprehensive evidence on the system complexities; (2) *biased* to specific data, conditions or settings; (3) *shallow* without deep insights into the virus and disease nature; (4) *partial* without a full picture of the SARS-CoV-2 and COVID-19 complexities, in understanding COVID complex systems and their data complexities, as discussed in Sections 3.2 and 3.3 [288, 41].

To address these issues, the COVID-19 modeling has to start with building a comprehensive understanding of the coronavirus nature and the fundamental complexities of COVID-19 ecosystems. Of the many questions to explore, we highlight the following important unknowns. They require cross-disciplinary scientific explorations by integrating medical science, virology, bio-medicine, and data-driven discovery.

- *The hidden nature of SARS-CoV-2 and COVID-19* : How does the coronavirus interact with human and animal hosts? What does the viral genetic system look like? What are the epidemiological attributes of the virus and disease characteristics after infections under different contexts, e.g., demographics, community (population) scenarios, ethnics, seasonality, and weather conditions? What are the high risk factors or high risk factor combinations of COVID-19 infection and mortality? What causes the different levels of symptom onset and how do asymptomatic and mild symptomatic infections differ from severe symptomatic infections?
- *The mysterious mutation mechanisms of SARS-CoV-2* : What genomic and pathological factors determine how the coronavirus transforms from one generation to another and over time? What genomic and pathological mechanisms drive the variations? Why the genetic variants differ from region to region and between populations of different ethnicities?
- *The influence of external factors on coronavirus spread and evolution*: How does the virus evolve under different ethnicities, environmental and intervention (including pharmaceutical and non-pharmaceutical) contexts? How do external factors such as demographics, ethnics, environment, and healthcare quality influence virus transformation? How do personal hygiene, public health systems, public activities, population mobility, and daily commuting affect the coronavirus spread and evolution?
- *Coronavirus adaption to vaccines and drugs*: What are the relations between key factors such as the various vaccines and drugs available for treating COVID-19, the increasing virus mutants and their more contagious new strains, the widespread Delta and Omicron strain outbreaks, and unpredictable resurgences? How do the virus variants adapt to vaccines and drugs? How do the different types of vaccines and vaccination strategies and coverage affect virus evolution?
- *Herd immunity, mitigation vs. zero COVID of the coronavirus*: What is the new COVID normalcy, i.e., should a ‘zero tolerance (zero COVID) approach (or strategy)’ for the virus be the target and eventually remove the SARS-COV-2 like occurred for SARS? Should mitigation (i.e., ‘quotidian existence’) be the new normal such that humans live with the virus, similar to influenza? Or should herd immunity be taken as the eventual approach? For the first two approaches, what is the herd immunity threshold for a manageable normal to coexist with the virus? Where is the manageable risk level of balancing the vaccination rate, public health system capacity, ethnic and community conditions, and acceptable daily numbers of infections and deaths? How does the regional inequality of vaccinations and public health systems affect global recovery?

#### 11.1.2 Gaps in modeling the COVID-19 system complexities

The modeling gaps emanate from both a poor understanding of the nature of the coronavirus and COVID disease and the limitations in modeling their characteristics and complexities. On one hand, even though massive efforts have been made in modeling COVID-19, the existing modeling work is still in its early stage. The weaknesses and limitations of the existing work lie in various aspects, such as

- an average description of the population-wise coronavirus and the disease’s epidemiological characteristics and observations after applying mitigation and control measures, where no fine-grained and microlevel analysis and findings are available;
- a direct application of existing (even very simple and classic) modeling methods without COVID-specific and optimal modeling mechanisms, typically by applying overparameterized or independently pretrained deep neural models or complex statistical and compartment models on low-quality and often small COVID data;
- simple data-driven modeling purely motivated by applying advanced models (typically deep models) on COVID-19 data without a deep incorporation of domain and external knowledge and factors; and
- a purposeful design without a comprehensive design or exploration of the multifaceted COVID-19 characteristics and challenges in one framework or system.

On the other hand, the general applications of existing methods are also unsuitable and incapable of tackling the complexities of the complex coronavirus and COVID disease. Table 11 compares the major modeling methods and their pros and cons in modeling COVID-19. Consequently, it is common that the existing models and their modeling results

**Table 11.** Comparison of COVID-19 Modeling Methods.

| Methods | Pros | Cons |
| --- | --- | --- |
| Time-series analysis | Temporal representations and interaction modelings of periodic and aperiodic components, relations and trends of COVID-19 cases at different states (e.g., new, asymptomatic, infected, recovered, and death) and external temporal factors | Weak capacity in modeling other rich factors (e.g., demographics and clinical attributes) and complex data characteristics (e.g., nonstationarity) and discovering the insight of COVID-19 driving forces and interventions |
| General machine learning | Multifaceted factor and relation analysis, outlier detection, profiling, clustering, classification, prediction and impact analysis of disease diagnosis, cases, external influence, etc. often on small and clean COVID-19 data | Weak capability of modeling poor-quality, multisource and large data, weak but complex interactions, couplings, high-dimensional dependencies, heterogeneity, nonstationarity, and other COVID-19 disease and data challenges |
| Statistical modeling | Modeling COVID distributional dynamics, uncertainty and dependency with analytical explanation and parameter settings, complementing other methods such as epidemic modeling | Requires informative prior knowledge, high modeling and computational complexity on poor-quality COVID data, weak capacity in modeling complex interactions and heterogeneous multisource factors and data |
| Epidemic modeling | Built on epidemiological knowledge on coronavirus and COVID-19, straightforward but domain-friendly and explainable hypothesis test, strong characterization of infection processes, population segmentation, state transitions, nonlinear characterization, and parameter selection | Weak capacity in capturing complex epidemic transmission characteristics, factors, causal relations and processes in COVID-19 developments; strong hypothesis of homogeneous disease transmissions |
| Deep learning | High-performing on large and complex COVID-19 data (e.g., medical imaging)-based case and disease prediction and identification with annotated samples; pretrained models easily adaptable to new tasks; low handcrafting and engineering | Requiring annotated ground truth of COVID-19 learning targets, under-to-over fit small and low-quality COVID-19 data, network vulnerability, poor interpretability, high computational cost |
| Simulation | Imitating and replicating complex COVID-19 mechanisms and processes, cost-effective, reproducible, risk-averse, and manually controllable for purposeful test and optimization, opportunities in disclosing intrinsic realities in the COVID virus and disease | Requiring proper knowledge and hypotheses about the COVID-19 epidemic systems, transmission, and factor interactions; high experimental complexity, inactionable for evolving and random real-life COVID scenarios |
| Hybrid methods | Addressing multi-objectives or multi-tasks; flexible and powerful in fitting small COVID-19 multisource data; taking advantage of multi-methods for combined COVID-19 tasks and data | Requiring an understanding of the constituents for their best ensemble, aligning hybridization to specific COVID-19 challenges with appropriate design complexity, less flexible in combination optimization and explainability |

- often only work for a specific population, cohort-based average estimation, or specific hypothesis of epidemic transmission, losing a personalized applicability to individual cases or scenarios, making it difficult to undertake personalized treatment;
- are too specific and unexpandable to other regions and scenarios, irreproducible and untransferrable to other regions without (significant) changes, making it unsuitable for broad applications;
- over- or under-fit the given data and hypothesis settings, they are often difficult to validate in a fine-grained way and have weak robustness or generalization for a general but deep understanding of the problems; and
- lack the ability and capacity to disclose the intrinsic complexities and general insights about the SARS-CoV-2 virus, COVID-19 disease, and their interventions.

#### 11.1.3 Gaps in actionable COVID-19 modeling and validation

The aforementioned COVID-19 problem nature and system complexities and their mismatch to the existing COVID-19 modeling and results may result in the limited-to-no actionability of existing COVID-19 modeling. Actionability is a common concern in analytics and learning, as well as in other data and model-driven designs, as discussed in domain-driven actionable knowledge discovery and actionable intelligence [45, 38–40, 73, 204]. Actionability is further related to issues including data quality, sampling fairness and bias, algorithmic bias, evaluation bias, result reproductionality, transparency, and interpretability, and more generally ethics [174]. The existing COVID-19 modeling also suffers from the weak actionability of their methodologies, models, evaluation, and results.

In general, such weak actionability may be caused by the aforementioned gaps in understanding the COVID-19 complexities and ecosystem reality. In practice, the weak actionability of existing COVID-19 modeling may be caused and indicated by different reasons or factors along their methodologies, designs, mechanisms, assumptions, settings, data manipulation, feature selection, learning processes, evaluation methods, and result presentation. Examples include:

- the potential flaws in the adopted methodologies, designs, mechanisms, and processes of modeling COVID-19 problems [39, 174, 266];
- a partial-to-inappropriate understanding and quantification of the aforementioned COVID-19 problem nature and system complexities;
- no involvement of the relevant domain knowledge, domain factors, business objectives, and socioeconomic factors in designing the COVID-19 modeling and evaluation systems [40];
- a lack of appropriate data science thinking [42] in data-driven modeling to guide data-driven discovery design, processes, evaluation, and deployment;
- a misunderstanding or misinterpretation leading to a partial capturing of the underlying data characteristics and system complexities in COVID-19, such as in terms of data hierarchy, interactions, heterogeneity, non-IIDness, uncertainty, dimensionality, and dynamics [42];
- an inappropriate feature set with partial, irrelevant, over-simplified, over-manipulated, over-confident, correlated, biased, redundant, or noisy attributes of multi-modal, cross-domain, low-quality, or irregular COVID-19 data;
- inappropriate data manipulation and learning processes such as methodological bias to data fitting, over- or under-sampling, over- or under-fitting, inappropriate conditioning, constraints or assumptions;
- the inappropriate application of pretrained, decoupled and homogeneous deep neural networks in modeling small and limited COVID-19 data;
- the unfairness and bias of data sampling, feature selection, model selection, modeling settings and conditions, test benchmarking, and learning processes;
- evaluation and validation flaws such as inappropriate or biased training-test-validation set partitioning or sampling, inappropriate evaluation metrics, missing business and subjective measures, and the mismatch between modeling conditions and evaluation settings.

### 11.2 Opportunities for AI, Data Science and Machine Learning

There are enormous opportunities and numerous future directions in modeling COVID-19. Examples include (1) fundamentally characterizing COVID system complexities, (2) addressing the aforementioned limitations of existing work on modeling COVID-19, and (3) exploring new directions and alternatives for deeper and more insightful and actionable COVID-19 modeling. These are particularly valid for AI, data science and machine learning, which play a prominent role in data-driven COVID-19 modeling.

#### 11.2.1 Characterizing COVID-19 ecosystem complexities

To discover the mysteries of the COVID virus and disease, the most important opportunities come from understanding their reality and system characteristics and complexities, as discussed in Sections 3.2 and 11.1.1 on COVID-19 disease challenges and gaps in understanding the nature of the problem. By combining domain-driven and data-driven thinking and techniques, there are various directions in characterizing the COVID problem reality and system complexities:

- extracting, representing and distinguishing observable and latent factors and metrics to describe the epidemiological, biological (genomic), medical (clinical and pathological), and social attributes, liveliness, and dynamic processes of the virus, virus mutations, the disease and its variants from other similar viruses and diseases;
- identifying and characterizing external entities and factors (e.g., drugs, vaccines, ethnics, environment) and how they interact with the coronavirus and COVID disease and influence their evolution;
- characterizing and simulating the diversified (e.g., explicit vs. implicit, global vs. local, domain-specific vs. general) interactions and relations between the above-extracted explicit and implicit internal and external factors and their dynamics;
- quantifying and simulating the coronavirus and COVID disease’s system dynamics and genetic mechanisms (e.g., self-organization, genomic expression, genetic crossover and mutation, interaction and adaptation with external environment) in terms of temporal and dependent variables and major transformations;
- simulating and quantifying the virus parasitism, interactions, adaptation, and evolution with human, animal and living hosts on a large scale.

#### 11.2.2 Enhancing deep COVID-19 analytics and learning

To address the modeling gaps discussed in Section 11.1.2 and those rarely and poorly explored areas and challenges discussed in Sections 3.3 and 3.4, we highlight the following major directions to enhance deep COVID-19 analytics and learning.

##### Rarely-to-poorly addressed areas in COVID-19 modeling

First, opportunities to address the areas rarely or poorly addressed in the existing COVID-19 modeling include: (1) characterizing the effective NPIs on the variants of the SARS-CoV-2 virus and comparing them with those on the original strains; (2) quantifying the effect of COVID-19 vaccines, pharmaceutical and NPI interventions on infection control, mobility, mental health, society and the economy, e.g., the efficacy of vaccinated population percentage on herd immunity, and the effect of variable close-contact interactions and individual actions on epidemic de-escalating; (3) balancing NPI strength and socioeconomic recovery, e.g., modeling the effect of full vs partial business close-downs and border control on virus confinement at different stages and for different sectors, and characterizing the effect of increasing daily commuting and workforce movement vs working-from-home and telecommuting on virus confinement; (4) capturing the temporally evolving interplay and interactions between virus propagation and external interventions; and (5) modeling target problems by systemically coupling relevant multisource data and multiple modeling techniques, e.g., by involving pathogen-related, societal, environmental and ethnicity factors and the disparities between developing and developed countries, age groups, and races.

##### Enhancing hybrid COVID-19 modeling

Second, the hybridization of relevant data and techniques offers significant opportunities to improve and expand the existing modeling capacity and results. Examples include integrating (1) coarse-grained and fine-grained modeling, e.g., epidemic modeling by SIR variants to inform the analysis of the effect of further specific NPIs; (2) static and dynamic modeling, e.g., from population-based static epidemic modeling to specific NPI-varying and time-varying case forecasting; (3) observable and hidden factors and relations, e.g., multisource-based attributed modeling with deep abstraction and representation of interactions between multisource factors; (4) local-to-micro-level and global-to-macro-level factors, e.g., involving patient clinical and demographic records with their environmental and socioeconomic context in survival and mortality prediction and medical resource planning; and (5) domain, data and models for domain-specific, interpretable, evidence-based and actionable findings. These typically involve compound modeling objectives, multisource data, and multi-method ensembles.

##### Advancing COVID-19 modeling

Third, another set of new opportunities is to undertake sequential or multi-phase modeling. Examples include (1) from coarse-grained to fine-grained modeling: e.g., applying epidemic models like SIR and SEIR on COVID-19 in the initial stage and then modeling the impact of NPIs, and the mobility and behavioral change of a population on epidemic dynamics; (2) from static to dynamic modeling: e.g., testing constant epidemic parameters and then time-varying settings such as NPI-sensitive varying parameters; and (3) from core to contextual factors: e.g., modeling epidemic processes on case data and then involving pathogen-related, societal and environment (like temperature and humidity) variables to model their influence on epidemic movements.

##### Alternative COVID-19 modeling opportunities

Lastly, various alternative opportunities exist to address the gaps and opportunities in existing COVID-19 modeling. Examples include (1) developing COVID-19-specific modeling methods, benchmarks and evaluation measures to address the challenges of the virus and disease and their data challenges for an intrinsic interpretation of the nature and dynamics of the virus and disease; (2) trans-disciplinarily integrating the relevant domain knowledge and hypotheses from biomedical science, pathology, epidemiology, statistics and computing science to address the multifaceted challenges of the virus, disease, data and modeling and to form a comprehensive understanding of the virus and disease; (3) defining multifaceted modeling objectives and tasks to directly address comprehensive epidemiological, clinical, social, economic or political concerns and their challenges in one framework; and (4) ethical and explainable COVID-19 modeling with privacy-preserving and distributed heterogeneous information integration, augmentation, representation and learning by utilizing personal computing devices (e.g., smart phones) and cloud analytics.

#### 11.2.3 Exploring new epidemic and crisis modeling opportunities

In addition to many specific perspectives, such as hybridizing modeling objectives, data and methods in Section 11.2.2 and addressing the shortcomings in Section 11.1.2, here we highlight some other opportunities that may particularly benefit (from) AI, data science, and machine learning advances.

##### Quantifying the coronavirus nature and complexities

An imperative yet challenging task for the AI, data science and machine learning communities is to ‘quantify’ the reality and complexities of the coronavirus and COVID disease and address the fundamental questions on the virus nature and complexities raised in Section 11.1.1. Building on multi-disciplinary knowledge such as epidemiology, other methods including genetic computing and theories of complex systems and large-scale agent-based epidemic simulation systems are demanding to test and improve genetic, clinical and epidemiological hypotheses and knowledge about the virus and characterize the coronavirus genetic evolution mechanisms.

- *Large-scale COVID epidemiological dynamics*: to obtain quantitative results and verification in relation to the questions in Section 11.1.1, such as: how does a virus evolve, cross-over and mutate? how do billions of coronaviruses interact, compete, and transform over time? and how do environmental factors affect the virus life and genetic evolution?
- *Large-scale human-virus interactions*: to characterize the experiments in relation to questions such as: how does the full population of a country interact with the virus under their varied demographic profiles, hygiene protection habits, health conditions, vaccination conditions, mobility settings, etc. by mimicking their physical census data and circumstances in the real world? what is the vaccination threshold to build herd immunity for a country by considering their specific circumstances? and how to compare the simulation results with the reality of various waves of COVID-19 epidemic occurring in the country?
- *Large-scale intervention influence on human-virus interactions*: to quantify and evaluate questions such as: how do the residents in a country respond to various intervention policies and restrictions on public and household activities over time? how does enforcing or relaxing interventions and restrictions affect the virus spread, infection numbers, and public heath system quality? and what vaccination and intervention preconditions make business reopening possible?

##### Data-driven discovery of COVID mysteries

There are increasing and comprehensive sources of COVID-19 data available publicly and through private providers. Data-driven discovery on this COVID-19 data can substantially leverage other domain-specific research on COVID to disclose the mysteries of COVID.

- *COVID data genomics*: producing the ‘data genomics’ [46] of COVID for a person, country, community or task by automatically extracting and fusing possibly relevant data, e.g., contacts, personal health, mobility, clinical reports, exposure to infected people, and household and public activities in a privacy-preserving manner.
- *COVID data augmentation*: developing new techniques to address the various data quality issues embedded in the data, as discussed in Section 3.3 and novel augmented analytics and learning methods to directly learn from poor quality COVID data.
- *All-purpose representation of COVID attributes*: learning the universal representations [46] on all-relevant COVID data that can be used to describe the full profile of COVID and support diverse learning objectives and tasks in an ethical and privacy-preserving manner.
- *Automated COVID screening and diagnosis*: developing techniques and systems to automatically detect, screen, predict and signal an alert to the potential infection of the virus and disease on COVID data genomics.
- *Virus detection and interaction modeling*: developing personal IoT assistants and sensors to detect the virus, trace its movement and origin, and visualize the ‘COVID net’ showing its propagation paths, interactions and networking with other viruses and hosts.
- *COVID knowledge graph*: generating knowledge graphs showing the ontology of the virus; ontological connections between concepts of the virus; relations between knowledge on the virus and its protection, intervention, treatment and influence; and important highlights such as new knowledge discovered and misinformation detected.
- *COVID safety and risk management* : developing systems and tools (including mobile apps) for an individual’s personal and organizational daily management of COVID safety and risk, e.g., COVID-safe physical and emotional health management, mobility planning, risk estimation and alerting, infection tests, immunity estimation, and compliance management.
- *Metasynthetic COVID decision-support systems*: developing evidence-based decision support systems to fuse real-time and relevant big data, simulate and replay the outbreaks, estimate NPI effects, discover evidence from data and modeling, engage domain experts in the modeling and optimization processes, generate recommendations for decision-making, and support data-driven analytics and the management of severe disasters and emergencies.

## 12 Concluding Remarks

The COVID-19 pandemic’s short-to-long-term influence and impact on public health (both physical and mental health), human daily life, global society, the economy, and geopolitics are unprecedented, lasting, evolving yet quantified and verified. This review paints a comprehensive picture of the field of COVID-19 modeling. The multidisciplinary methods including mathematical modeling, AI, data science, and shallow and deep machine learning on COVID-19 data have deepened our understanding of the SARS-CoV-2 virus and its COVID-19 disease’s complexities and nature. They have contributed to characterizing their propagation, evaluating and assisting in preventive and control measures, detecting COVID-19 infections, predicting next outbreaks, and estimating the influence and impact of COVID-19 on psychological, economic, and social aspects.

The review also highlights the important demands and significant gaps and opportunities of deeply and systemically (1) characterizing COVID-19-related problems and complexities and (2) developing effective, interpretable and actionable models. Our future work is required to characterize, measure, imitate, evaluate and predict broad-based challenges and problems and to proactively and effectively intervene to mitigate them. Such COVID-19 modeling research proposes many significant challenges and opportunities to the multidisciplinary modeling communities in the next decade. These include immediately gaining intrinsic knowledge and proactive insight into the evolving coronavirus and its disease outbreak, infection, transmission, influence and intervention. It is also necessary to tackle future global health, financial, economic, security-related and other black-swan events and disasters.

## Data Availability

All data produced in the present work are contained in the manuscript with further information available at https://datasciences.org/covid19-modeling/.

## Acknowledgements

This work is partially sponsored by the Australian Research Council Discovery grant DP190101079 and the ARC Future Fellowship grant FT190100734. We thank Wenfeng Hou, Siyuan Ren, Yawen Zheng, Qinfeng Wang and Yang Yang for their assistance in the literature collection.

### 13 Appendix: List of modeling keywords

The following lists the base keywords combined with ‘COVID-19’ for the literature search on COVID-19 modeling. The keywords are categorized into machine learning, deep learning, mathematical modeling, epidemic modeling, and other general methods.

Machine learning:

- Machine learning
- Sentiment analysis
- Support vector machine
- Transfer learning
- Random forest
- Decision tree
- Natural language processing
- Artificial neural network

Deep learning:

- Deep learning
- Convolutional neural network
- Neural network
- Long short-term memory
- Deep neural network

Mathematical modeling:

- Mathematical modeling
- Regression model linear regression
- Multivariate statistics
- Logistic regression
- Statistical model
- Bayesian
- Mathematical model
- Cox regression
- Univariate analysis
- Autoregressive integrated moving average
- Poisson regression
- Time-series analysis
- Linear model
- Bivariate analysis
- Multiple linear regression
- Multivariate regression

Epidemic modeling:

- Compartmental modeling
- Susceptible-exposed-infectious-removed
- Susceptible-infectious-recovered
- Transmission model
- Epidemiological model

General and other methods and keywords:

- Artificial intelligence
- Big data
- Compartmental modeling
- Data analytics
- Data mining
- Data science
- Decision making
- Forecasting model
- Predictive model
- Simulation

## Footnotes

1 Section 3.7 Extracting modeling publications [44], more information about the modeling publications can be found in the global scientist response dataset in Kaggle: https://www.kaggle.com/datasets/datascienceslab/covid19-global-scientists-response.

2 Note, here the quoted numerical results are illustrative, which may not represent the state-of-the-art performance and are subject to specific conditions. Interested readers may refer to [44] and specific references for more comprehensive information about how global scientists have responded to model COVID-19.

3 This review complements another literature analysis in [44], and together they are the only initiatives to examine the research map from a cross-disciplinary and cross-domain perspective.

4 WHO Coronavirus (COVID-19) Dashboard: https://covid19.who.int/.

5 COVID-19 Coronavirus Pandemic: https://www.worldometers.info/coronavirus/\# countries, accessed on 27 July 2022.

6 SARS-CoV-2 variants: https://www.who.int/activities/ tracking-SARS-CoV-2-variants.

7 Coronavirus disease (COVID-19): https://www.who.int/health-topics/coronavirus.

8 Tracking SARS-CoV-2 variants: https://www.who.int/activities/tracking-SARS-CoV-2-variants.

9 SARS-CoV-2 variants of concern: https://www.ecdc.europa.eu/en/covid-19/variants-concern.

10 Tracking SARS-CoV-2 variants: https://www.who.int/en/activities/tracking-SARS-CoV-2-variants/.

11 The effects of virus variants on COVID-19 vaccines: https://www.who.int/news-room/feature-stories/detail/the-effects-of-virus-variants-on-covid-19-vaccines?gclid=EAIaIQobChMIgbWS4v3I8QIVRNeWCh3B6wdHEAAYASAAEgLXBfD\_BwE/.

12 Coronavirus research and data: https://ourworldindata.org/coronavirus; CORD-19: The Covid-19 Open Research Dataset: https://www.kaggle.com/allen-institute-for-ai/CORD-19-research-challenge; and global COVID-19 publication data: https://datasciences.org/covid19-modeling/.

13 COVID-19 global literature on coronavirus disease: https://search.bvsalud.org/global-literature-on-novel-coronavirus-2019-ncov/, accessed on 29 July 2022.

14 The literature was informed by the COVID-19 Open Research Dataset (CORD-19) from SemanticScholar https://ai2-semanticscholar-cord-19.s3-us-west-2.amazonaws.com/historical_releases.html.

15 Tracking SARS-CoV-2 variants: https://www.who.int/en/activities/tracking-SARS-CoV-2-variants/.

16 The effects of virus variants on COVID-19 vaccines: https://www.who.int/news-room/feature-stories/detail/the-effects-of-virus-variants-on-covid-19-vaccines?gclid=EAIaIQobChMIgbWS4v3I8QIVRNeWCh3B6wdHEAAYASAAEgLXBfD_BwE/.

17 More details are available in the full technical report on ‘How have global scientists responded to tackling COVID-19?’ [44] and in https://datasciences.org/covid19-modeling on COVID-19 modeling [44]

